# Agreement between mega-trials and smaller trials: a meta-research study

**DOI:** 10.1101/2024.05.09.24307122

**Authors:** Lum Kastrati, Hamidreza Raeisi-Dehkordi, Erand Llanaj, Hugo G. Quezada-Pinedo, Farnaz Khatami, Noushin Sadat Ahanchi, Adea Llane, Renald Meçani, Taulant Muka, John P.A. Ioannidis

## Abstract

**Importance:** Mega-trials can provide large-scale evidence on important questions.

**Objective:** To explore how the results of mega-trails compare to the meta-analysis results of trials with smaller sample sizes.

**Data Sources:** Clinicaltrials.gov was searched for mega-trials until 10.01.2023. PubMed was searched until June 2023 for meta-analyses incorporating the results of the eligible mega-trials.

**Study Selection:** Mega-trials were eligible if they were non-cluster non-vaccine randomized control trials (RCTs); had a sample size over 10,000; and had a peer-reviewed meta-analysis publication presenting results for the primary outcome of the mega-trials and/or all-cause mortality.

**Data Extraction and Synthesis:** For each selected meta-analysis, we extracted results of smaller trials and mega-trials included in the summary effect estimate, and combined them separately using random effects. These estimates were used to calculate the ratio of odds ratios (ROR) between mega-trials and smaller trials in each meta-analysis. Next, the ROR were combined using random-effects. Risk of bias was extracted for each trial included in our analyses (or when not available, assessed only for mega-trials).

**Main Outcomes and Measures:** The main outcomes were the summary ROR for the primary outcome and all-cause mortality between mega-trials and smaller trials. Sensitivity analyses were performed with respect to the time of publishing, masking, weight, type of intervention, and specialty.

**Results:** Of 120 mega-trials identified, 39 (33%) had significant benefits for the primary outcome and 18 (15%) had significant benefits for all-cause mortality for the intervention. In 35 comparisons of primary outcomes (including 85 point estimates from 69 unique mega-trials and 272 point estimatesfrom smaller trials) and 26 comparisons of all-cause mortality (including 70 point estimates from 65 unique mega-trials and 267 point estimates from smaller trials), ROR was 1.00 (95% CI, 0.97-1.04) and 1.00 (95% CI, 0.97-1.04), respectively. For the primary outcomes, smaller trials published before the mega-trials had more favorable results than the mega-trials (ROR 1.05, 95% CI, 1.01-1.10), and than the subsequent smaller trials (ROR 0.91, 95% CI, 0.85-0.96).

**Conclusions and Relevance:** Meta-analyses of smaller studies show in general comparable results with mega-trials, but smaller trials published before the mega-trials give more favorable results than the mega-trials.

## INTRODUCTION

Most randomized comparisons of interventions in medicine use small to modest sample sizes. The need to conduct more “mega-trials” (also called large simple trials) with over 10,000 participants has been long proposed [1, 2]. Mega-trials have been rare in the past, but there has been a renewed interest recently. Several mega-trials have found that certain interventions, like vitamin D supplementation, may not be as effective as previously thought [3, 4]. Conversely, some other mega-trials, such as the ISIS-2 trial on streptokinase and aspirin after myocardial infarction [5] have found favorable results with a major impact on clinical practice. The conduct of mega-trials may be facilitated by the growth of interest in pragmatic research [6, 7], new platforms that facilitate the recruitment of participants [8], and the wide recognition of the limitations of small trials. Therefore, it is important to understand and compare the results of mega-trials to those of smaller trials on the same topic.

Meta-analyses rarely include large trials and small trials have traditionally been considered more susceptible to biases, including more prominent selective reporting biases [9, 10]. Previous literature comparing results of meta-analyses of small trials with subsequent large trials has shown considerable variation in the extent of agreement or disagreement [11-14]. Several factors may contribute to this variation, including different methods used to define agreement, different event rates in the control group of the considered trials (baseline risk), differences in trial quality, and variable susceptibility to bias of the health outcomes under investigation [11]. In previous work, there was also no clear consensus on what constitutes a "large trial". Some [15] have considered the amount of evidence in each trial (inverse of variance or sample size) as a continuum, some have tried to separate trials that have sufficient (e.g., 80%) power to detect plausible effects [16], and some have used arbitrary sample size thresholds to separate large trials, e.g. 1,000 participants [12, 14]. There has been no comprehensive empirical examination that systematically compares the results of mega-trials with sample sizes exceeding 10,000 participants to those of smaller trials.

Here, we aimed to systematically identify such completed registered mega-trials. We then determined whether and which of these mega-trials have been included in meta-analyses for their primary outcomes and/or for mortality outcomes, compared the results of these mega-trials against the combined results of smaller trials, and tried to identify potential factors associated with discrepancies.

## METHODS

### Design and elligibility criteria for mega-trials

This was a meta-research project and the original protocol was registered in OSF (https://osf.io/trsd7). We analysed meta-analyses of clinical trials that have included the mega-trial results in their analysis for calculations of a summary effect for the primary endpoint of the mega-trial. Additionally, we considered data on all-cause mortality as a secondary outcome since it is the most severe and objective outcome.

Mega-trials were considered for analysis if they were non-cluster non-vaccine randomized controlled trials regardless of masking and degree of their design; had sample size exceeding 10,000 participants; and had a peer-reviewed publication presenting the results of the primary endpoint included in a meta-analysis. We excluded cluster trials because the effective sample size is much smaller than the number of participants. We excluded vaccine trials since very large vaccine trials usually have different considerations (e.g., targeting large populations rather than specific sets/types of patients/diseases) and types of outcomes (e.g., often surrogate outcomes of immune response rather than hard clinical outcomes) than mega-trials of other interventions.

For a meta-analysis to be included in the analysis, it had to have a systematic review design and include the results of the mega-trial for the primary outcome along with any number of other trials in obtaining summary effect estimates with the effect size and variance data available (or possible to calculate) for each trial from presented information.

### Search strategy

We performed a search in clinicaltrials.gov for registered completed randomized controlled trials that were characterized as phase 3 or 4 and not have the status of recruitment as “Not yet recruiting” or “Recruiting”. After identifying the clinical trials that fulfilled our eligibility criteria, we searched the first primary publication for these trials that included any primary outcome(s) registered in clinicaltrials.gov. If no primary publication was registered in clinicaltrials.gov, we searched PubMed using the mega-trial name and/or registration number.

Next, in PubMed, we used the option “cited by” and selected further the option “meta-analysis” to identify meta-analyses that had cited the papers of interest. If more than one meta-analysis was identified, we screened them starting with the most recently indexed one and moving backward until a suitable meta-analysis was found that included the mega-trial results in calculations of a summary effect for a primary endpoint of the mega-trial. If summary effect calculations for multiple primary endpoints of the mega-trial were presented in an eligible meta-analysis, we prioritized binary over continuous outcomes; and the primary outcome with the largest number of events in the mega-trial (or the smallest variance if all the endpoints were continuous).

To identify a meta-analysis that included the mega-trial for mortality effect summary estimate calculations, we similarly screened meta-analyses that had cited the earliest main publication of the mega-trials backward in time until the 10^th^ meta-analysis. The same process in PubMed was used to identify citing meta-analyses. If 10 citing meta-analyses did not evaluate the outcome of mortality with eligible, usable data, we excluded that mega-trial from the mortality analysis.

Whenever a mega-trial included more than one active arm versus control, e.g. in a three-arm trial, or two or more different comparisons, e.g. in factorial design, we considered each eligible comparison separately and tried to identify respective meta-analyses.

Clinicaltrials.gov searches were last updated on 10/01/2023. PubMed searches were last updated on June 2023 by independent screeners and were done in duplicate.

### Data extraction

For each selected meta-analysis, we extracted the results of RCTs included in the summary effect estimate that incorporated the effect estimate of the mega-trial. For each selected meta-analysis, we recorded the first author’s name, publication year, eligible endpoint, comparison of intervention versus control, type of masking, topic, and type of intervention. For each trial in each eligible meta-analysis, we recorded the first author’s name or acronym, publication year, total sample size, 2x2 table (or log (odds ratio) and variance thereof, if 2x2 table was not provided) for dichotomized outcomes and standardized mean difference (and variance thereof) for continuous outcomes. We also extracted information, whenever available, on the risk of bias assessments for each included trial based on Cochrane risk of bias tools (original, revised, and version 2).

All data extractions were performed in duplicate, and differences were settled by discussion. For any unsettled discrepancies, a third senior reviewer was invited to arbitrate.

Mega-trials (and their corresponding meta-analyses) that compare two active and overlapping interventions were analyzed as follows: if one intervention is a subset of the other, the subset intervention was considered as the control arm (e.g. in a trial comparing X+Y+Z to X+Y, X+Y was the control arm). If the interventions were not subsets of each other (e.g., X versus Y), the intervention that was approved by the FDA first was considered as the control arm.

### Statistical analysis

In each eligible meta-analysis, we combined the results from non-mega-trials using random effects (and fixed effects as sensitivity analysis) and compared them against the results of the mega-trial. In meta-analyses where several mega-trials were available, the results of the mega-trials were combined using random effects first before being compared against the results of smaller trials. Any cluster trials included in the meta-analysis were considered to be non-mega-trials.

The odds ratio (OR) was used as the metric of choice. Standardized mean differences (for any eligible continuous outcomes) were also converted to ORs [17]. Between-trial heterogeneity in each group of trials (mega-trials and other trials) and for all trials considered together were assessed using τ^2^ between-study variance estimator, Q test, and I^2^ statistics[18].

We obtained the log(relative odds ratio) (logROR) and its variance (the sum of the variances of the logOR in the two groups) between the mega-trial(s) and the smaller trials for each eligible outcome. The logROR estimates were combined then across each outcome using the DerSimonian and Laird random effects calculations.

In all calculations, treatment effects in single trials and meta-analyses thereof were coined consistently in such a way that OR<1 means a more favorable outcome for the intervention than the control arm. In all analyses, heterogeneity was assessed using the τ^2^ between-study variance estimator, Q test, and I^2^ statistics [18].

A sensitivity analysis was performed to assess whether the results were different when non-mega-trials were included in the calculations only if they were published up until (and including) the year of publication of any mega-trials. This analysis more specifically targets the research question whether mega-trials corroborate the results of smaller trials that have been performed before them. A separate analysis also compared the results of non-mega-trials published up until the year of publication of the mega-trial versus those published subsequently.

Separate subgroup analyses were performed for the comparison of results in mega-trials versus other trials according to masking (open-label vs masked); intervention type; specialty (cardiovascular, others); and per heterogeneity (low vs non-low) of the mega-trials. We also performed exploratory meta-regressions considering the same variables (masking, type of outcome, type of intervention, specialty) and also risk of bias in the mega-trials (high versus other), risk of bias in the other trials (proportion at high risk), median number of participants in non-mega-trials, and total number of participants in non-mega-trials.

We also performed exploratory tests for small study effects (Egger’s test) [19], (when there were more than 10 trials).

### Amendments to the original protocol

We extracted information for all mega-trials on whether they found statistically significant or nonsignificant results, and whether they were designed to show noninferiority. In several meta-analyses, some trials that did not pass the 10,000 threshold, but were substantially large to blur the effects. Therefore, we compared the results of mega-trials versus only the smaller trials that weighted less than 1/5^th^ of the least weighted mega-trial; and another sensitivity analysis versus those that weighted less than 1/10^th^ of the least weighted mega-trial. We then further restricted these trials to those published only before or up to the first trial.

Finally, we also assessed the risk of bias for the mega-trials that had not been assessed (or had been assessed using various non-Cochrane tools e.g. Jadad scale) using the Cochrane risk-of-bias tool [20].

## RESULTS

### Identification of mega-trials and the respective meta-analyses

A total of 180 registered completed phase 3 or 4 trials that did not involve vaccines and that had 10,000 or more participants were identified through our search. Among these, 91 were randomized, non-cluster, non-vaccine mega-trials; but 38/91 lacked an appropriate meta-analysis and 2/91 had no published results, leaving 51 mega-trials with an eligible meta-analysis for either primary outcome and/or mortality. Results were compared to smaller trials across 58 meta-analyses for primary outcome (n=35 comparisons) and/or all-cause mortality (n=26 comparisons). In 3 topics, all-cause mortality was the mega-trial’s primary outcome. For 19 mega-trials that had a composite primary outcome, no eligible meta-analysis was identified for the complete composite outcome, therefore the meta-analysis of one of the subsets of the composite outcome with the highest number of events was analysed.

The eligible meta-analyses included estimates from another 30 mega-trials that had randomized, non-cluster design and >10,000 participants but had not been identified in our searches. 26/30 were not registered in clinicaltrials.gov, while the other 2/30 had no listed location (we had identified mega-trials by screening trials registered at clinicaltrials.gov by extracting the trials listed for every country), 1/30 had listed no results in clinicaltrials.gov, and for 1/30 no reason for missingness was identified. These 30 trials with their estimates for primary outcomes (n=20) and all-cause mortality (n=22) were considered in the mega-trials group in all calculations. The meta-analyses included an additional 1 mega-trial that had initially been identified by our search but had no eligible meta-analysis for the primary outcome and/or all-cause mortality but was meta-analyzed for another outcome.

In total, 82 mega-trials estimates were included across all meta-analyses for the primary outcome (n=69) and all-cause mortality (n=65). Detailed information regarding the selection of mega-trials is in Figure 1.

**Figure 1.**
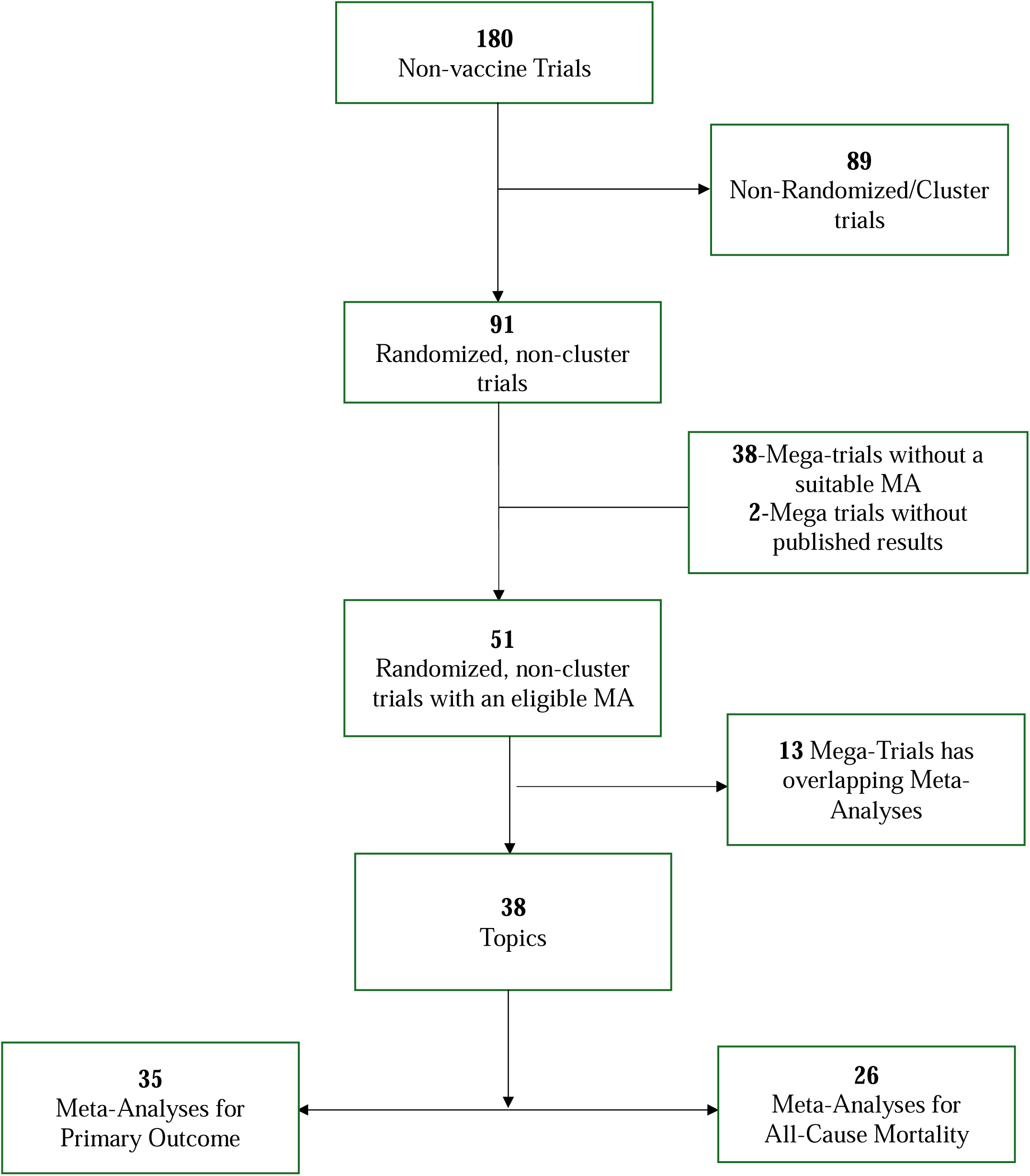
Flow chart of mega-trial selection.

### Characteristics of mega-trials

64/82 mega-trials incuded in our analyses (Table 1, Supplementary Table 1&2) investigated cardiovascular outcomes. 17/82 mega-trials were centered around nutritional interventions, while the remaining covered various other medical interventions intervention types, such as pharmacological treatment (Table 1). Moreover, 15/82 mega-trials were open-label, while the remaining were double-blinded (n=65) or employed varying degrees of masking (n=2) (Table 1). 14/82 mega-trials were judged at high risk of bias. 32/82 had statistically significant results at p<0.05 for the primary outcome (30 favoring the intervention arm) and only 17/82 had statistically significant results at p<0.05 for all-cause mortality (13 favoring the intervention arm) (Table 1, Supplementary Table 1&2).

**Table 1.**
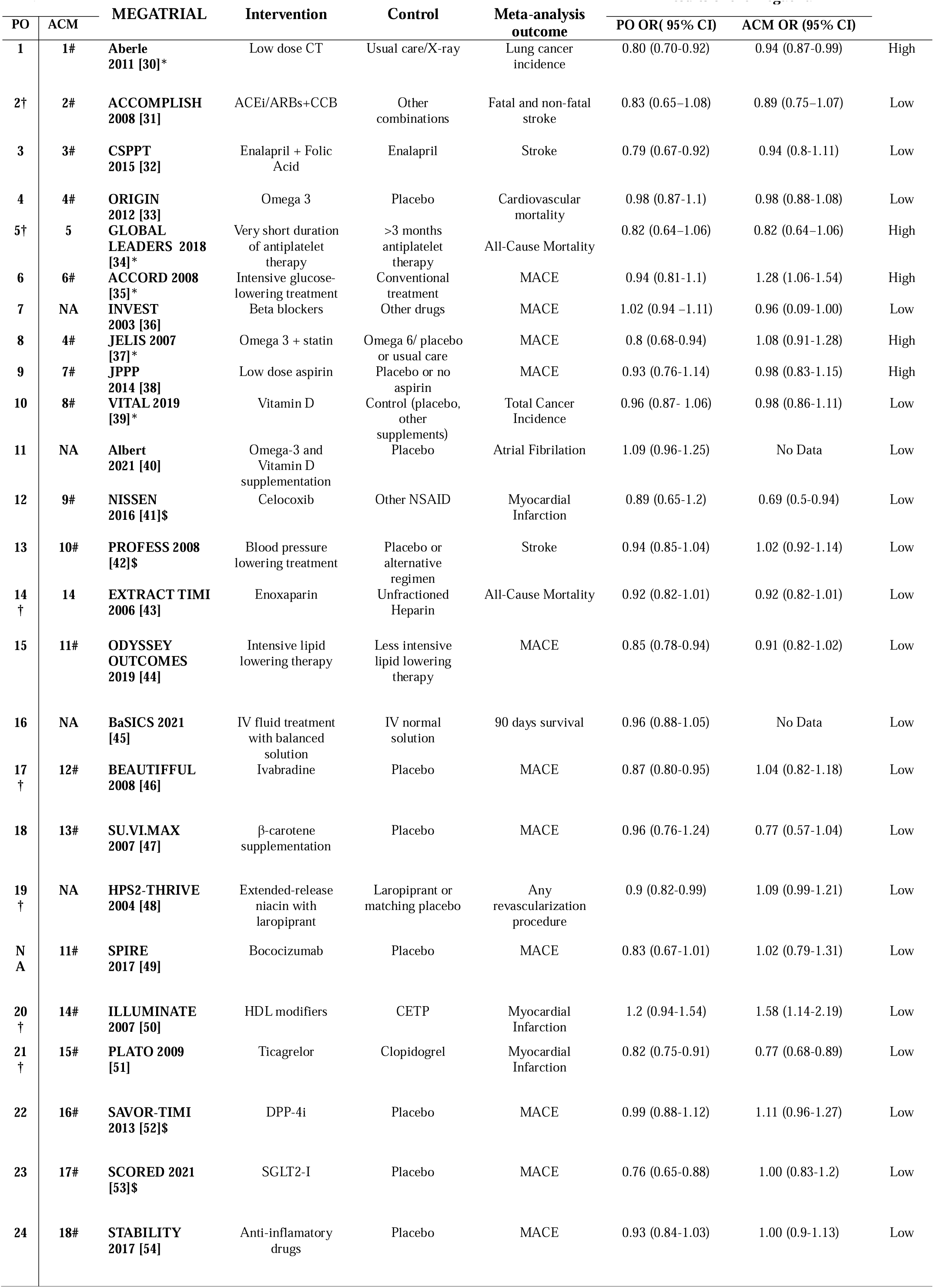
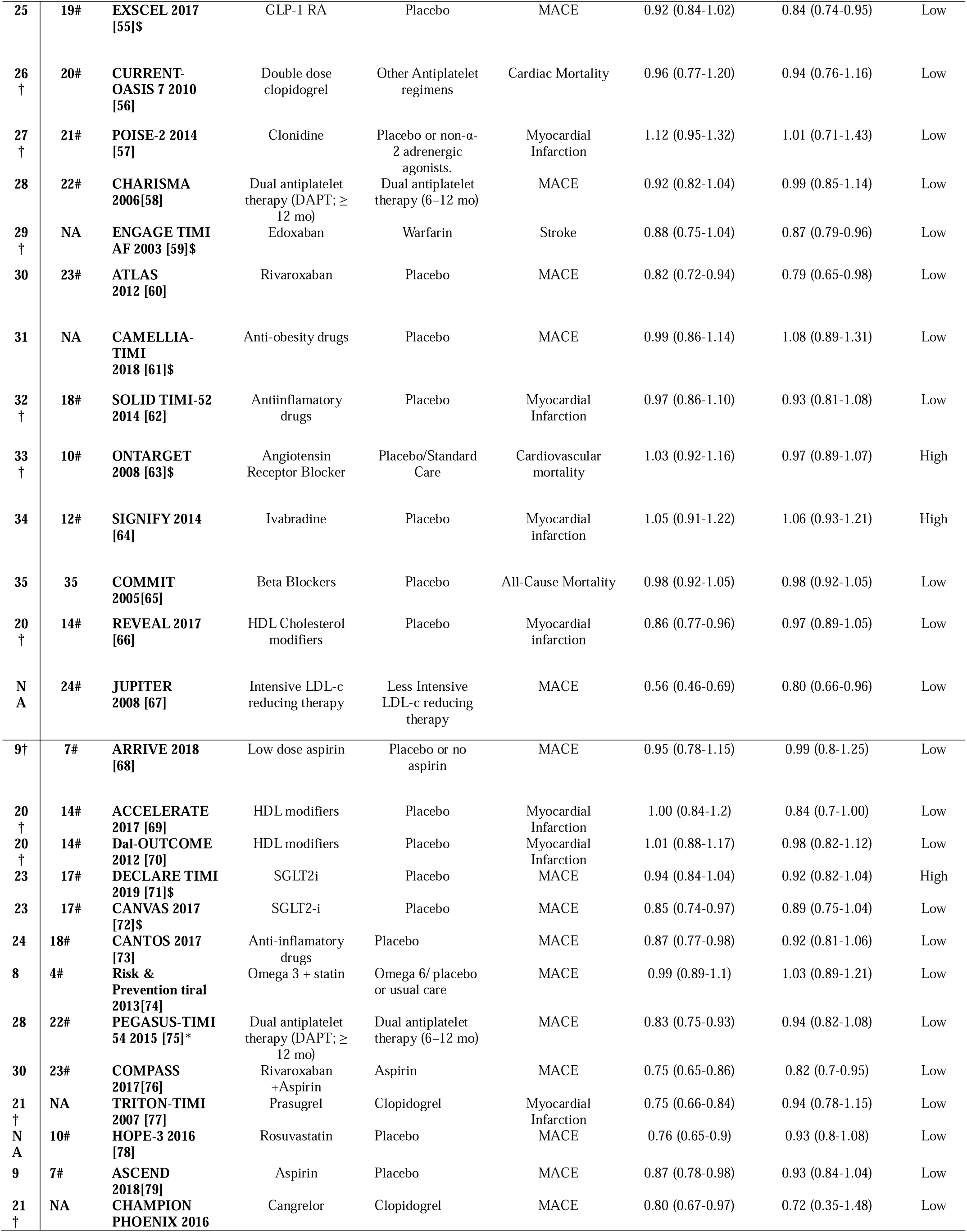
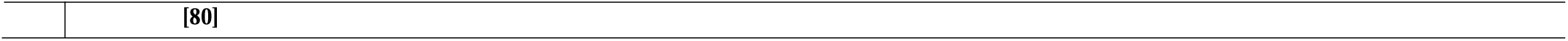
General characteristics of the included mega-trials. **MA ID-** The numbers listed for PO and MA, represent the respective meta-analyses that the respective mega-trials are included and are references for table 2 and 3. **MA-** Meta-analysis, **PO-** Primary Outcome, **ACM-** All-Cause Mortality, **OR-** Odds Ratio, **CI-**Confidence Interval, **ROB-** Risk of Bias, **ACEi-**Angiotenzin Converting Enzyme inhibitor, **ARBs-** Angiotenzing Receptor Blockers, **GPI-** Glycoprotein IIb/IIIa Inhibitors, **DDP-14-** Dipeptidyl Peptidase 4 inhibitors, **SGLT2-I,** Sodium Glucose cotransporter-2 inhibitors, **GLP-1 RA-** Glucagon-like Peptide-1, **LDL-** Low Density Lipoprotein **HDL-** High Density Lipoprotein, **NSAID-** Nonsteroidal Anti-Inflamatory Drugs, **MACE-** Major Adverse Cardiovascular Event, **CETP-** Cholesteryl ester transfer protein. Trials denoted by **\*** were open label. All the other trials were blinded. Trials denoted by **†** have had a composite primary outcome but have been meta-analyzed for only one subset of it. Information on the composite outcome results can be found on Supplementary Table S1. Trials denoted by $ had been designed for proving non-inferiority.

| PO | ACM | MEGATRIAL | Intervention | Control | Meta-analysis outcome | PO OR( 95% CI) | ACM OR (95% CI) |  |
| --- | --- | --- | --- | --- | --- | --- | --- | --- |
| 1 | 1# | Aberle 2011 [30]* | Low dose CT | Usual care/X-ray | Lung cancer incidence | 0.80 (0.70-0.92) | 0.94 (0.87-0.99) | High |
| 2† | 2# | ACCOMPLISH 2008 [31] | ACEi/ARBs+CCB | Other combinations | Fatal and non-fatal stroke | 0.83 (0.65–1.08) | 0.89 (0.75–1.07) | Low |
| 3 | 3# | CSPPT 2015 [32] | Enalapril + Folic Acid | Enalapril | Stroke | 0.79 (0.67-0.92) | 0.94 (0.8-1.11) | Low |
| 4 | 4# | ORIGIN 2012 [33] | Omega 3 | Placebo | Cardiovascular mortality | 0.98 (0.87-1.1) | 0.98 (0.88-1.08) | Low |
| 5† | 5 | GLOBAL LEADERS 2018 [34]* | Very short duration of antiplatelet therapy | >3 months antiplatelet therapy | All-Cause Mortality | 0.82 (0.64–1.06) | 0.82 (0.64–1.06) | High |
| 6 | 6# | ACCORD 2008 [35]* | Intensive glucose-lowering treatment | Conventional treatment | MACE | 0.94 (0.81-1.1) | 1.28 (1.06-1.54) | High |
| 7 | NA | INVEST 2003 [36] | Beta blockers | Other drugs | MACE | 1.02 (0.94 –1.11) | 0.96 (0.09-1.00) | Low |
| 8 | 4# | JELIS 2007 [37]* | Omega 3 + statin | Omega 6/ placebo or usual care | MACE | 0.8 (0.68-0.94) | 1.08 (0.91-1.28) | High |
| 9 | 7# | JPPP 2014 [38] | Low dose aspirin | Placebo or no aspirin | MACE | 0.93 (0.76-1.14) | 0.98 (0.83-1.15) | High |
| 10 | 8# | VITAL 2019 [39]* | Vitamin D | Control (placebo, other supplements) | Total Cancer Incidence | 0.96 (0.87- 1.06) | 0.98 (0.86-1.11) | Low |
| 11 | NA | Albert 2021 [40] | Omega-3 and Vitamin D supplementation | Placebo | Atrial Fibrillation | 1.09 (0.96-1.25) | No Data | Low |
| 12 | 9# | NISSEN 2016 [41]\$ | Celcoxib | Other NSAID | Myocardial Infarction | 0.89 (0.65-1.2) | 0.69 (0.5-0.94) | Low |
| 13 | 10# | PROFESS 2008 [42]\$ | Blood pressure lowering treatment | Placebo or alternative regimen | Stroke | 0.94 (0.85-1.04) | 1.02 (0.92-1.14) | Low |
| 14† | 14 | EXTRACT TIMI 2006 [43] | Enoxaparin | Unfractionated Heparin | All-Cause Mortality | 0.92 (0.82-1.01) | 0.92 (0.82-1.01) | Low |
| 15 | 11# | ODYSSEY OUTCOMES 2019 [44] | Intensive lipid lowering therapy | Less intensive lipid lowering therapy | MACE | 0.85 (0.78-0.94) | 0.91 (0.82-1.02) | Low |
| 16 | NA | BaSICS 2021 [45] | IV fluid treatment with balanced solution | IV normal solution | 90 days survival | 0.96 (0.88-1.05) | No Data | Low |
| 17† | 12# | BEAUTIFUL 2008 [46] | Ivabradine | Placebo | MACE | 0.87 (0.80-0.95) | 1.04 (0.82-1.18) | Low |
| 18 | 13# | SU.VI.MAX 2007 [47] | β-carotene supplementation | Placebo | MACE | 0.96 (0.76-1.24) | 0.77 (0.57-1.04) | Low |
| 19† | NA | HPS2-THRIVE 2004 [48] | Extended-release niacin with laropiprant | Laropiprant or matching placebo | Any revascularization procedure | 0.9 (0.82-0.99) | 1.09 (0.99-1.21) | Low |
| NA | 11# | SPIRE 2017 [49] | Bococizumab | Placebo | MACE | 0.83 (0.67-1.01) | 1.02 (0.79-1.31) | Low |
| 20† | 14# | ILLUMINATE 2007 [50] | HDL modifiers | CETP | Myocardial Infarction | 1.2 (0.94-1.54) | 1.58 (1.14-2.19) | Low |
| 21† | 15# | PLATO 2009 [51] | Ticagrelor | Clopidogrel | Myocardial Infarction | 0.82 (0.75-0.91) | 0.77 (0.68-0.89) | Low |
| 22 | 16# | SAVOR-TIMI 2013 [52]\$ | DPP-4i | Placebo | MACE | 0.99 (0.88-1.12) | 1.11 (0.96-1.27) | Low |
| 23 | 17# | SCORED 2021 [53]\$ | SGLT2-I | Placebo | MACE | 0.76 (0.65-0.88) | 1.00 (0.83-1.2) | Low |
| 24 | 18# | STABILITY 2017 [54] | Anti-inflammatory drugs | Placebo | MACE | 0.93 (0.84-1.03) | 1.00 (0.9-1.13) | Low |
| 25 | 19# | EXSCEL 2017 [55]\$ | GLP-1 RA | Placebo | MACE | 0.92 (0.84-1.02) | 0.84 (0.74-0.95) | Low |
| 26<br>† | 20# | CURRENT-OASIS 7 2010 [56] | Double dose clopidogrel | Other Antiplatelet regimens | Cardiac Mortality | 0.96 (0.77-1.20) | 0.94 (0.76-1.16) | Low |
| 27<br>† | 21# | POISE-2 2014 [57] | Clonidine | Placebo or non-α-2 adrenergic agonists. | Myocardial Infarction | 1.12 (0.95-1.32) | 1.01 (0.71-1.43) | Low |
| 28 | 22# | CHARISMA 2006[58] | Dual antiplatelet therapy (DAPT; ≥ 12 mo) | Dual antiplatelet therapy (6–12 mo) | MACE | 0.92 (0.82-1.04) | 0.99 (0.85-1.14) | Low |
| 29<br>† | NA | ENGAGE TIMI AF 2003 [59]\$ | Edoxaban | Warfarin | Stroke | 0.88 (0.75-1.04) | 0.87 (0.79-0.96) | Low |
| 30 | 23# | ATLAS 2012 [60] | Rivaroxaban | Placebo | MACE | 0.82 (0.72-0.94) | 0.79 (0.65-0.98) | Low |
| 31 | NA | CAMELLIA-TIMI 2018 [61]\$ | Anti-obesity drugs | Placebo | MACE | 0.99 (0.86-1.14) | 1.08 (0.89-1.31) | Low |
| 32<br>† | 18# | SOLID TIMI-52 2014 [62] | Antiinflammatory drugs | Placebo | Myocardial Infarction | 0.97 (0.86-1.10) | 0.93 (0.81-1.08) | Low |
| 33<br>† | 10# | ONTARGET 2008 [63]\$ | Angiotensin Receptor Blocker | Placebo/Standard Care | Cardiovascular mortality | 1.03 (0.92-1.16) | 0.97 (0.89-1.07) | High |
| 34 | 12# | SIGNIFY 2014 [64] | Ivabradine | Placebo | Myocardial infarction | 1.05 (0.91-1.22) | 1.06 (0.93-1.21) | High |
| 35 | 35 | COMMIT 2005[65] | Beta Blockers | Placebo | All-Cause Mortality | 0.98 (0.92-1.05) | 0.98 (0.92-1.05) | Low |
| 20<br>† | 14# | REVEAL 2017 [66] | HDL Cholesterol modifiers | Placebo | Myocardial infarction | 0.86 (0.77-0.96) | 0.97 (0.89-1.05) | Low |
| N<br>A | 24# | JUPITER 2008 [67] | Intensive LDL-c reducing therapy | Less Intensive LDL-c reducing therapy | MACE | 0.56 (0.46-0.69) | 0.80 (0.66-0.96) | Low |
| 9† | 7# | ARRIVE 2018 [68] | Low dose aspirin | Placebo or no aspirin | MACE | 0.95 (0.78-1.15) | 0.99 (0.8-1.25) | Low |
| 20<br>† | 14# | ACCELERATE 2017 [69] | HDL modifiers | Placebo | Myocardial Infarction | 1.00 (0.84-1.2) | 0.84 (0.7-1.00) | Low |
| 20<br>† | 14# | Dal-OUTCOME 2012 [70] | HDL modifiers | Placebo | Myocardial Infarction | 1.01 (0.88-1.17) | 0.98 (0.82-1.12) | Low |
| 23 | 17# | DECLARE TIMI 2019 [71]\$ | SGLT2i | Placebo | MACE | 0.94 (0.84-1.04) | 0.92 (0.82-1.04) | High |
| 23 | 17# | CANVAS 2017 [72]\$ | SGLT2-i | Placebo | MACE | 0.85 (0.74-0.97) | 0.89 (0.75-1.04) | Low |
| 24 | 18# | CANTOS 2017 [73] | Anti-inflammatory drugs | Placebo | MACE | 0.87 (0.77-0.98) | 0.92 (0.81-1.06) | Low |
| 8 | 4# | Risk & Prevention tiral 2013[74] | Omega 3 + statin | Omega 6/ placebo or usual care | MACE | 0.99 (0.89-1.1) | 1.03 (0.89-1.21) | Low |
| 28 | 22# | PEGASUS-TIMI 54 2015 [75]* | Dual antiplatelet therapy (DAPT; ≥ 12 mo) | Dual antiplatelet therapy (6–12 mo) | MACE | 0.83 (0.75-0.93) | 0.94 (0.82-1.08) | Low |
| 30 | 23# | COMPASS 2017[76] | Rivaroxaban +Aspirin | Aspirin | MACE | 0.75 (0.65-0.86) | 0.82 (0.7-0.95) | Low |
| 21<br>† | NA | TRITON-TIMI 2007 [77] | Prasugrel | Clopidogrel | Myocardial Infarction | 0.75 (0.66-0.84) | 0.94 (0.78-1.15) | Low |
| N<br>A | 10# | HOPE-3 2016 [78] | Rosuvastatin | Placebo | MACE | 0.76 (0.65-0.9) | 0.93 (0.8-1.08) | Low |
| 9 | 7# | ASCEND 2018[79] | Aspirin | Placebo | MACE | 0.87 (0.78-0.98) | 0.93 (0.84-1.04) | Low |
| 21<br>† | NA | CHAMPION PHOENIX 2016 | Cangrelor | Clopidogrel | MACE | 0.80 (0.67-0.97) | 0.72 (0.35-1.48) | Low |

### Comparisons of mega-trials versus smaller trials: primary outcome

35 comparisons of mega-trials versus other trials were available. 85 estimates coming from 69 unique mega-trials were considered in these comparisons.

The total number of participants in the mega-trials had a median (IQR) of 15715 (12530-20114) across the 35 topics (Table 2). The total number of smaller trials across these 35 topics was 272 (median 6, range 1-45) (Table 2). The total number of participants in the smaller trials had a median (IQR) of 1639 (297-4128) across the 35 topics. 133/272 smaller trials were published before or up to the year of the first mega-trial of the respective topic. In 7 meta-analyses, the cumulative sample size of all the other, smaller trials exceeded the cumulative sample size of the mega-trials (Table 2).

**Table 2.**
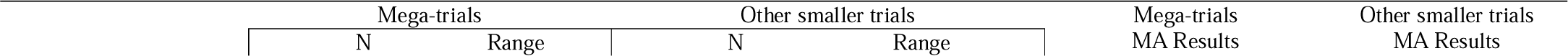
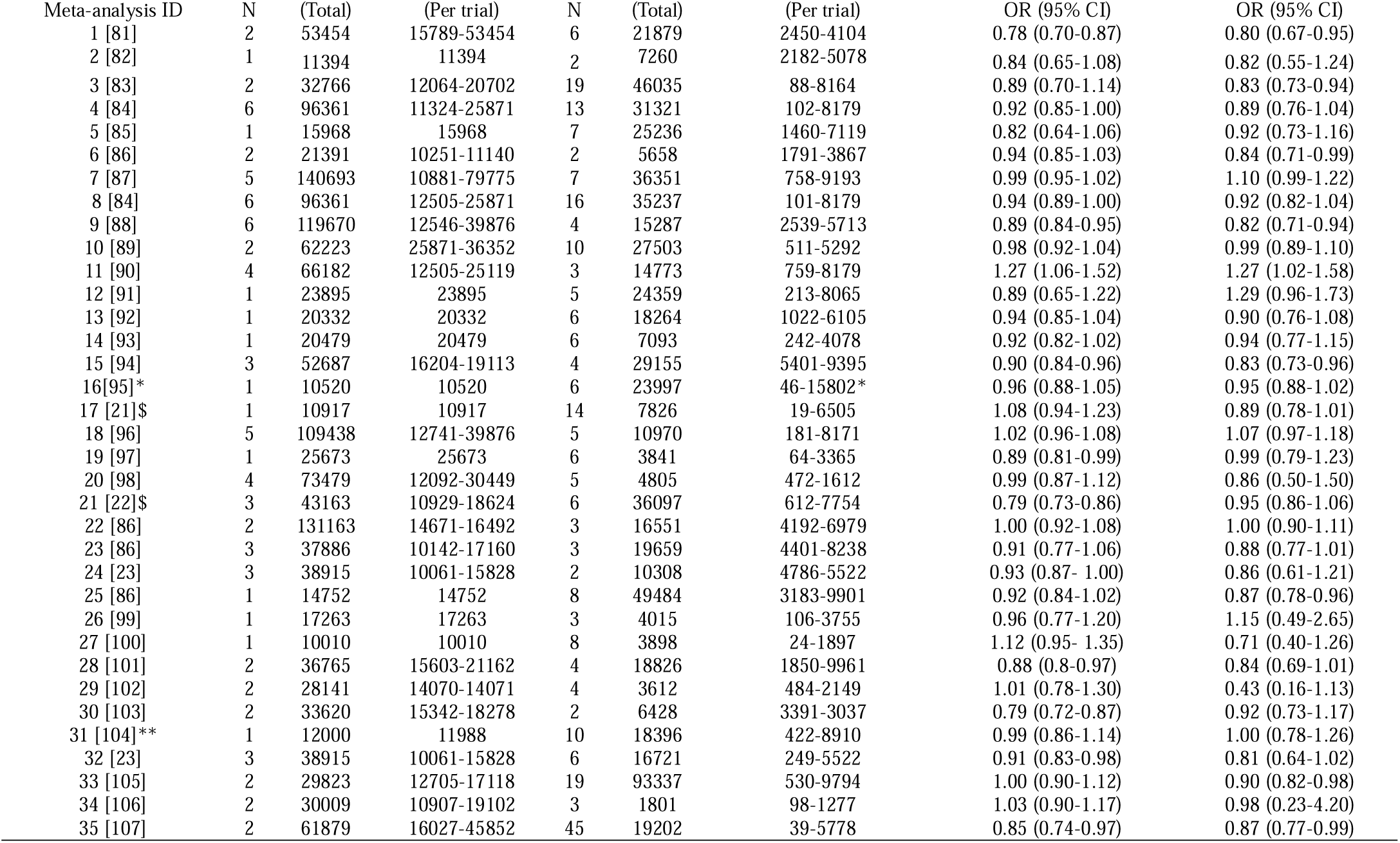
Comparison of results of meta-analyses of mega-trials and other, smaller trials for primary outcome. MA- Meta-analysis **\*\*** No info on the sample size of one of the smaller trials **\***One megatrial was clustered therefore was accounted for as a smaller trial $ Comparisons with significant differences between mega-trials and smaller trial

| Meta-analysis ID | N | (Total) | (Per trial) | N | (Total) | (Per trial) | OR (95% CI) | OR (95% CI) |
| --- | --- | --- | --- | --- | --- | --- | --- | --- |
| 1 [81] | 2 | 53454 | 15789-53454 | 6 | 21879 | 2450-4104 | 0.78 (0.70-0.87) | 0.80 (0.67-0.95) |
| 2 [82] | 1 | 11394 | 11394 | 2 | 7260 | 2182-5078 | 0.84 (0.65-1.08) | 0.82 (0.55-1.24) |
| 3 [83] | 2 | 32766 | 12064-20702 | 19 | 46035 | 88-8164 | 0.89 (0.70-1.14) | 0.83 (0.73-0.94) |
| 4 [84] | 6 | 96361 | 11324-25871 | 13 | 31321 | 102-8179 | 0.92 (0.85-1.00) | 0.89 (0.76-1.04) |
| 5 [85] | 1 | 15968 | 15968 | 7 | 25236 | 1460-7119 | 0.82 (0.64-1.06) | 0.92 (0.73-1.16) |
| 6 [86] | 2 | 21391 | 10251-11140 | 2 | 5658 | 1791-3867 | 0.94 (0.85-1.03) | 0.84 (0.71-0.99) |
| 7 [87] | 5 | 140693 | 10881-79775 | 7 | 36351 | 758-9193 | 0.99 (0.95-1.02) | 1.10 (0.99-1.22) |
| 8 [84] | 6 | 96361 | 12505-25871 | 16 | 35237 | 101-8179 | 0.94 (0.89-1.00) | 0.92 (0.82-1.04) |
| 9 [88] | 6 | 119670 | 12546-39876 | 4 | 15287 | 2539-5713 | 0.89 (0.84-0.95) | 0.82 (0.71-0.94) |
| 10 [89] | 2 | 62223 | 25871-36352 | 10 | 27503 | 511-5292 | 0.98 (0.92-1.04) | 0.99 (0.89-1.10) |
| 11 [90] | 4 | 66182 | 12505-25119 | 3 | 14773 | 759-8179 | 1.27 (1.06-1.52) | 1.27 (1.02-1.58) |
| 12 [91] | 1 | 23895 | 23895 | 5 | 24359 | 213-8065 | 0.89 (0.65-1.22) | 1.29 (0.96-1.73) |
| 13 [92] | 1 | 20332 | 20332 | 6 | 18264 | 1022-6105 | 0.94 (0.85-1.04) | 0.90 (0.76-1.08) |
| 14 [93] | 1 | 20479 | 20479 | 6 | 7093 | 242-4078 | 0.92 (0.82-1.02) | 0.94 (0.77-1.15) |
| 15 [94] | 3 | 52687 | 16204-19113 | 4 | 29155 | 5401-9395 | 0.90 (0.84-0.96) | 0.83 (0.73-0.96) |
| 16[95]* | 1 | 10520 | 10520 | 6 | 23997 | 46-15802* | 0.96 (0.88-1.05) | 0.95 (0.88-1.02) |
| 17 [21]\$ | 1 | 10917 | 10917 | 14 | 7826 | 19-6505 | 1.08 (0.94-1.23) | 0.89 (0.78-1.01) |
| 18 [96] | 5 | 109438 | 12741-39876 | 5 | 10970 | 181-8171 | 1.02 (0.96-1.08) | 1.07 (0.97-1.18) |
| 19 [97] | 1 | 25673 | 25673 | 6 | 3841 | 64-3365 | 0.89 (0.81-0.99) | 0.99 (0.79-1.23) |
| 20 [98] | 4 | 73479 | 12092-30449 | 5 | 4805 | 472-1612 | 0.99 (0.87-1.12) | 0.86 (0.50-1.50) |
| 21 [22]\$ | 3 | 43163 | 10929-18624 | 6 | 36097 | 612-7754 | 0.79 (0.73-0.86) | 0.95 (0.86-1.06) |
| 22 [86] | 2 | 131163 | 14671-16492 | 3 | 16551 | 4192-6979 | 1.00 (0.92-1.08) | 1.00 (0.90-1.11) |
| 23 [86] | 3 | 37886 | 10142-17160 | 3 | 19659 | 4401-8238 | 0.91 (0.77-1.06) | 0.88 (0.77-1.01) |
| 24 [23] | 3 | 38915 | 10061-15828 | 2 | 10308 | 4786-5522 | 0.93 (0.87- 1.00) | 0.86 (0.61-1.21) |
| 25 [86] | 1 | 14752 | 14752 | 8 | 49484 | 3183-9901 | 0.92 (0.84-1.02) | 0.87 (0.78-0.96) |
| 26 [99] | 1 | 17263 | 17263 | 3 | 4015 | 106-3755 | 0.96 (0.77-1.20) | 1.15 (0.49-2.65) |
| 27 [100] | 1 | 10010 | 10010 | 8 | 3898 | 24-1897 | 1.12 (0.95- 1.35) | 0.71 (0.40-1.26) |
| 28 [101] | 2 | 36765 | 15603-21162 | 4 | 18826 | 1850-9961 | 0.88 (0.8-0.97) | 0.84 (0.69-1.01) |
| 29 [102] | 2 | 28141 | 14070-14071 | 4 | 3612 | 484-2149 | 1.01 (0.78-1.30) | 0.43 (0.16-1.13) |
| 30 [103] | 2 | 33620 | 15342-18278 | 2 | 6428 | 3391-3037 | 0.79 (0.72-0.87) | 0.92 (0.73-1.17) |
| 31 [104]** | 1 | 12000 | 11988 | 10 | 18396 | 422-8910 | 0.99 (0.86-1.14) | 1.00 (0.78-1.26) |
| 32 [23] | 3 | 38915 | 10061-15828 | 6 | 16721 | 249-5522 | 0.91 (0.83-0.98) | 0.81 (0.64-1.02) |
| 33 [105] | 2 | 29823 | 12705-17118 | 19 | 93337 | 530-9794 | 1.00 (0.90-1.12) | 0.90 (0.82-0.98) |
| 34 [106] | 2 | 30009 | 10907-19102 | 3 | 1801 | 98-1277 | 1.03 (0.90-1.17) | 0.98 (0.23-4.20) |
| 35 [107] | 2 | 61879 | 16027-45852 | 45 | 19202 | 39-5778 | 0.85 (0.74-0.97) | 0.87 (0.77-0.99) |

Detailed information with forest plots on all of the 35 meta-analyses appears in Supplementary Material 1. In the summary analysis, there was no noteworthy discrepancy observed between the results of the mega-trials and those of smaller trials (summary ROR 1.00, 95% confidence interval [CI] 0.97-1.04; I^2^=0.0, P-value for heterogeneity=0.478; Figure 2). In two instances when comparing ivabradine to placebo and new adenosine diphosphate receptor agonist versus clopidogrel, the disagreement between the mega-trials and the respective smaller trials was beyond chance (ROR 1.21, 95% CI, 1.0-1.47 and ROR 0.83, 95% CI, 0.73, 0.95, respectively) [21, 22].

**Figure 2.**
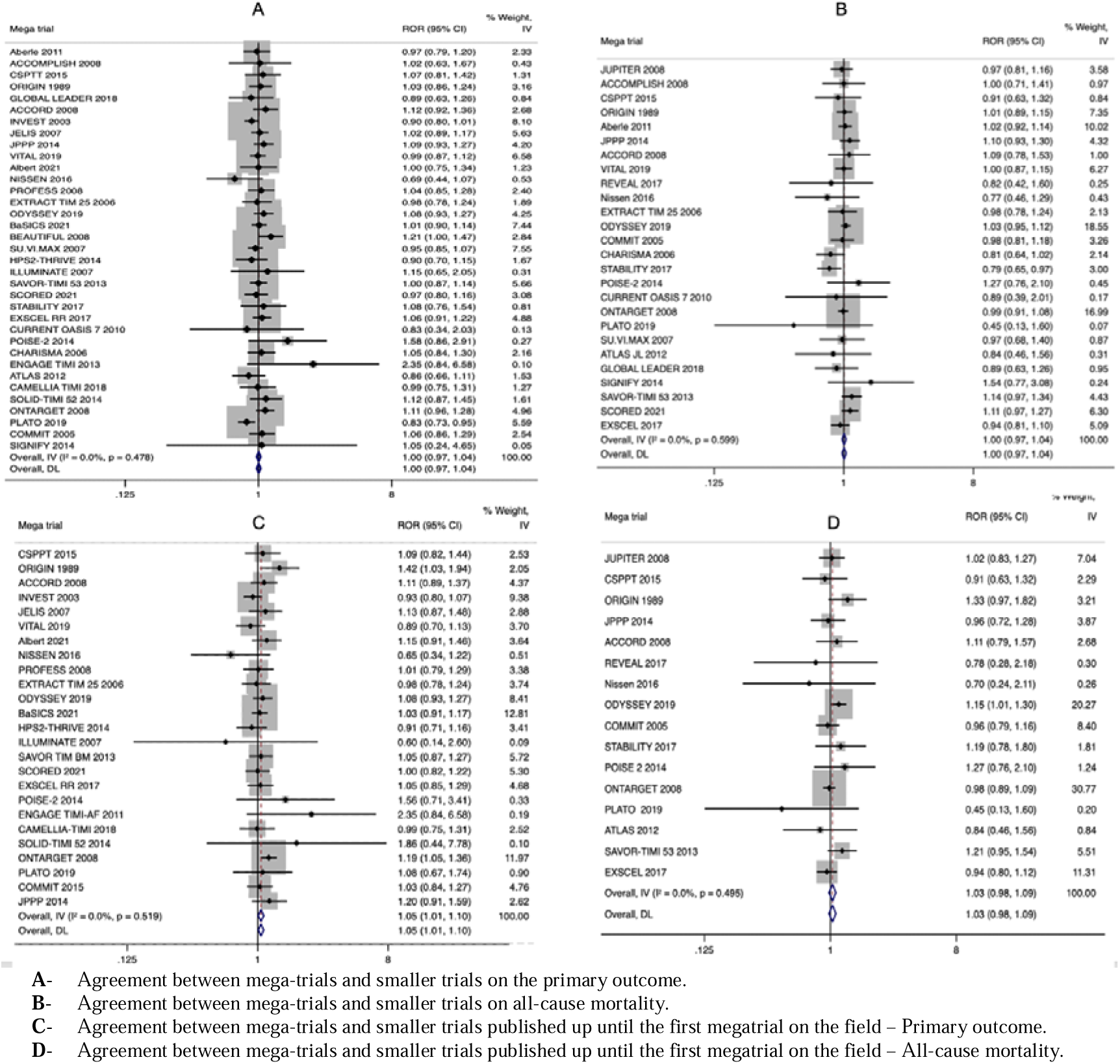
Agreement between mega-trials and smaller trials on the primary outcome and all-cause mortality.

### Comparisons of mega-trials versus smaller trials: all-cause mortality

26 comparisons of mega-trials versus other trials were available. 70 estimates coming from 65 unique mega-trials were considered in these comparisons (Table 3). The total number of participants in all of the mega-trials had a median of 15919 (IQR, 12524-18857).

**Table 3.** Comparison of results of meta-analyses of mega-trials and smaller trialls for all-cause mortality. MA- Meta-analysis *All-Cause Mortality was the Primary Outcome $ Comparison with significant differences between mega-trials and smaller trial

| Mega-trials | Smaller trials | Mega-trials MA Results | Smaller MA Results |
| --- | --- | --- | --- |

**Table 3. Comparison of results of meta-analyses of mega-trials and smaller trials for all-cause mortality**
| MA ID | N | N<br>(Total) | Range<br>(Per trial) | N | N<br>(Total) | Range<br>(Per trial) | OR (LC-IUCi) | OR (LC-IUCi) |
| --- | --- | --- | --- | --- | --- | --- | --- | --- |
| 24# [24] | 2 | 30573 | 12705-17802 | 14 | 51672 | 505-9270 | 0.80 (0.75-1.01) | 0.90 (0.82-0.99) |
| 2# [82] | 1 | 11506 | 11506 | 2 | 7340 | 2199-5141 | 0.89 (0.75-1.07) | 0.89 (0.67-1.19) |
| 3# [108] | 1 | 20702 | 20702 | 3 | 5351 | 553-3090 | 0.94 (0.80-1.11) | 1.03 (0.74-1.42) |
| 4# [84] | 5 | 80889 | 12513-25871 | 18 | 35548 | 101-8179 | 0.97 (0.92-1.03) | 0.96 (0.86-1.07) |
| 1# [81] | 2 | 43501 | 15970-27531 | 6 | 22119 | 2509-4143 | 0.95 (0.90-1.00) | 0.93 (0.85-1.02) |
| 7# [88] | 6 | 120270 | 12546-39876 | 4 | 15287 | 2539-5713 | 0.99 (0.92-1.06) | 0.90 (0.78-1.04) |
| 6# [109] | 2 | 21391 | 10251-11140 | 10 | 13576 | 43-5238 | 1.08 (0.79-1.48) | 0.99 (0.89-1.11) |
| 8# [89] | 2 | 31105 | 18177-12928 | 4 | 6488 | 1015-2650 | 0.94 (0.87-1.02) | 0.94 (0.84-1.04) |
| 14# [110] | 4 | 72479 | 11092-30449 | 4 | 3834 | 130-1612 | 1.02 (0.91-1.14) | 1.25 (0.65-2.39) |
| 9# [91] | 1 | 239553 | 239553 | 4 | 24248 | 916-8067 | 0.69 (0.50-0.94) | 0.90 (0.61-1.35) |
| 15[93]* | 1 | 20479 | 20479 | 6 | 7093 | 242-4078 | 0.92 (0.82-1.02) | 0.94 (0.77-1.15) |
| 11# [24] | 12 | 181434 | 10001-27564 | 47 | 140831 | 250-9270 | 0.93 (0.88-0.98) | 0.90 (0.85-0.95) |
| 12#[106] | 2 | 30019 | 10917-19102 | 13 | 3408 | 19-1277 | 1.05 (0.96-1.15) | 0.68 (0.35-1.34) |
| 37 [107]* | 2 | 61879 | 16027-45852 | 45 | 19202 | 39-5778 | 0.85 (0.74-0.97) | 0.87 (0.77-0.99) |
| 22# [101] | 3 | 46726 | 9961-21162 | 3 | 8837 | 1822-5045 | 1.02 (0.88-1.17) | 1.12 (0.86-1.47) |
| 18# [23]\$ | 3 | 38915 | 10061-15828 | 7 | 21476 | 249-5522 | 0.96 (0.89-1.04) | 1.21 (1.02-1.45) |
| 19#[111] | 1 | 14932 | 14932 | 8 | 41963 | 355-9901 | 0.84 (0.74-0.95) | 0.89 (0.82-0.96) |
| 21# [100] | 1 | 10010 | 10010 | 12 | 4071 | 20-1897 | 1.01 (0.72-1.44) | 0.80 (0.56-1.13) |
| 20#[99] | 1 | 17263 | 17263 | 2 | 4831 | 1076-3755 | 0.94 (0.76-1.17) | 1.06 (0.49-2.30) |
| 10# [112] | 4 | 65400 | 12705-20332 | 32 | 94942 | 80-9794 | 1.00 (0.95-1.06) | 1.01 (0.95-1.07) |
| 15#[22] | 1 | 18624 | 18624 | 1 | 661 | 661 | 0.77 (0.67-0.88) | 1.72 (0.50-5.96) |
| 16#[111] | 2 | 31163 | 14671-16492 | 5 | 17119 | 91-6979 | 1.08 (0.97-1.19) | 0.95 (0.84-1.07) |
| 17#[113] | 3 | 37886 | 10142-17160 | 8 | 42188 | 1222-8246 | 0.93 (0.85-1.01) | 0.84(0.76-0.93) |
| 13# [96] | 5 | 81816 | 11550-22071 | 1 | 840 | 840 | 1.08 (0.99-1.18) | 1.11 (0.79-1.58) |
| 23#[103] | 2 | 33602 | 15342-18278 | 1 | 3037 | 3037 | 0.81 (0.72-0.92) | 0.96 (0.53-1.72) |
| 5 [85]* | 1 | 15968 | 15698 | 7 | 25236 | 1460-7119 | 0.82 (0.64-1.06) | 0.92 (0.73-1.16) |

The total number of smaller trials in these 26 meta-analyses was 267 (median per meta-analysis was 6, range 1-47). The cumulative number of participants from smaller trials was 1132 (IQR, 250-4038). 117/268 smaller trials were published before or up to the year of the first mega-trial of the respective topic. In 5 meta-analyses the cumulative number of participants in the other, smaller trials exceeded the total number of participants in the mega-trials (Table 3).

Comprehensive details and forest plots about the 26 meta-analyses appear in Supplementary Material 2.

In the summary analysis, no difference existed between the outcomes of the mega-trials and those of the smaller trials (summary ROR 1.00, 95% CI: 0.97-1.04; I^2^=0, P-value for heterogeneity=0.60; Figure 3). In one instance testing effects of anti-inflammatory versus placebo in patients with coronary artery diseases the results differed beyond chance between mega-trials and the other, smaller trials (ROR, 0.79, 95% CI, 0.65-0.97), with mega-trial showing no effect while meta-analysis of smaller trials an increased risk [23].

### Sensitivity analyses

Smaller trials showed a significant trend for larger effects when compared to mega-trials when they were published before the first megatrial (ROR 1.05, 95% CI, 1.01-1.10) for the primary outcome and a non-significant trend for all-cause mortality 1.03 (95% CI, 0.98-1.09) (Figure 4a & 4b). Results of smaller trials published before the mega-trial showed significantly higher benefits as compared to smaller trials published subsequently (ROR 0.91, 95% CI, 0.85-0.96) for primary outcome and similar trend (ROR 0.94, 95% CI, 0.87-1.02) for all-cause mortality (Supplementary Figure S1).

No difference was seen when results were pooled using fixed effects and other subgroup analyses and meta-regressions were also non-revealing (Supplementary Figures S2-S8).

No small-study effects were found for the meta-analyses for the primary outcome and one meta-analysis had a significant result for all-cause mortality [24].

### Mega-trials not included in meta-analyses

Of the 38 mega-trials that were otherwise eligible but for which we could not retrieve any meta-analysis that included them (Supplementary Table 4), 9/38 had a statistically significant benefit at p<0.05 for the primary outcome (all favoring the intervention) and 5/38 had significant results for all-cause mortality (all favoring intervention).

### Significance and non-inferiority across all mega-trials

In total, we analysed and/or described the results from 120 mega-trials. 41/120 showed a significant result for the primary outcome and 22/120 for all-cause mortality (33/120 (28%) and 18/120 (15%), respectively favoring the intervention over control). For studies with non-inferiority designs (n=17/120), 15/17 had reached noninferiority and 2/18 had significantly better results in the experimental arm versus the control for the primary outcome (Table 1, Supplementary Table 1&2).

## DISCUSSION

Overall, outcomes from meta-analyses of other, smaller clinical trials aligned on average with those of mega-trials in the clinical topics that we examined. However, mega-trials tended to have less favorable results than the smaller trials that preceded them timewise; and smaller trials published after the mega-trials tended to have less favorable results than the smaller trials published before the mega-trials and aligned with the mega-trials. Most mega-trials do not show statistically significant benefits for the primary outcome of interest, and statistically significant benefits for mortality are rare. Mega-trials are not available for most medical topics. Given that small trials and their meta-analyses may give unreliable, inflated estimates of benefit, mega-trials, or at least substantially large trials with sufficient power, may need to be considered and performed more frequently.

The diminished benefits in late smaller trials versus early small trials are consistent also with prior meta-research studies that have shown that the reported effects of interventions change over time, with wider oscillations of results in early studies [25]. It has been observed that it is more frequent for treatment effects to decrease rather than increase over time [26-28]. In our examined topics, the mega-trials may have corrected some inflated effects seen in the earlier trials that preceded them. Then, the subsequent trials might have been more aligned with what mega-trials had shown since the mega-trials are likely to have been considered very influential.

Previous meta-research assessments have shown different levels of agreement between the results of meta-analyses of smaller trials and large clinical trials. For example, Cappelleri et al. reported compatible results of meta-analysis of smaller studies with the results of large trials, although discrepancies in their results were found in up to 10% of the cases [11]. However, other meta-studies on this topic showed larger differences with a discrepancy rate of up to 39% [13]. These previous studies used a definition of a large trial having enrolled 1,000 participants or more. In contrast, we used a sample size of 10,000 participants to define a mega-trial, and therefore having a larger power to detect effects.

A limitation of our study is that several early mega-trials are not included in the clinicaltrials.gov registry. Nevertheless, by screening the eligible meta-analysis we were able to identify several of these trials, and they were considered in our calculations. No differences in significance for primary outcome and mortality were found between mega-trials registered in clinicaltrials.gov and not, which suggests that if other meta-trials were missed, this would likely not affect our findings. Our comparative results versus smaller trials still do not include all mega-trials, since for some mega-trials retrieved in clinicaltrials.gov we found no relevant meta-analysis where they had been included. However, we did examine the main conclusions of these mega-trials and they also had low rates of statistically significant results. Therefore, we can conclude that mega-trials in general tend to give “negative” results for tested interventions. Mega-trials are unlikely to be launched unless there is genuine equipoise. Nevertheless, the low rate of significant benefits, as opposed to the much higher rates of favorable results seen in typical phase 3 trials is remarkable [29]. For example, one assessment found superiority results for 80% of industry-funded and 44% of publicly-funded phase 3 trials in oncology [29].

Results from smaller and larger trials are needed as they both contribute to the generation of evidence. Confirmatory – well-designed and well-powered large studies are needed for common clinical questions. Mega-trials are done very sparingly to date, but it would be beneficial to add more such trials to the clinical research armamentarium.

## Data Availability

All data in the present study are available upon reasonable request to the authors

## Notes

**Conflicts of interest: None**

**Funding:** The work of John Ioannidis is supported by an unrestricted gift from Sue and Bob O’Donnell to Stanford University. The work of Lum Kastrati is supported by Swiss Government Scholarship for Excellence and funding from University of Bern and Insel Spital.

### Competing Interest Statement

The authors have declared no competing interest.

### Clinical Protocols

https://osf.io/trsd7

### Funding Statement

This study did not receive any funding

