## Supplementary Figures for "Agreement between mega-trials and smaller trials: a meta-research study"

**Suplementary Figures**

**Figure S1a.** Agreement between smaller trials prior and after the publication of the first mega-trial – Primary outcome.

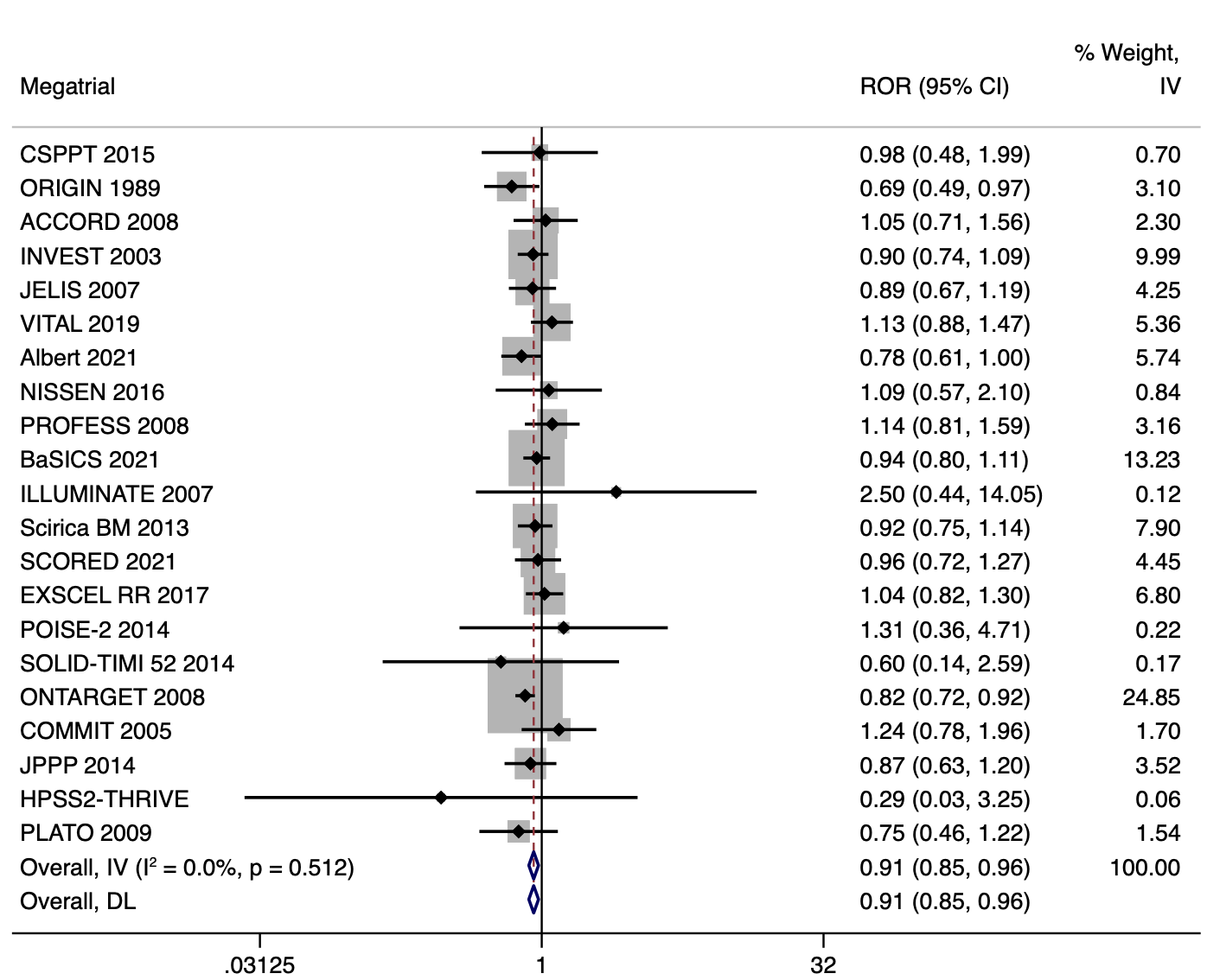

**Figure S1b.** Agreement between smaller trials prior and after the publication of the first mega-trial – All-cause mortality.

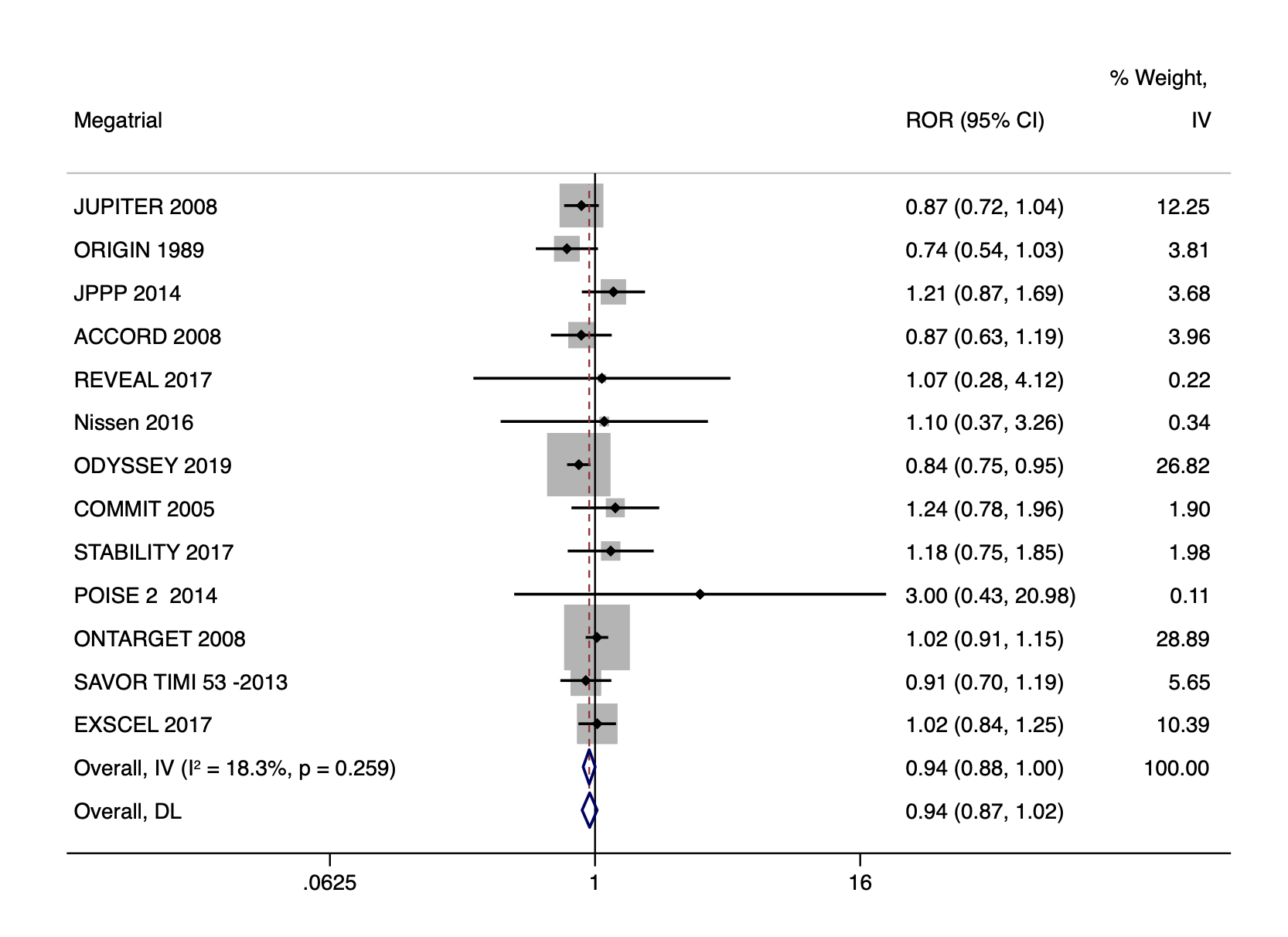

**Figure S2a**. Agreement between mega-trials and smaller trials with 1/5 of the least weighted mega-trial – Primary outcome.

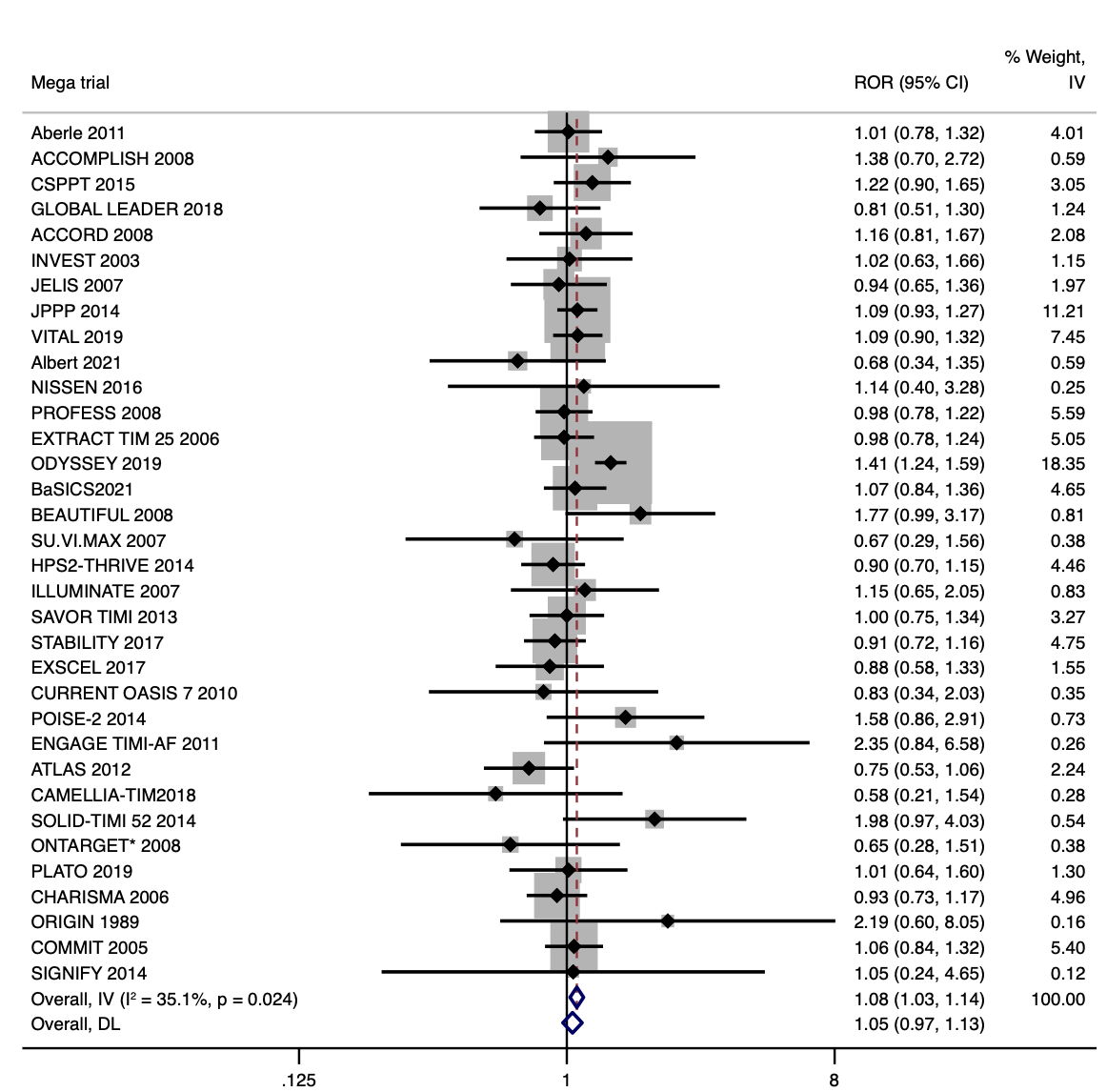

**Figure S2b**. Agreement between mega-trials and smaller trials with 1/5 of the least weighted mega-trial published prior or up until the mega-trial – Primary outcome.

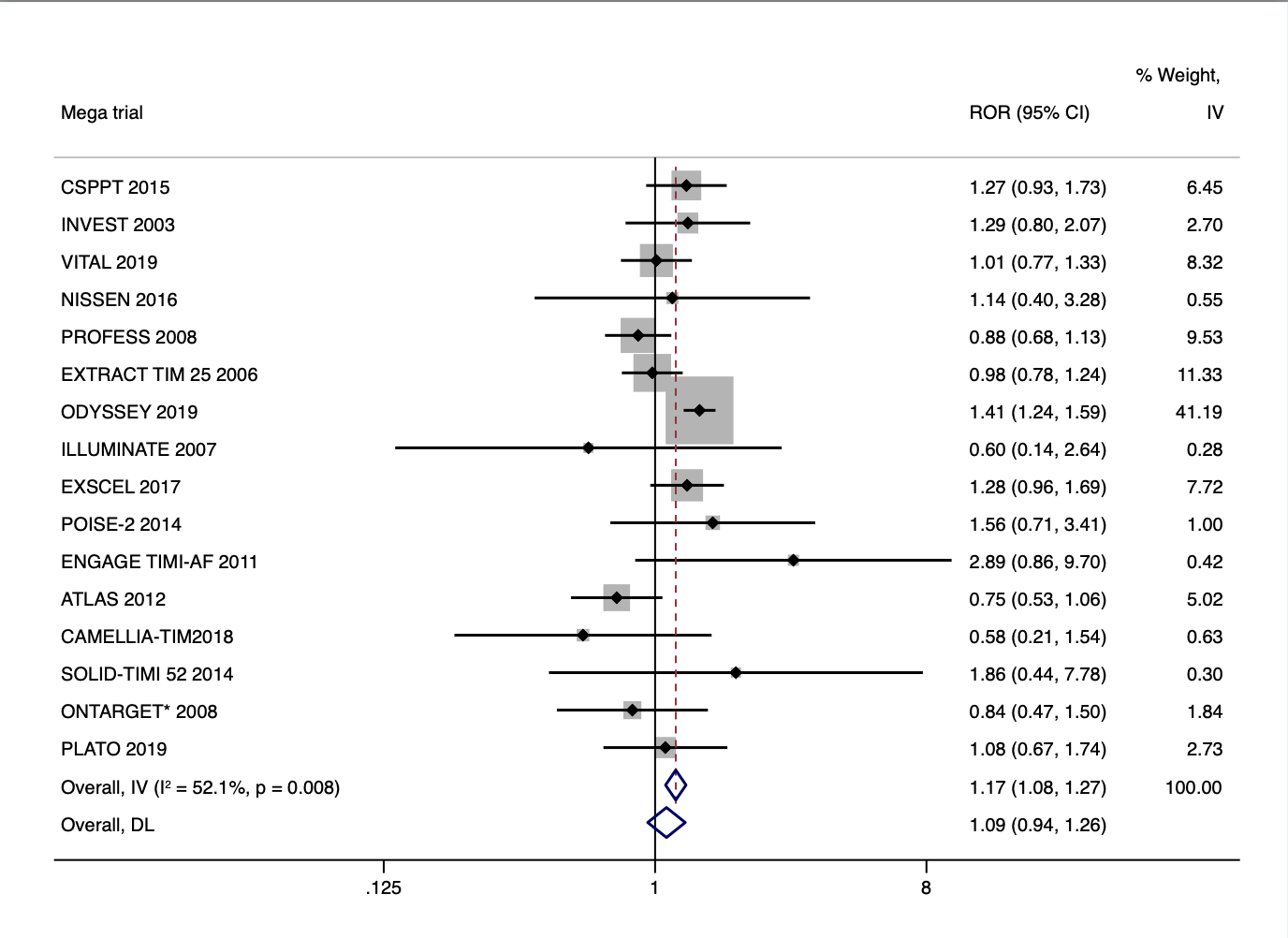

**Figure S2c.** Agreement between mega-trials and smaller trials trials with 1/5 of the least weighted mega-trial – All-cause mortality.

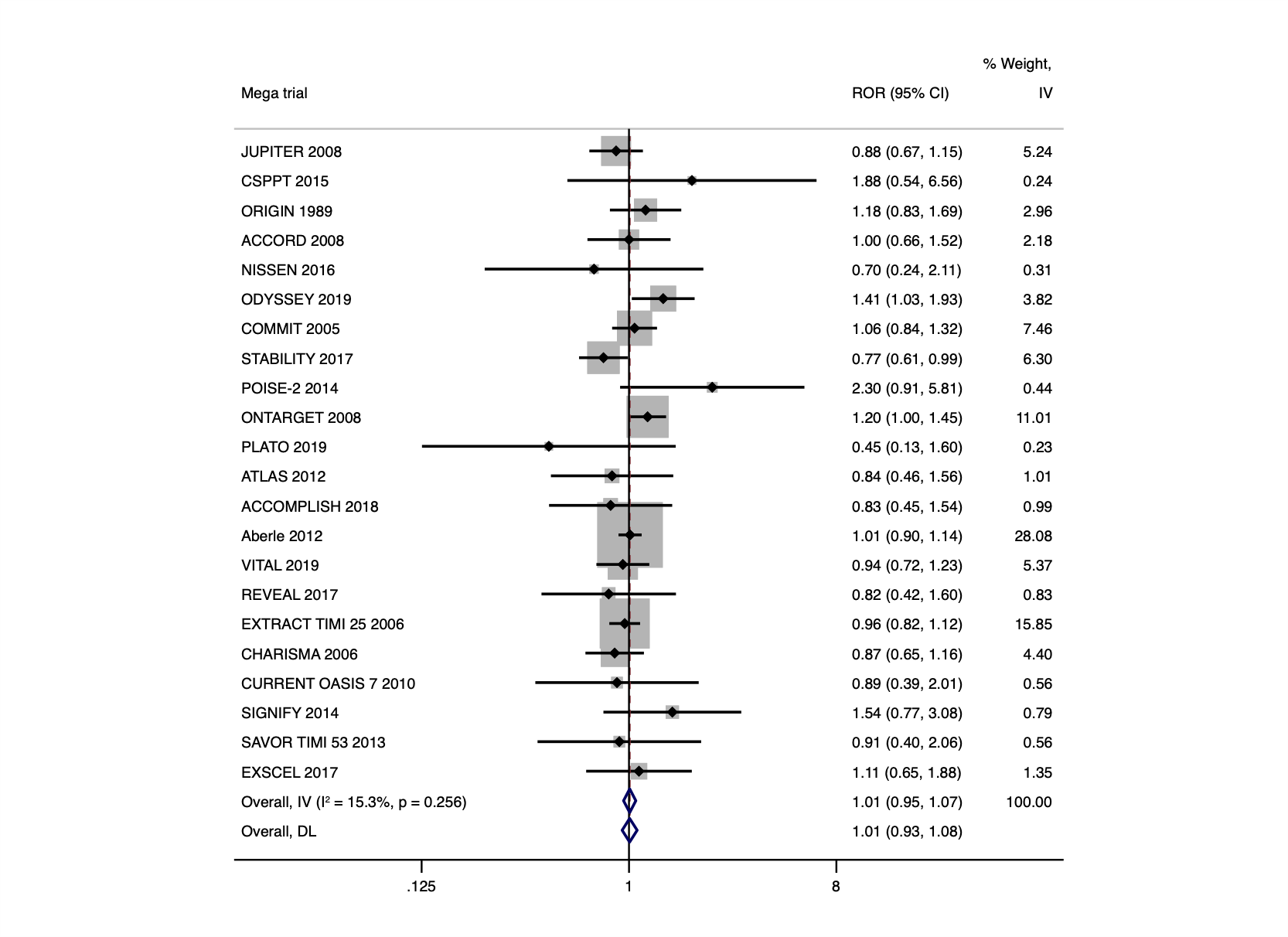

**Figure S2d.** Agreement between mega-trials and smaller trials trials with 1/5 of the least weighted mega-trial published prior or up until the mega-trial – All-cause mortality.

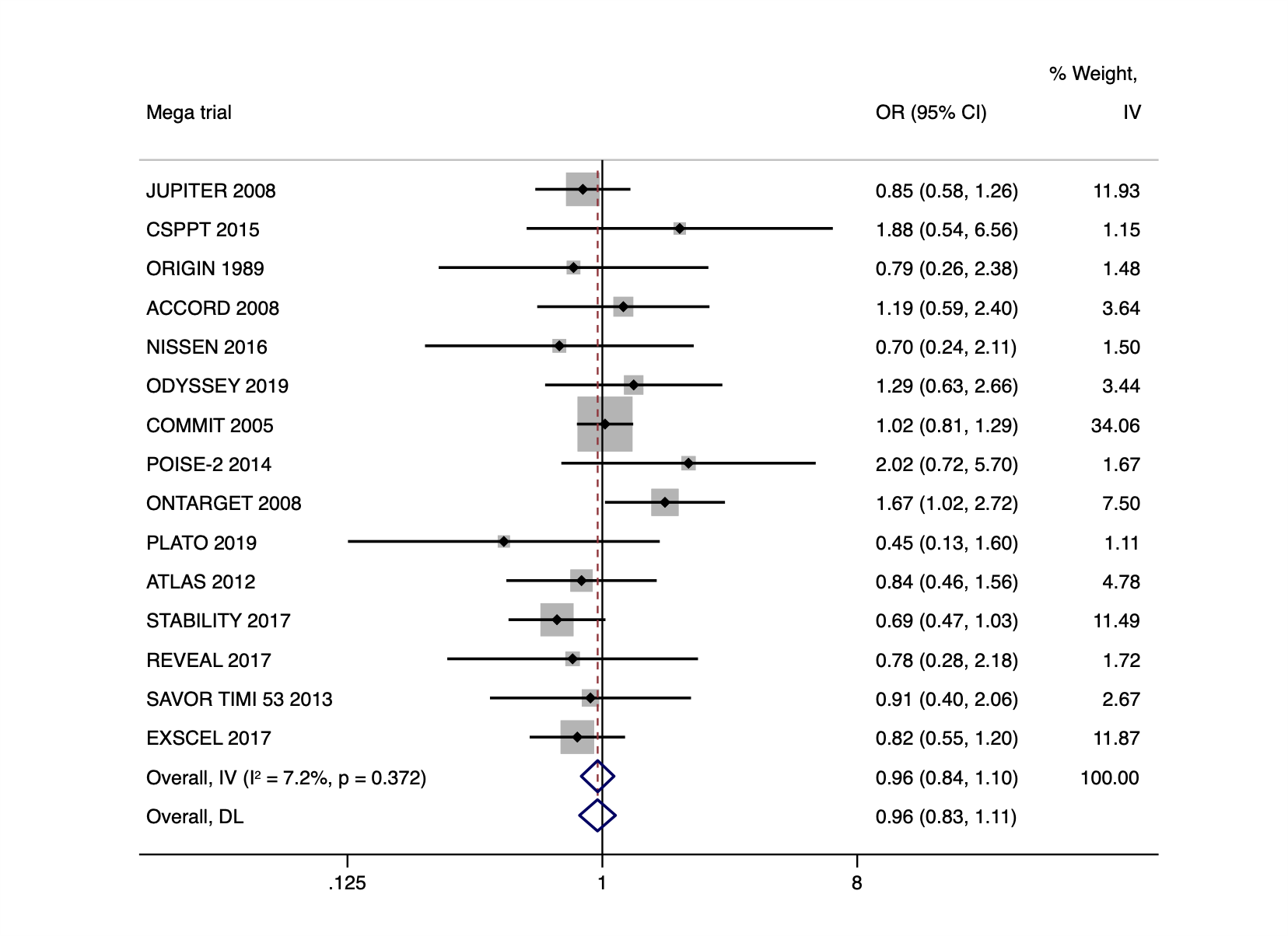

**Figure S3a**. Agreement between mega-trials and smaller trials with 1/10 of the least weighted megatrial – Primary outcome.

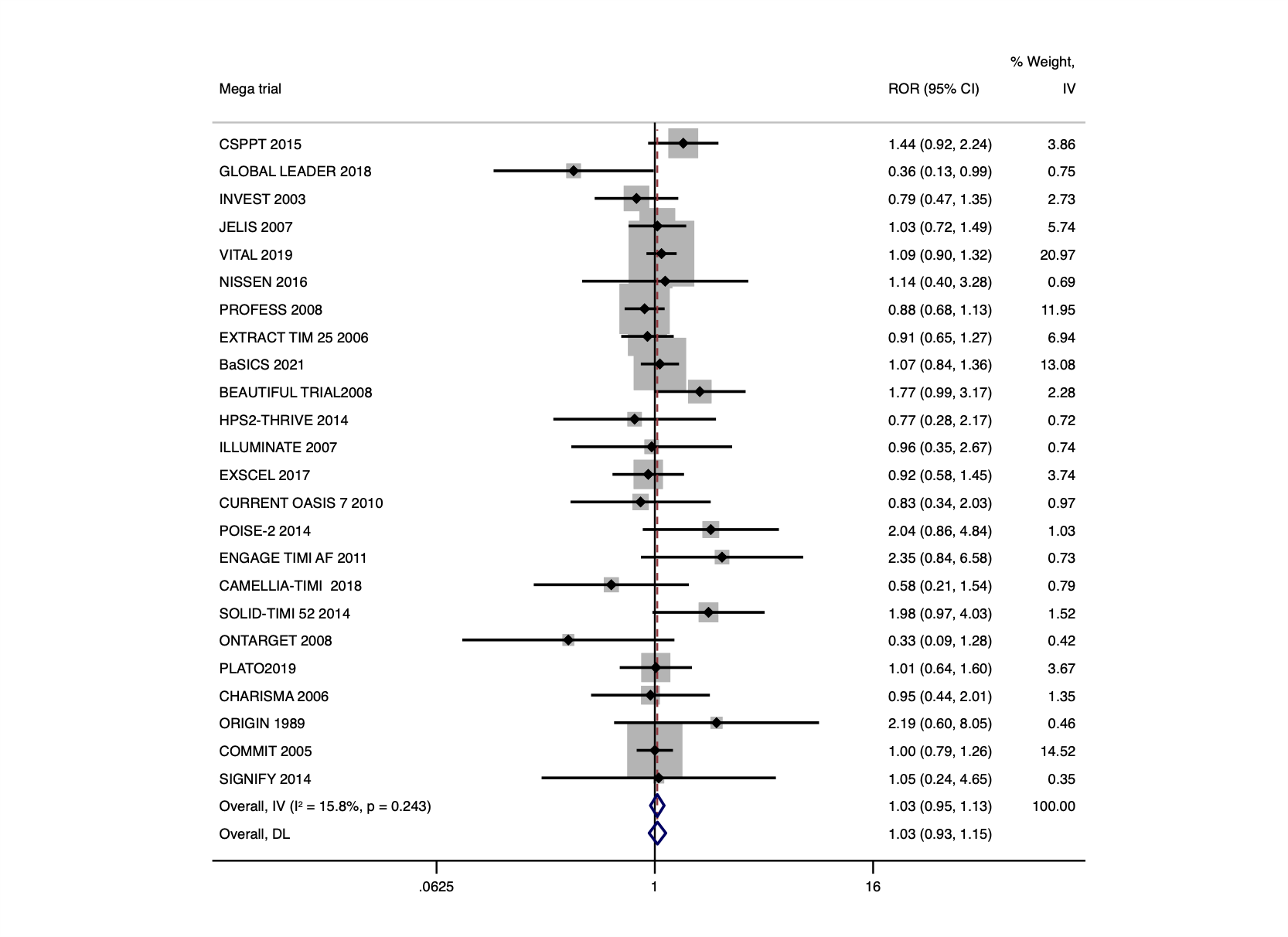

**Figure S3b**. Agreement between mega-trials and smaller trials with 1/10 of the least weighted mega-trial published prior or up until the mega-trial – Primary outcome.

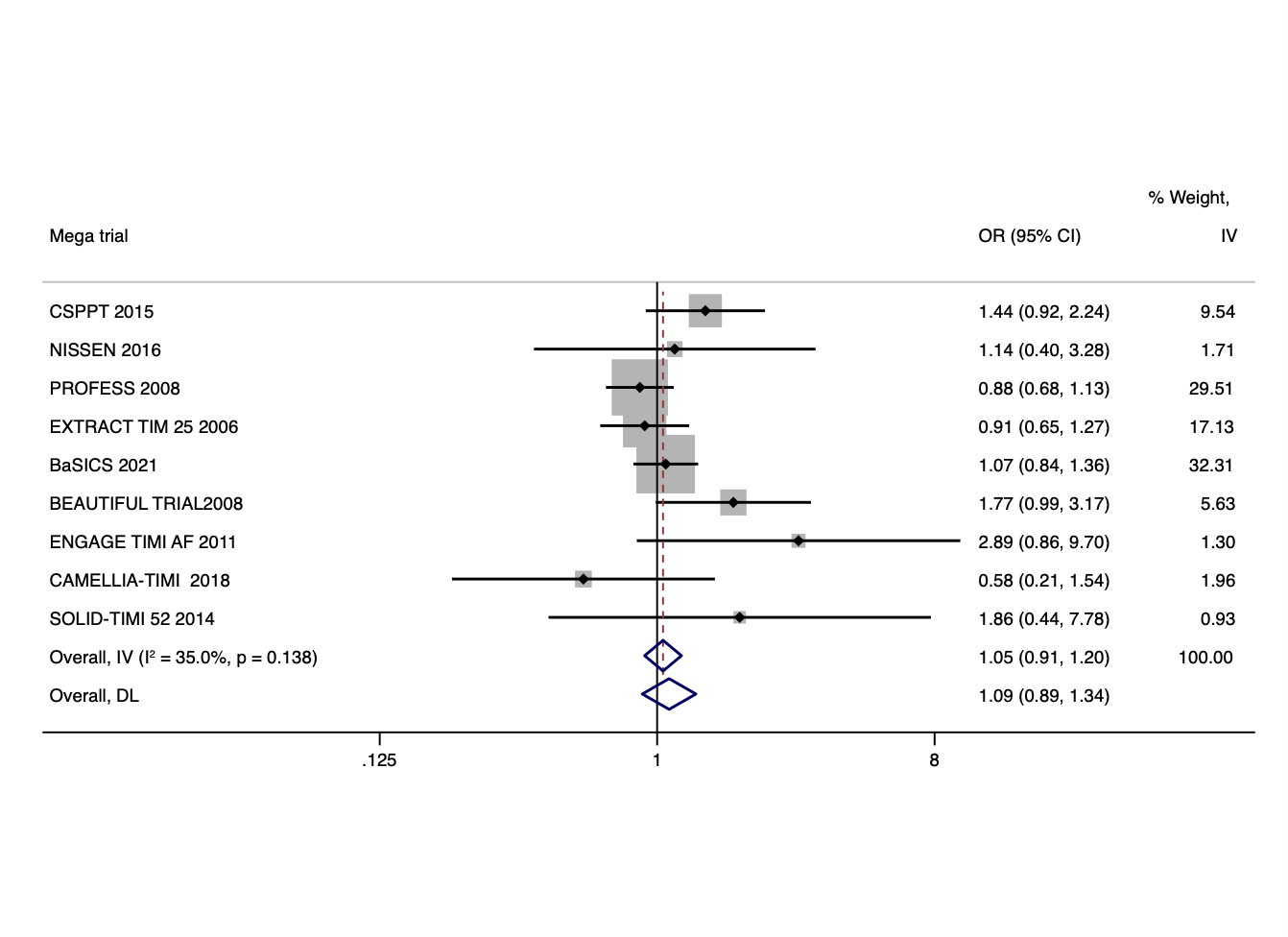

**Figure S3c.** Agreement between mega-trials and smaller trials with 1/10 of the least weighted megatrial – All-cause mortality.

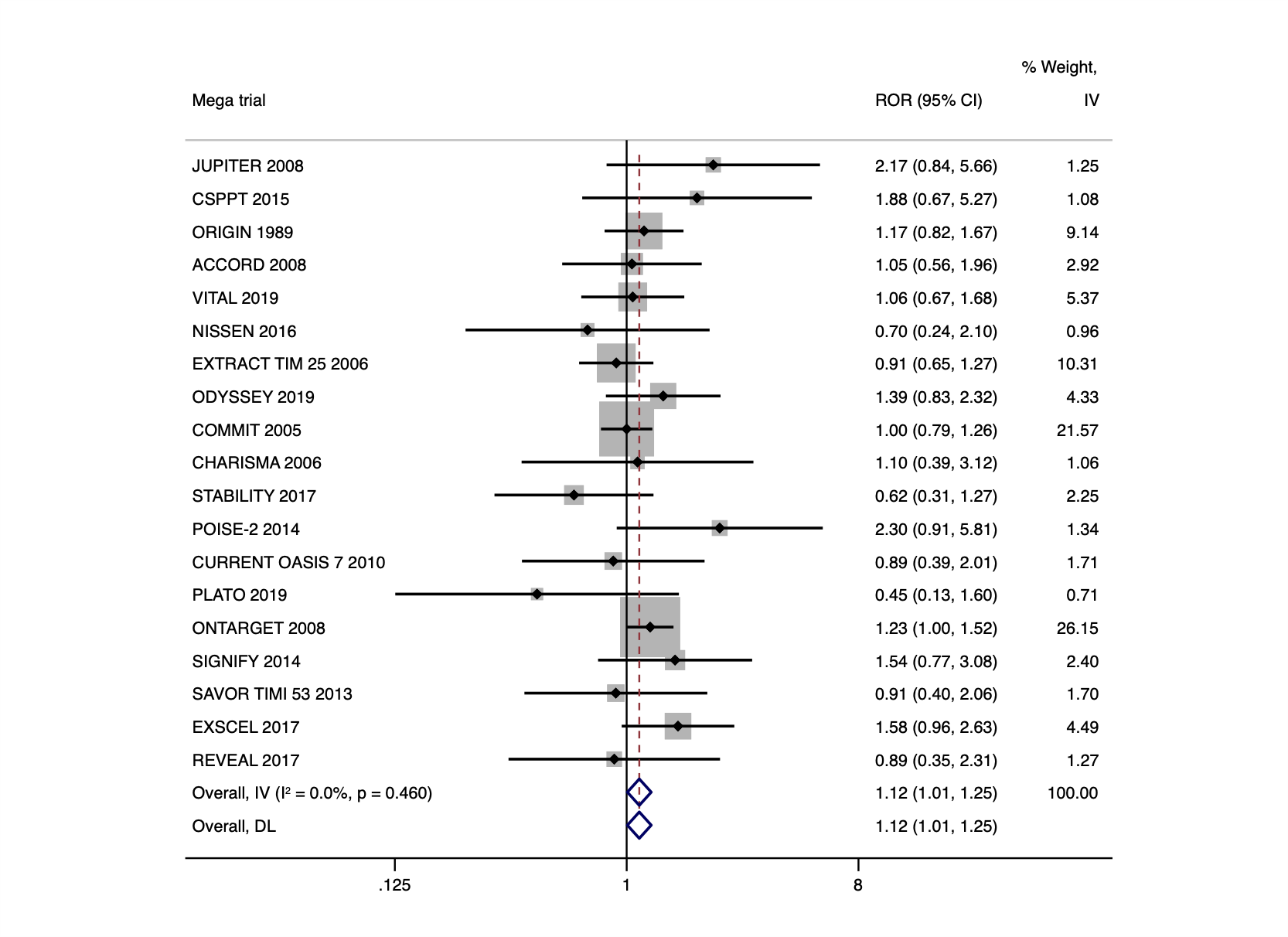

**Figure S3d.** Agreement between mega-trials and smaller trials with 1/10 of the least weighted mega-trial published prior or up until the mega-trial – Primary outcome.

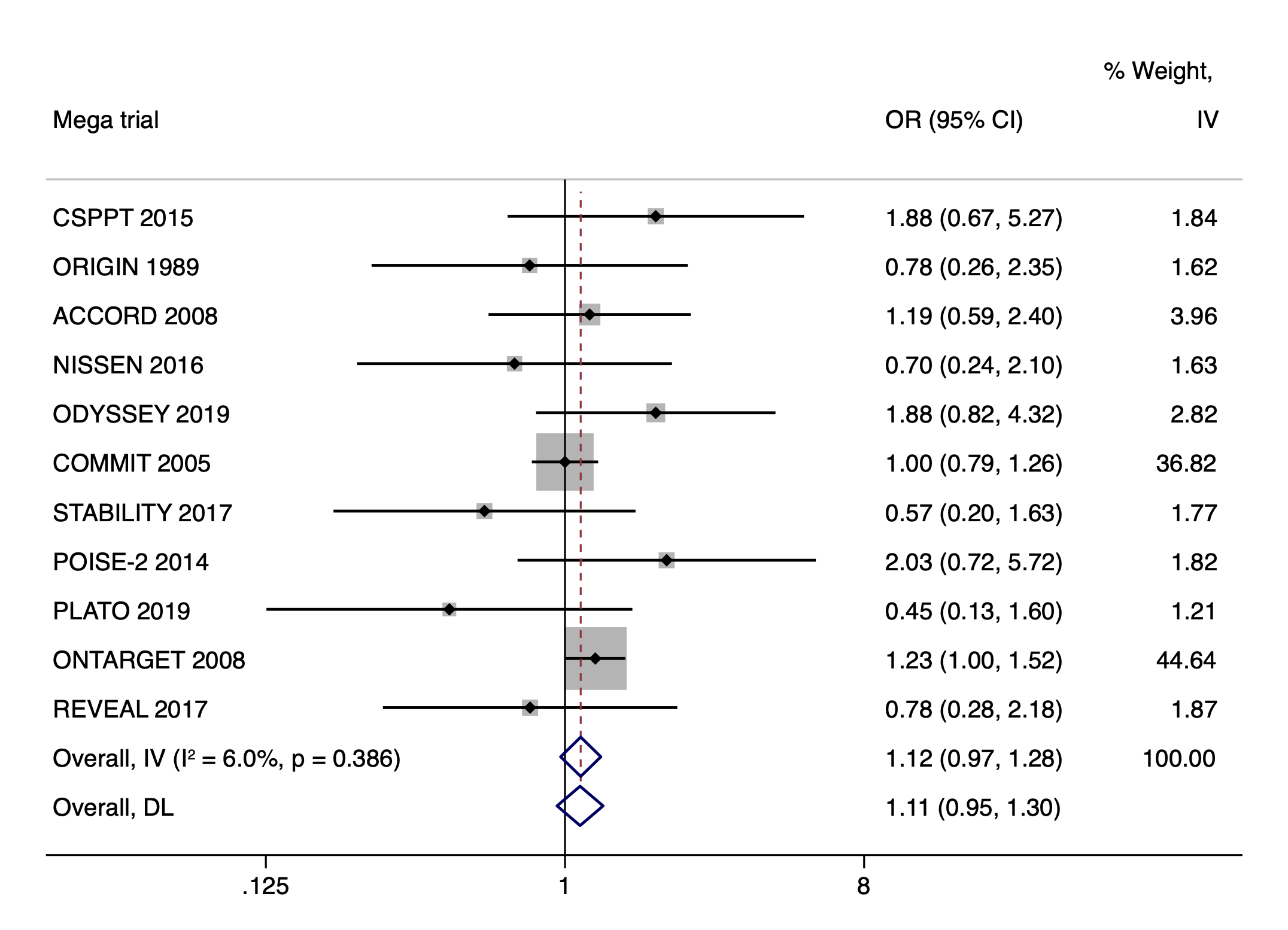

**Figure S4a**. Agreement between mega-trials and smaller trials on the primary outcome pooling the results using fixed effects.

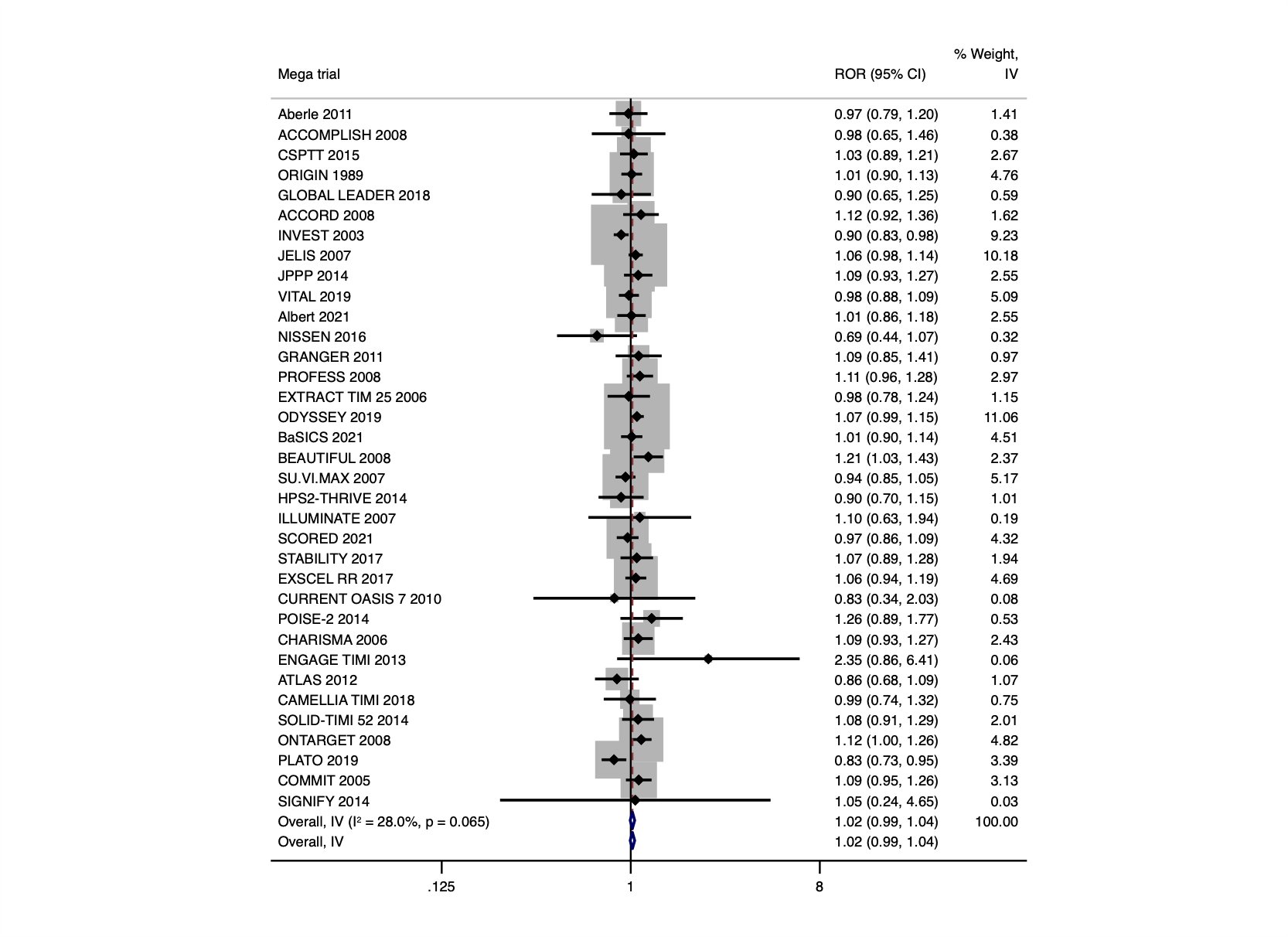

**Figure S4b.** Agreement between mega-trials and smaller trials on the all-cause mortality pooling the results using fixed effects.
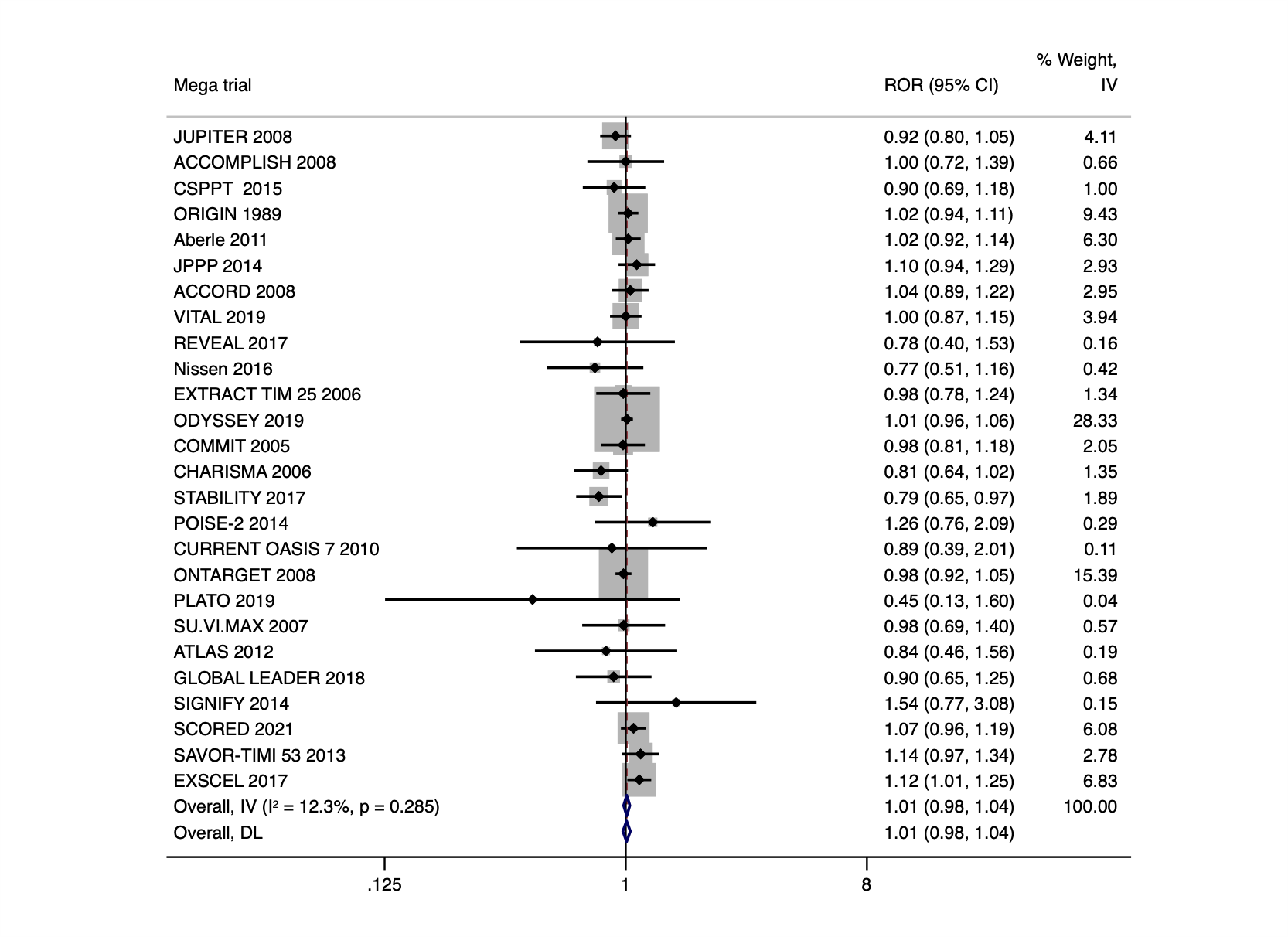

**Figure S5a.** Agreement between mega-trials and smaller trials stratified to blinding- Primary outcome.

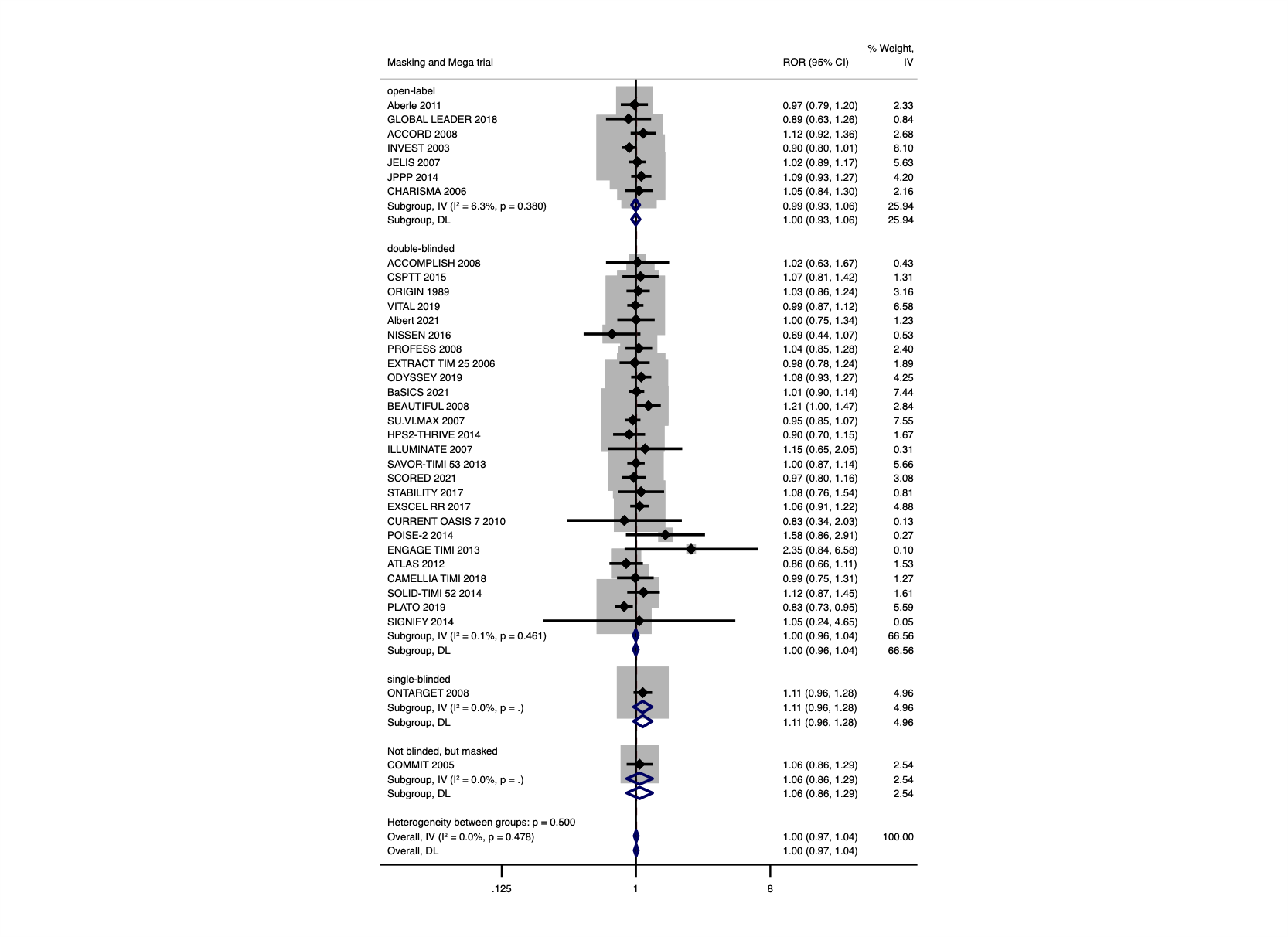

**Figure S5b.** Agreement between mega-trials and smaller trials stratified to blinding- All-cause mortality.

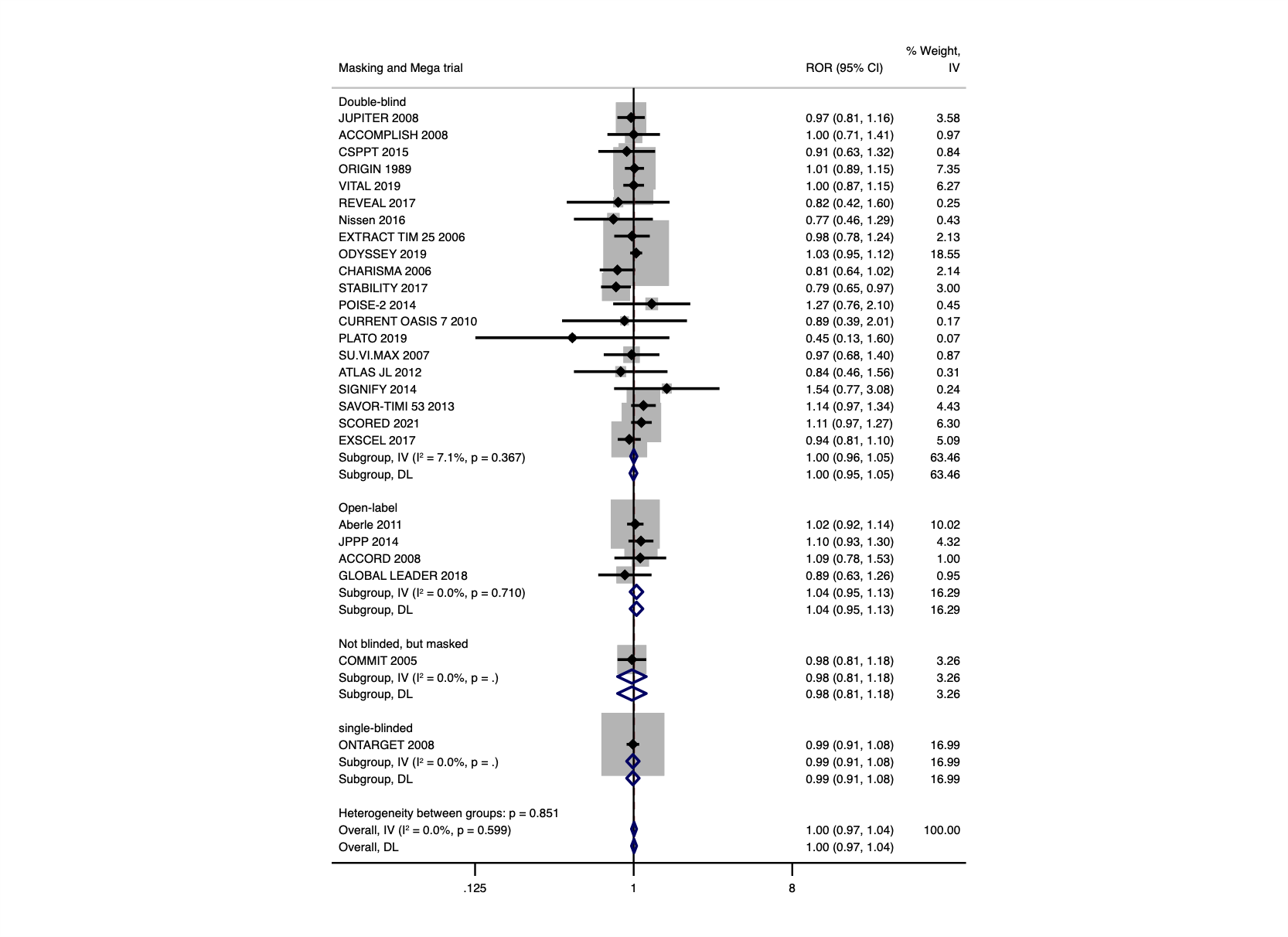

**Figure S6a.** Agreement between mega-trials and smaller trials stratified to intervention type- Primary outcome.
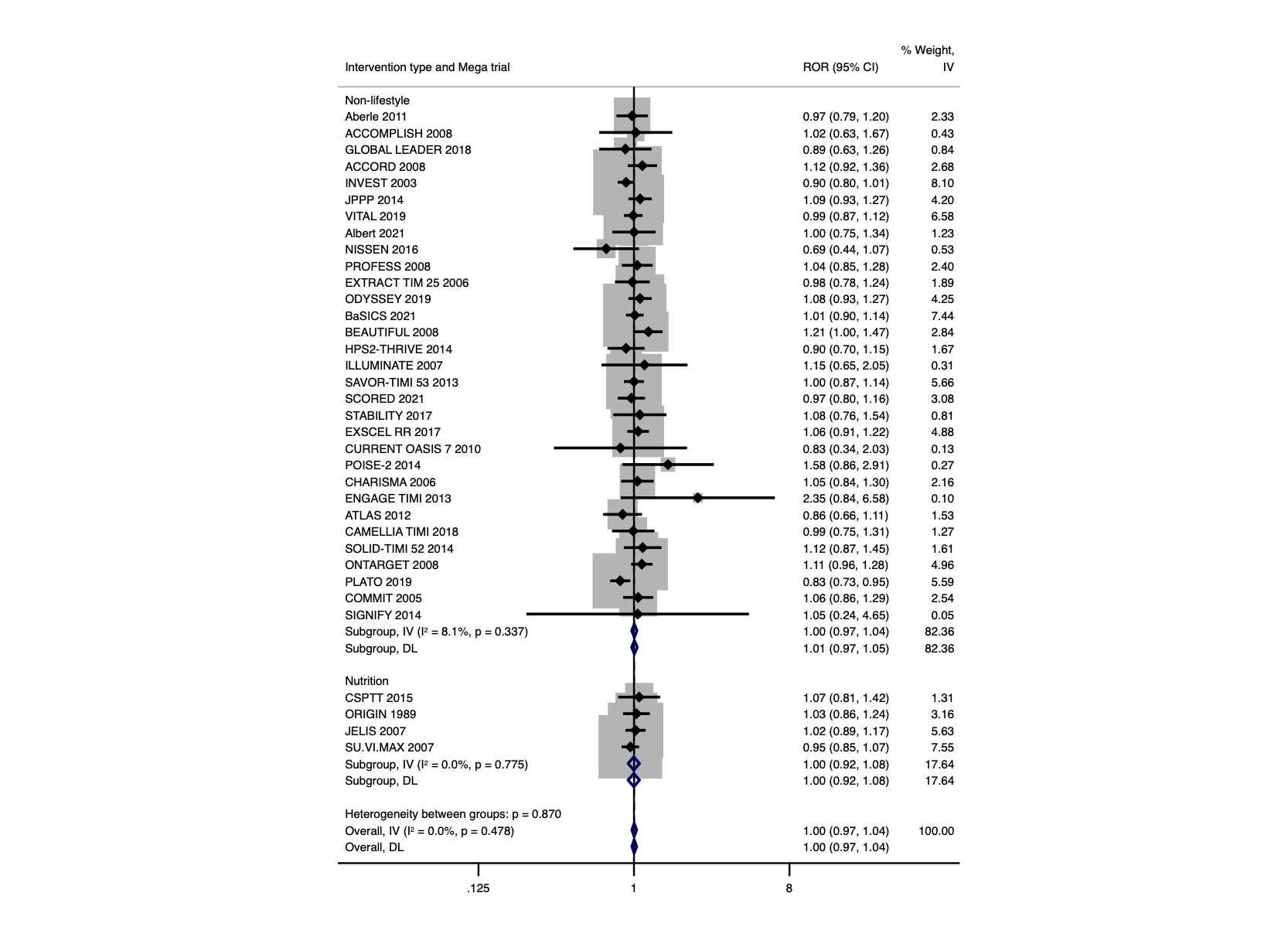

**Figure S6b.** Agreement between mega-trials and smaller trials stratified to intervention type- All-cause mortality.

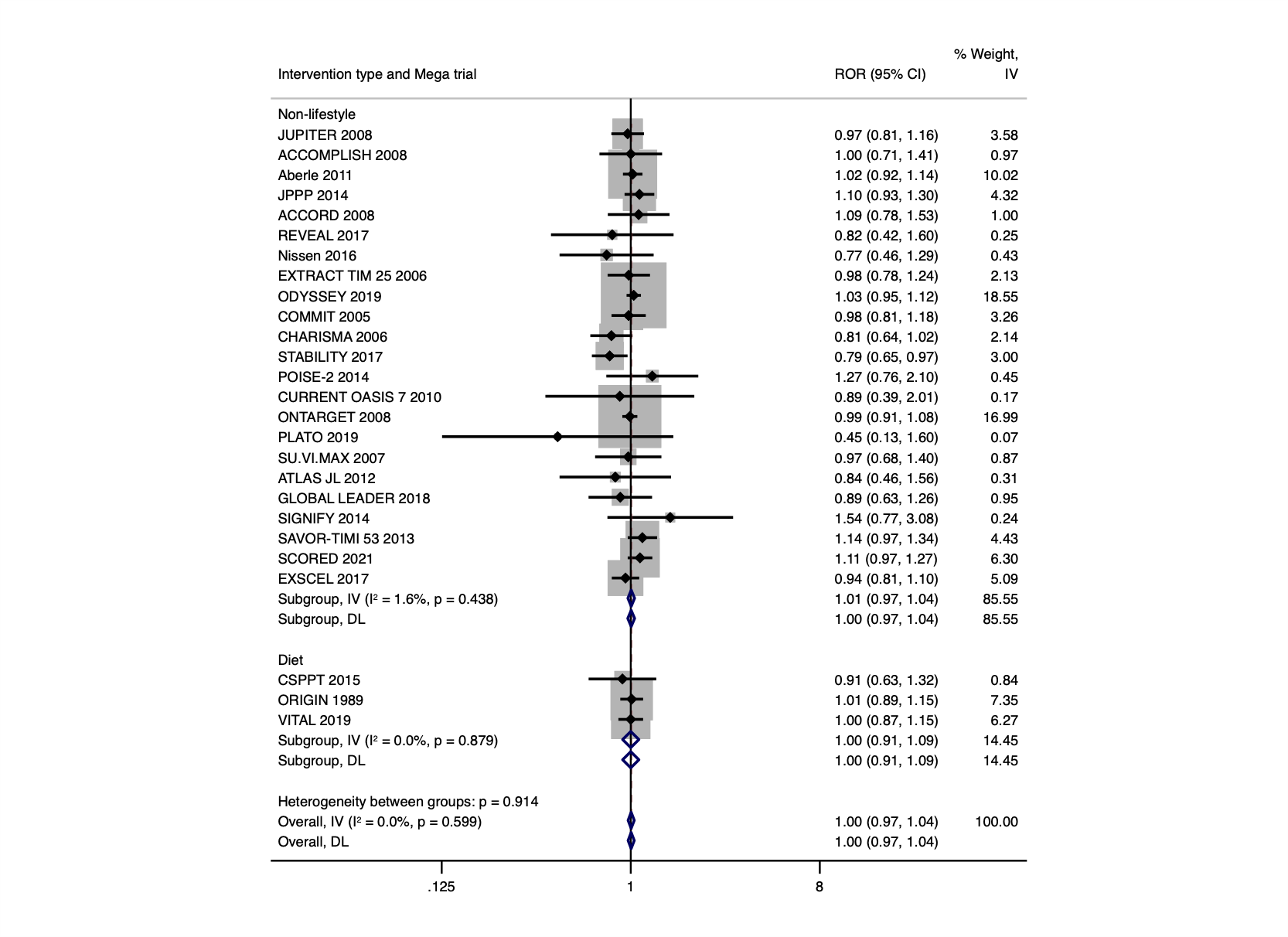

**Figure S7a.** Agreement between mega-trials and smaller trials stratified to specialty- Primary outcome.
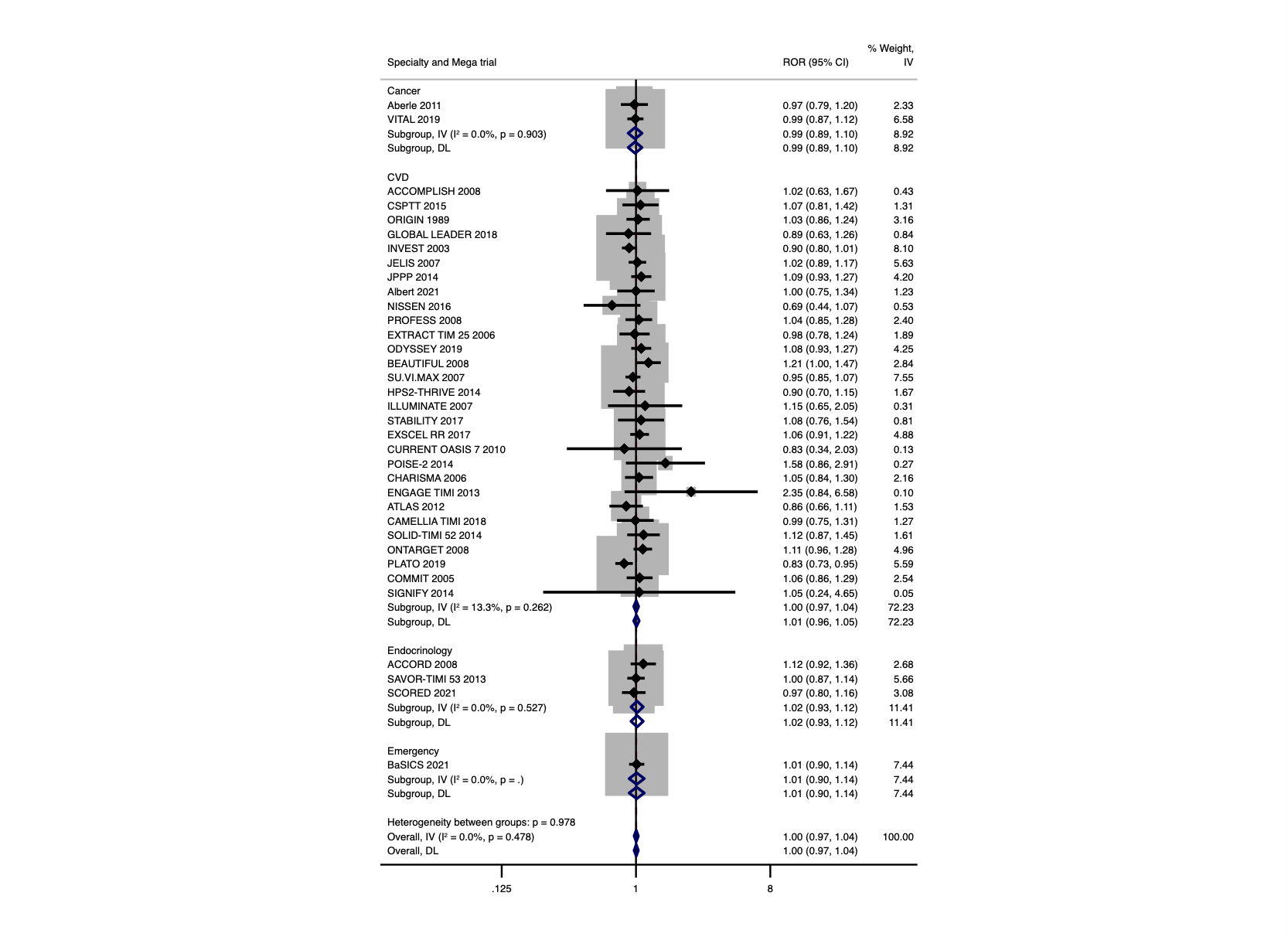

**Figure S7b.** Agreement between mega-trials and smaller trials stratified to specialty- All-cause mortality.

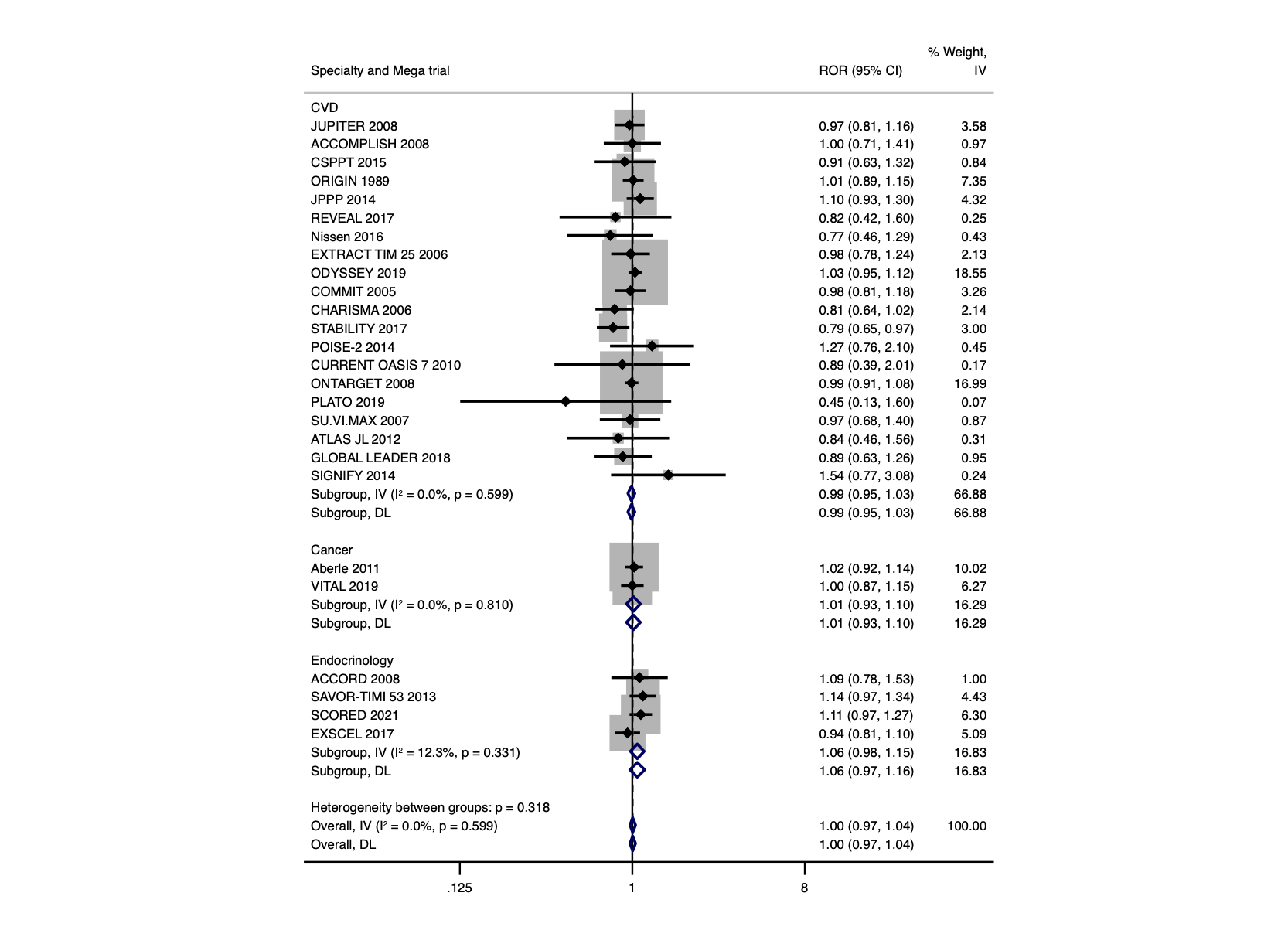

**Figure S8a.** Agreement between mega-trials and smaller trials stratified to heterogeneity- Primary outcome.

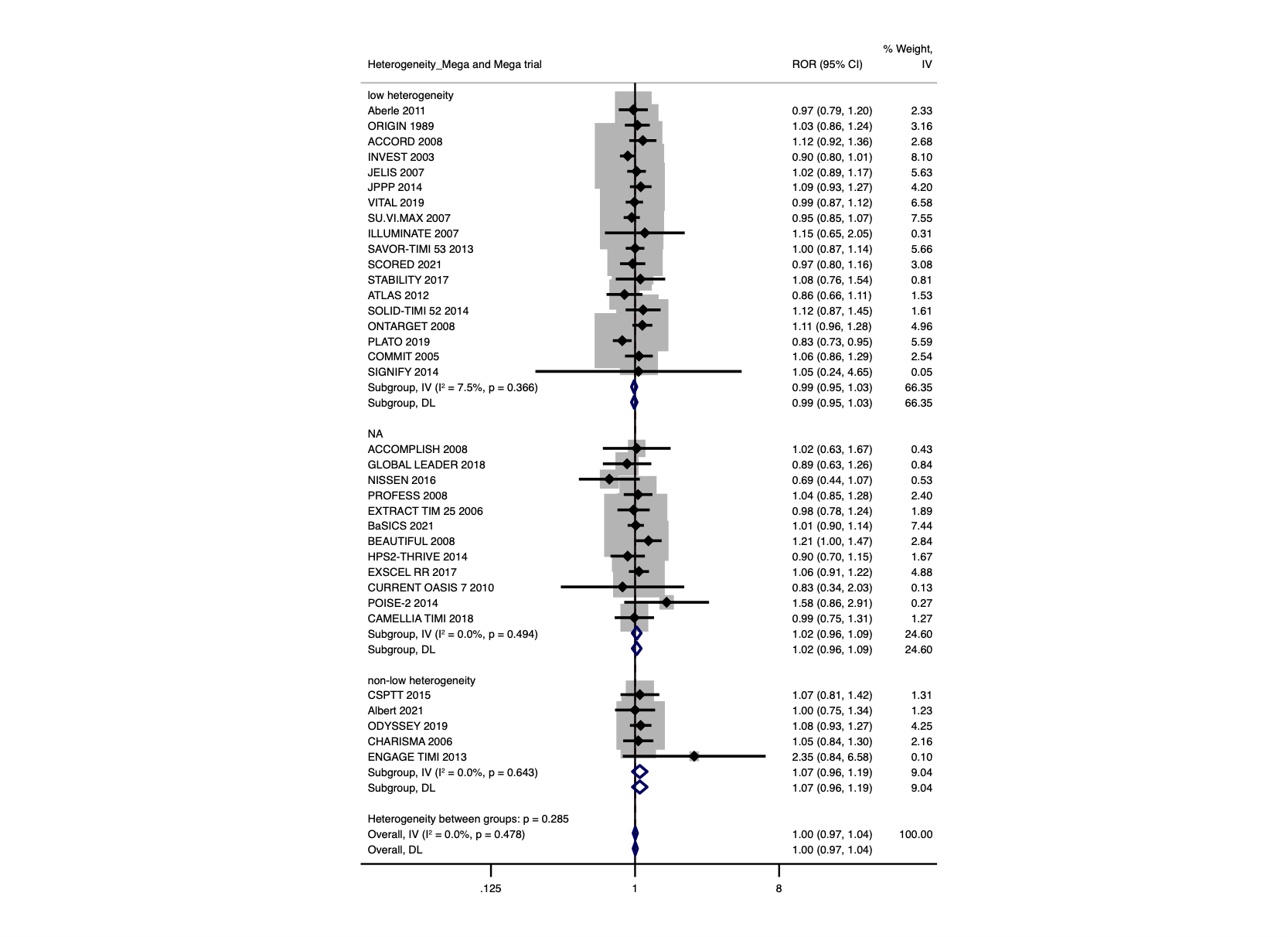

**Figure S8b.** Agreement between mega-trials and smaller trials stratified to heterogeneity - All-cause mortality.

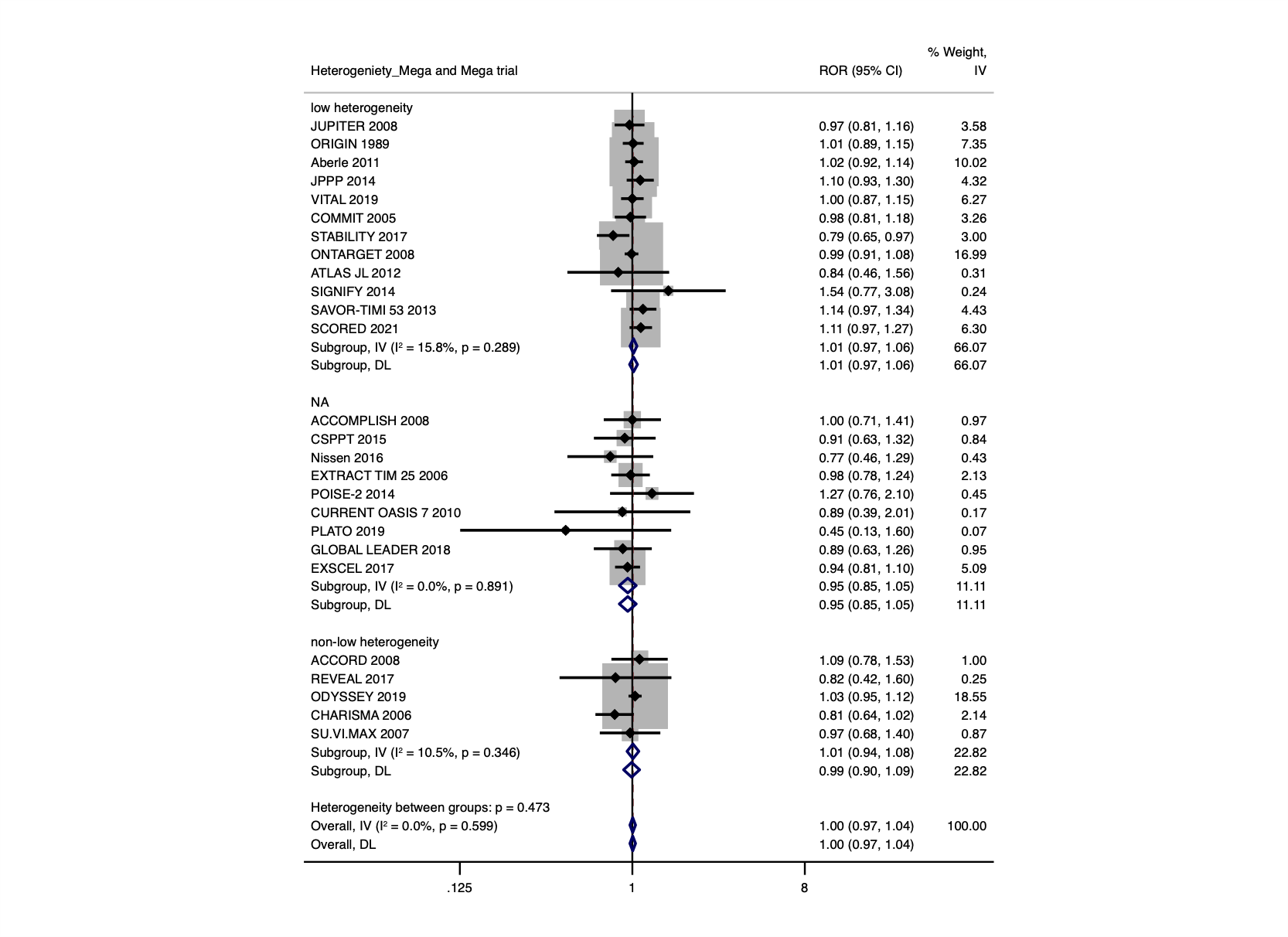
