## Supplementary Tables for "Agreement between mega-trials and smaller trials: a meta-research study"

| **MEGA-TRIAL** | **Primary outcome** | **Intervention** | **Control** | **Primary outcome**  **OR (95% CI)** |
| --- | --- | --- | --- | --- |
| **ACCOMPLISH 2008 [1]** | MACE | ACEi/ARBs+CCB | Other combinations | 0.8 (0.72-0.9) |
| **GLOBAL LEADER 2018 [2]** | All-cause mortality or new Q-wave myocardial infarction | Very short duration of antiplatelet therapy | >3 months antiplatelet therapy | 0.87 (0.75–1.01) |
| **NISSEN 2016 [3]** | Cardiovascular death | Celecoxib  Celecoxib | Naproxen  Ibuprofen | 0.9 (0.71-1.15)  0.81 (0.65-1.02) |
| **EXTRACT TIMI 2006 [4]** | Death or nonfatal recurrent myocardial infarction | Enoxaparin | Heparin | 0. 9 (0.8-1.01) |
| **BEAUTIFUL 2008 [5]** | MACE | Ivabradine | Placebo | 1.00 (0.91-1.10) |
| **HPS-2 THRIVE 2004 [6]** | MACE | Niacin-laropiprant | Placebo | 0.96 (0.9-1.03) |
| **ILLUMINATE 2007 [7]** | MACE | Torcetrapib+Atorvastatatin | Atorvastatin | 1.25 (1.09-1.44) |
| **PLATO 2009 [8]** | MACE | Ticagrelor | Clopidogrel | 0.85 (0.77-0.93) |
| **CURRENT OASIS 7 2010[9]** | MACE | Double dose clopidogrel | Standard dose | 0.85 (0.73-0.98) |
| **POISE-2 2014 [10]** | MACE | Clonidine | Placebo | 1.08 (0.93-1.26) |
| **ENGAGE TIMI AF [11]** | MACE | High dose Edoxaban  Low dose Edoxaban | Warfarin | 0.89 (0.83-0.96)  0.83 (0.77-0.9) |
| **SOLID TIMI 52 2014 [12]** | MACE | Darapladib | Placebo | 1.00 (0.91-1.09) |
| **ONTARGET 2008[13]** | MACE | Ramipril | Telmisartan | 1.01 (0.94-1.09) |
| **REVEAL 2017 [14]** | MACE | Anacetrapib | Placebo | 0.91 (0.85-0.97) |
| **ARRIVE 2018 [15]** | MACE | Aspirin | Placebo | 0.96 (0.81-1.13) |
| **ACCELERATE 2017 [16]** | MACE | Evacetrapib | Placebo | 1.01 (0.90-1.10) |
| **Dal-OUTCOME 2012 [17]** | MACE | Dalcetrapib | Placebo | 1.04 (0.92-1.16) |
| **TRITON TIMI 2007 [18]** | MACE | Prasugrel | Clopidogrel | 0.81 (0.73-0.9) |
| **CHAMPION PHOENIX 2016 [19]** | MACE | Cangrelor | Clopidogrel | 0.83 (0.67-1.01) |

**Table S1.** The composite primary outcome and effect estimates of mega-trials identified by our search but analyzed only for a subset of the primary outcome.

**MACE-** Major Adverse Cardiovascular Events,

| **Mega-Trial** | **Primary Outcome** | **Intervention** | **Control** | **Primary Outcome**  **OR (95% CI)** | **All-Cause Mortality**  **OR (95% CI)** | **Purpose of study** | **Risk of Bias** |
| --- | --- | --- | --- | --- | --- | --- | --- |
| **VALUE 2004 [20]** | MACE | Valsartan | Amlodipin | 1.04 (0.94-1.15) | 1.04 (0.94-1.14) |  | Low |
| **ATBC 2003 [21]** | Site-specific cancer incidence and total and cause-specific mortality and calendar time-specific risk for lung cancer incidence and total mortality. | Beta carotene | Placebo | Only one of sixteen outcomes had significant findings | Not significant  Multiple outcome estimates | - | Low |
| **Omenn 1996. [22]** | Incidence of lung cancer | Beta carotene and Vitamin A | Placebo | 1.28 (1.05-1.57) | 1.17 (1.03-1.33) | - | Low |
| **Hennekens 1996 [23]** | Malignant neoplasms and cardiovascular disease | Beta carotene | Placebo | 0.98 (0.91-1.06) | 1.01 (0.92-1.11) | - | Low |
| **Goodman 2004 [24]** | Lung cancer | Beta carotene | Placebo | 1.12 (0.97-1.31) | 1.08 (0.99-1.17) | - | Low |
| **ISIS-1 1986 [25]*** | Vascular Mortality | Atenolol | Control | 0.84 (0.72-0.99) | 0.85 (0.73-0.99) | - | High |
| **REAL CAD 2018 [26] *** | MACE | High dose Pitavastatin | Low dose Pitavastatin | 0.81 (0.69-0.95) | 0.81 (0.68-0.98) | - | High |
| **SEARCH 2010 [27]** | MACE | 80mg simvastatin | 20mg simvastatin | 0.94 (0.88-1.01) | 0.99 (0.91-1.09) | - | Low |
| **TNT 2005 [28]** | MACE | 80mg simvastatin | 10mg simvastatin | 0.78 (0.69-0.89) | 1.01 (0.85-1.19) | - | Low |
| **ASCOT LLA 2003 [29]*** | Non-fatal myocardial infarction and fatal CHD | Atorvastatin | Placebo | 0.64 (0.5-0.83) | 0.87 (0.71-1.06) | - | Low |
| **ALLHAT 2002 [30]*** | Fatal CHD or nonfatal myocardial infarction | Amlodipine  Lisinopril | Chlorthalidone | 0.98 (0.9-1.07)  0.99 (0.91-1.08) | 0.96 (0.89-1.02)  1.00 (0.94-1.08) | - | High |
| **Lacroix 2009 [31]** | Mortality | Calcium + Vitamine D | Placebo | 0.91 (0.83-1.01) | 0.91 (0.83-1.01) | - | Low |
| **HOT 1998 [32]** | MACE | Aspirin | Placebo | 0.85 (0.73-0.99) | 0.93 (0.79-1.09) | - | Low |
| **DeKoning 2020 [33]*** | Lung Cancer Mortality | Screening | Control | 0.74 (0.-61-0.90) | 0.98 (0.89-1.08) | - | High |
| **GISSI-P 1999 [34]*** | MACE | n-3 PUFA  vitamin E  both | Placebo | 0.90 (0.82–0.99)  0.95 (0.86–1.05)  0.86 (0.74–0.99) | 0.86 (0.76–0.97)  0.92 (0.82–1.04) | - | Low |
| **Leppälä 2000 [35]** | Stroke mortality | α-Tocopherol β-Carotene  both | Placebo | Non-significant | NA | - | Low |
| **Lee 1999 [36]** | Cancer and cardiovascular mortality | β-Carotene | Placebo | 1.03 (0.89-1.18)  1.14 (0.87-1.49) | 1.07 (0.74-1.56) | - | High |
| **ASPREE 2018 [37]** | Death, dementia, or persistent physical disability | Aspirin | Placebo | 1.01 (0.98-1.11) | 1.14 (1.01-1.29) | - | Low |
| **WHS 2005 [38]** | MACE | Aspirin | Placebo | 0.91 (0.8-1.03) | 0.95 (0.85-1.06) | - | Low |
| **CAPPP 1999 [39]*** | MACE | Captopril | Conventional antihypertensives | 1.05 (0.9-1.22) | 0.93 (0.76-1.14) | - | Low |
| **ASCOT-BPLA 2005 [40]*** | MACE | Amlodipine | Atenolol | 0.9 (0.79-1.02) | 0.89 (0.81-0.99) | - | Low |
| **CONVINCE 2003 [41]** | MACE | Verapamil | Atenolol or Hydrochlorthiazide | 1.02 (0.88-1.18) | 1.08 (0.93-1.26) | - | Low |
| **NORDIL 2000 [42]*** | MACE | Diltiazem | Diuretic and Beta-blockers | 1.00 (0.87-1.15) | 1.00 (0.83-1.20) | - | Low |
| **ADVANCE 2008 [43]*** | MACE and Major microvascular events | Intensive glucose control | Standard control | 0.88 (0.8-0.97) | 0.92 (0.81-1.05) | - | High |
| **WHI 2006 [44]** | Colorectal Cancer | Calcium | Placebo | 1.08 (0.86-1.34) | 0.91 (0.83-1.01) | - | Low |
| **IMPROVE IT 2015 [45]** | MACE | Ezetimibe + Statin | Statin | 0.94 (0.89-0.99) | 0.99 (0.91-1.07) | - | Low |
| **FOURIER 2017 [46]** | MACE | Evolocumab | Placebo | 0.85 (0.79-0.92) | 1.04 (0.91-1.19) | - | Low |
| **TECOS 2015 [47]** | MACE | Sitagliptin | Placebo | 0.98 (0.88-1.09) | 1.01 (0.9-1.14) | Noninferiorty | Low |
| **STRENGTH 2020 [48]** | MACE | Omega-3 | Corn oil | 0.99 (0.9-1.09) | 1.13 (0.97-1.42) | - | Low |
| **MRC/BHF 2002[49]** | MACE | Vitamins | Placebo | 0.99 (0.93-1.06) | 1.03 (0.92-1.17) | - | Low |
| **MRC/BHF 2002[50]** | All-cause Mortality | Simvastatin | Placebo | 0.87 (0.79-0.93) | 0.87 (0.79-0.93) | - | Low |

**Table S2.** Characteristics of the additional identified mega-trials that have not been identified by our search.

**OR**– Odds Ratio, **CI**- Confidence Intervals, **ACEi-** Angiotensin Converting Enzyme inhibitors ,**ARBs-** Angiotensin Receptor Blockers

**MACE-** Major Adverse Cardiovascular Events, **PUFA-** Polyunsaturated Fatty Acids

Trials denoted with * had an open-label design

|  | Primary Outcome |  | All-Cause Mortality |  |
| --- | --- | --- | --- | --- |
|  | Coefficient (SE) | P-value | Coefficient ( SE ) | P-value |
| Intervention | -0.01(0.08) | 0.89 | .009 (0.1) | 0.9 |
| Speciality | 0.01(0.05) | 0.757 | 0.02 (0.06) | 0.688 |
| ROB-MEGA | 0.01 (0.07) | 0.87 | 0.01 (0.05) | 0.76 |
| ROB-SMALL | 0.04 (0.05) | 0.307 | 0.01 (0.05) | 0.854 |
| Median_small | -3.93e-06 ( 0.00001) | 0.77 | -1.42e (0.00002) | 0.95 |
| Total-small | 1.28e-06 (1.74e-06) | 0.46 | 1.44e-07 (7.03e-07) | 0.838 |
| Multivariate analyses |  | Non-significant |  | Non-significant |

**Table S3.** Results of uni and multivariable meta regression.

**SE-** Standard Error

**ROB-MEGA-** Proportion of mega-trials at high risk of bias.

**ROB-SMALL-** Proportion of smaller trials at high risk of bias.

**Median-small-** The median number of participants on the smaller trials.

**Total-small-** The total number of participants in the smaller trials.

**Multivariate Analysis-** Meta regression including all of the above listed variables.

| **MEGA-TRIAL** | Primary outcome | Intervention | Control | Primary Outcome | All-Cause Mortality | NonInferiority |
| --- | --- | --- | --- | --- | --- | --- |
| **COSMOS 2022 [51]** | MACE | Cocoa Extract | Placebo | Non-Significiant | Non-Significant | - |
| **NAITRE 2022 [52]** | All-cause Mortaility | Azithromycin | Placebo | Non-Significiant | Non-Significiant | - |
| **ARISTOTLE 2011 [53]** | Stroke or systemic embolism | Apixaban | Warfarin | Significant | Significant | Noninferiority |
| **THEMIS-PCI 2019 [54]** | MACE | Ticargrelor | Placebo | Significiant | Non-Significiant | - |
| **RE-LY 2010 [55]** | MACE | Dabigatran | Warfarin | Non-significant | Non-Significant | Noninferiority |
| **SOCRATES 2016 [56]** | MACE | Ticargrelor | Placebo | Non-significant | Non-significant | - |
| **TAO 2013 [57]** | All-cause mortality or Myocardial Infarction | Otamixaban | Unfractioned Heparin + Eptifibatide | Non-significant | Non-significant | -Superiority |
| **THALES 2020 [58]** | Stroke or Death | Ticargrelor-Aspirin | Aspirin | Significant | Non-significant | Superiority |
| **COPD 2018 [59]** | Rate of severe COPD | Triple Therapy | Duo-Therpy | Significant | Significant | - |
| **TIOSPIR 2013 [60]** | Death and COPD | Tiotropium Respimat | Tiotropium HandiHaler | Non-significant | Non-significant | Noninferiority |
| **NISSEN 2023 [61]** | MACE | Bempedoic Acid | Placebo | Significant | Significant | - |
| **AUSTRI 2016 [62]** | Serious asthma-related event | Fluticasone-Salmeterol | Flucitasone Alone | Non-Significant | Non-Significant | Noninferiority |
| **SUMMIT 2016 [63]** | All-cause mortality | Fluticasone furate  Vilaterol  Combination | Placebo | Non-Significant | Non-Significant | - |
| **MARINER 2018 [64]** | Composite of any symptomatic VTE | Rivaroxaban | Placebo | Non-Significant | Non-Significant | Superiority |
| **STAR 2006 [65]** | Invasive breast cancer | Tamoxifen | Raloxifene | Non-Significant | Non-Significant | - |
| **EUCLID 2017 [66]** | MACE | Ticargrelor | Clopidogrel | Non-Significant | Non-Significant | Superiority |
| **Peters 2016 [67]** | Serious asthma-related event | Budesonide + Formoterol | Budesonide | Non-Significant | Non-Significant | Non-inferiority |
| **SELECT 2009 [68]** | Prostate cancer | Selenium /+ Vitamin E | Placebo | Non-Significant | Non-Significant | - |
| **RUTH 2006 [69]** | Coronary events and invasive breast cancer | Raloxifene | Placebo | Non-Significant | Non-Significant | - |
| **Christen 2015 [70]** | Incident Cataract | Selenium | Vitamin E | Non-Significant | NA | - |
| **SCOUT 2010 [71]** | MACE | Sibutramine | Placebo | Non-Significant | Non-Significant | - |
| **WOMAN 2017 [72]** | All-cause mortality or hysterectomy | Tranexamic acid | Placebo | Non-Significant | Non-Significant | - |
| **Chandramohan 2019 [73]** | Death or admission not due to trauma | Malaria chemoprevention + Azithromycin | Malaria chemoprevention + Placebo | Non-Significant | Non-Significant | - |
| **CRASH-3 2019 [74]** | Head injury related death | Tranexamic acid | Placebo | Non-Significant | Non-Significant | - |
| **CRASH-2 2010 [75]** | Death in hospital within 4 weeks of injury | Tranexamic acid | Placebo | Significant | Significant | - |
| **HEAT 2015 [76]** | Time to hospitalization or death due to peptic ulcer bleeding | Clarithromycin, metronidazole and lansoprazole | Placebo | Non-Significant | Non-Significant | - |
| **HALT-IT 2020 [77]** | Death due to bleeding | Tranexamic Acid | Placebo | Non-Significant | Non-Significant | - |
| **BRUNVOLL 2022 [78]** | 4 coprimary outcomes related to respiratory infections | Cod liver oil | Placebo | Non-Significant | NA | - |
| **CRESCENDO 2010 [79]** | MACE | Rimonabant | Placebo | Non-Significant | Non-Significant | - |
| **ROC 2011[80]** | Survival to discharge | Early analysis of cardiac rhythm | Late analysis of cardiac rhythm | Non-Significant | NA | - |
| **PROMINENT 2022 [81]** | MACE | Pemafibrate | Placebo | Non-Significant | Non-Significant | - |
| **Sazawal 2016 [82]** | Mortality | Chlorhexidine | Dry cord care | Non-Significant | Non-Significant | - |
| **Ishani 2022 [83]** | MACE | Chlorthalidone | Hydrochlorthiazide | Non-Significant | Non-Significant | Superiority |
| **ZODIAC 2011 [84]** | Non-suicide mortality | Ziprasidone | Olanzapine | Non-Significant | Non-Significant | - |
| **NORDIC 2016 [85]** | Participation in colonoscopy screening, cancer and adenoma yield, and participant experience | Colonoscopy | No Screening | Not Clear | NA | - |
| **Hygia 2020 [86]** | MACE | Antihypertensive drugs in the morning | Antihypertensive drugs in the night | Significant | Significant | - |
| **Hansen 2012 [87]** | Difference in physical activity | Web-based physical activity promotion | No intervention | Significant | NA | - |
| **ROCKET AF 2011 [88]** | Stroke or Systemic  Embolism | Rivaroxamban | Warfarin | Significant | Non-Significant | Noninferiority |

**Table S4**. Characteristics of mega-trials identified by our search but had no eligible meta-analysis.

**MACE-** Major Adverse Cardiovascular Events

**COPD-** Chronic Obstructive Pulmonary Disease
