## Supplementary Material 1 for "Agreement between mega-trials and smaller trials: a meta-research study"

1. **Aberle 2011**

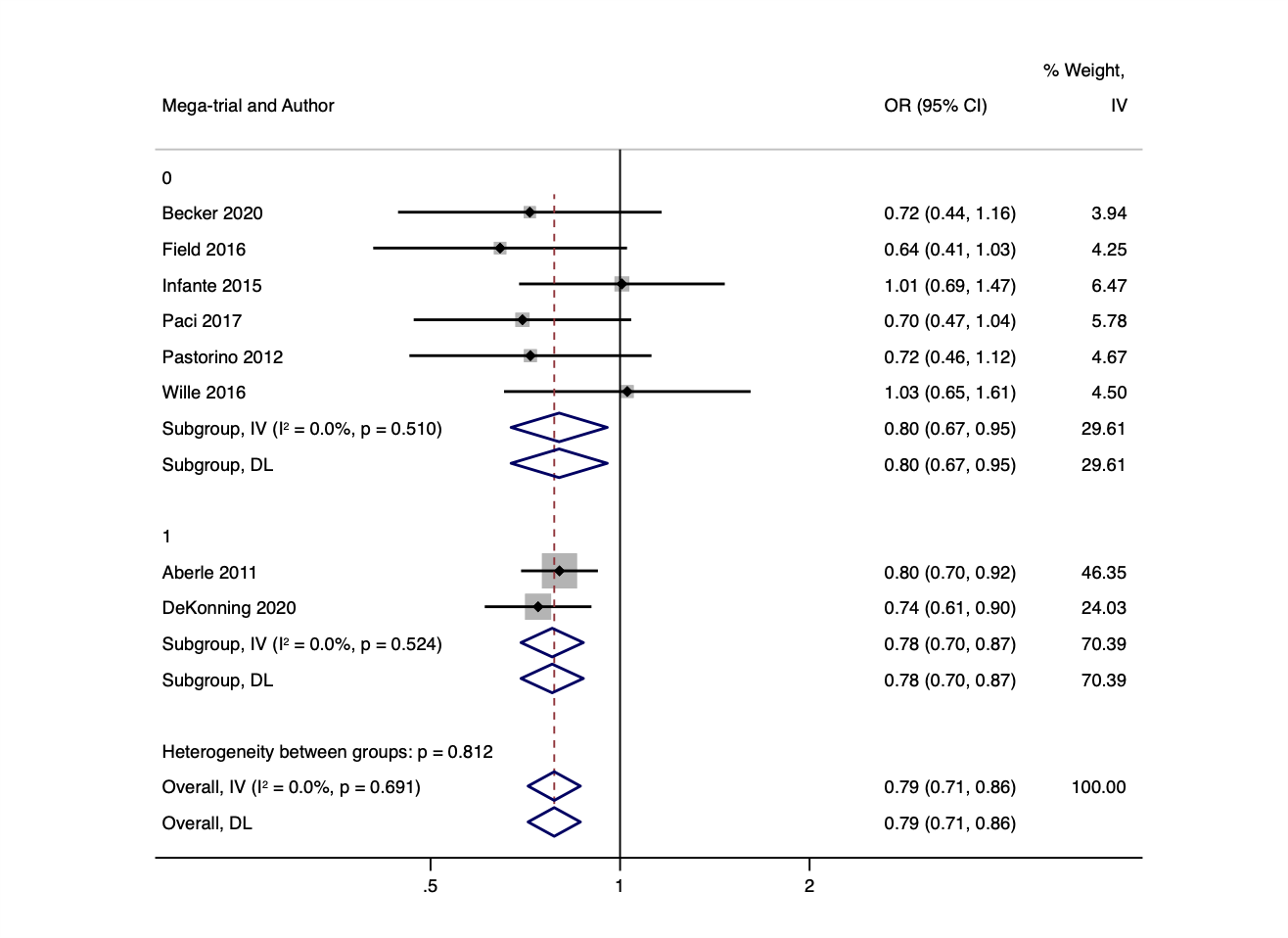

2. **ACCOMPLISH 2008**

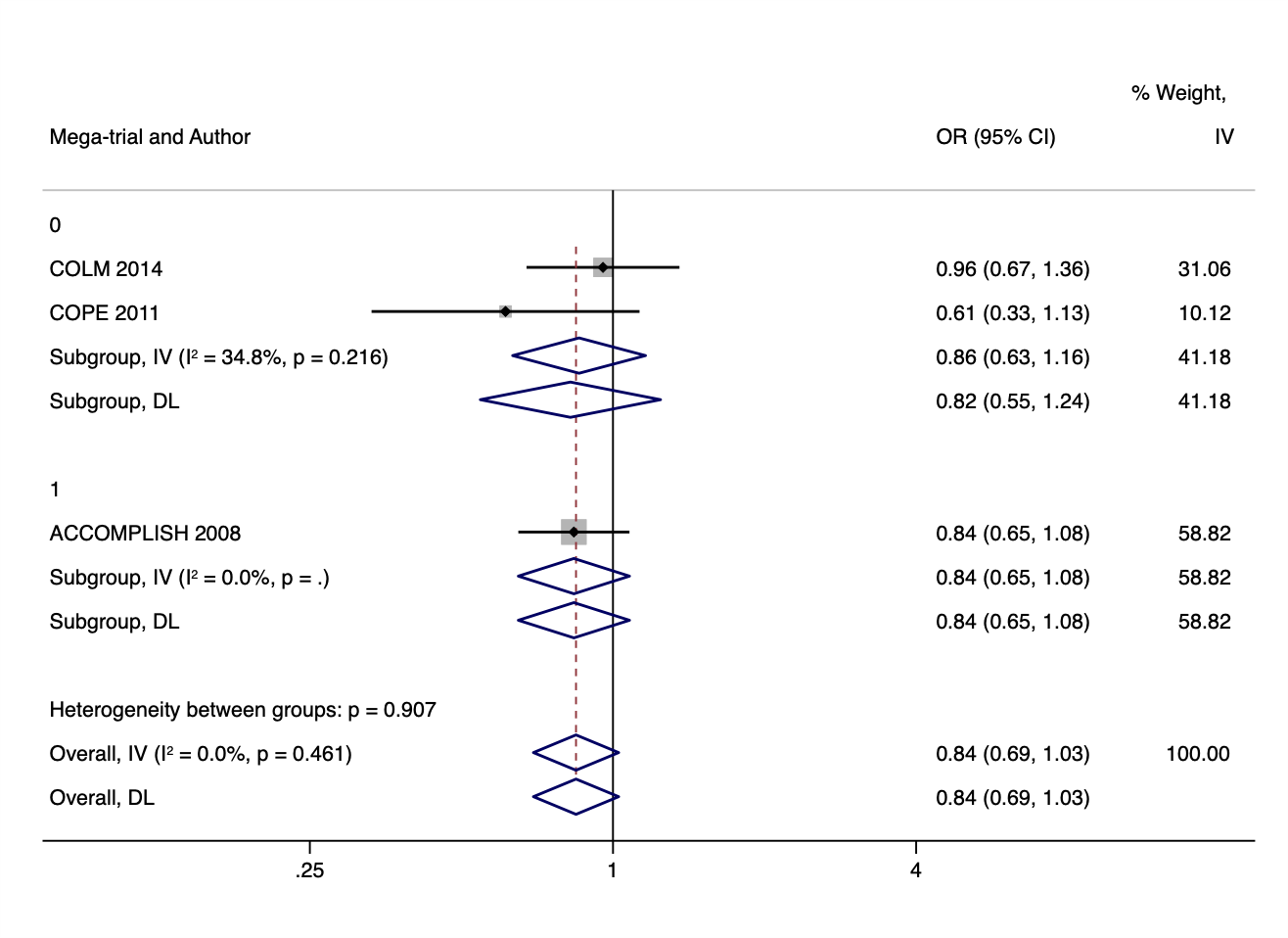

3. **CSPTT 2015**

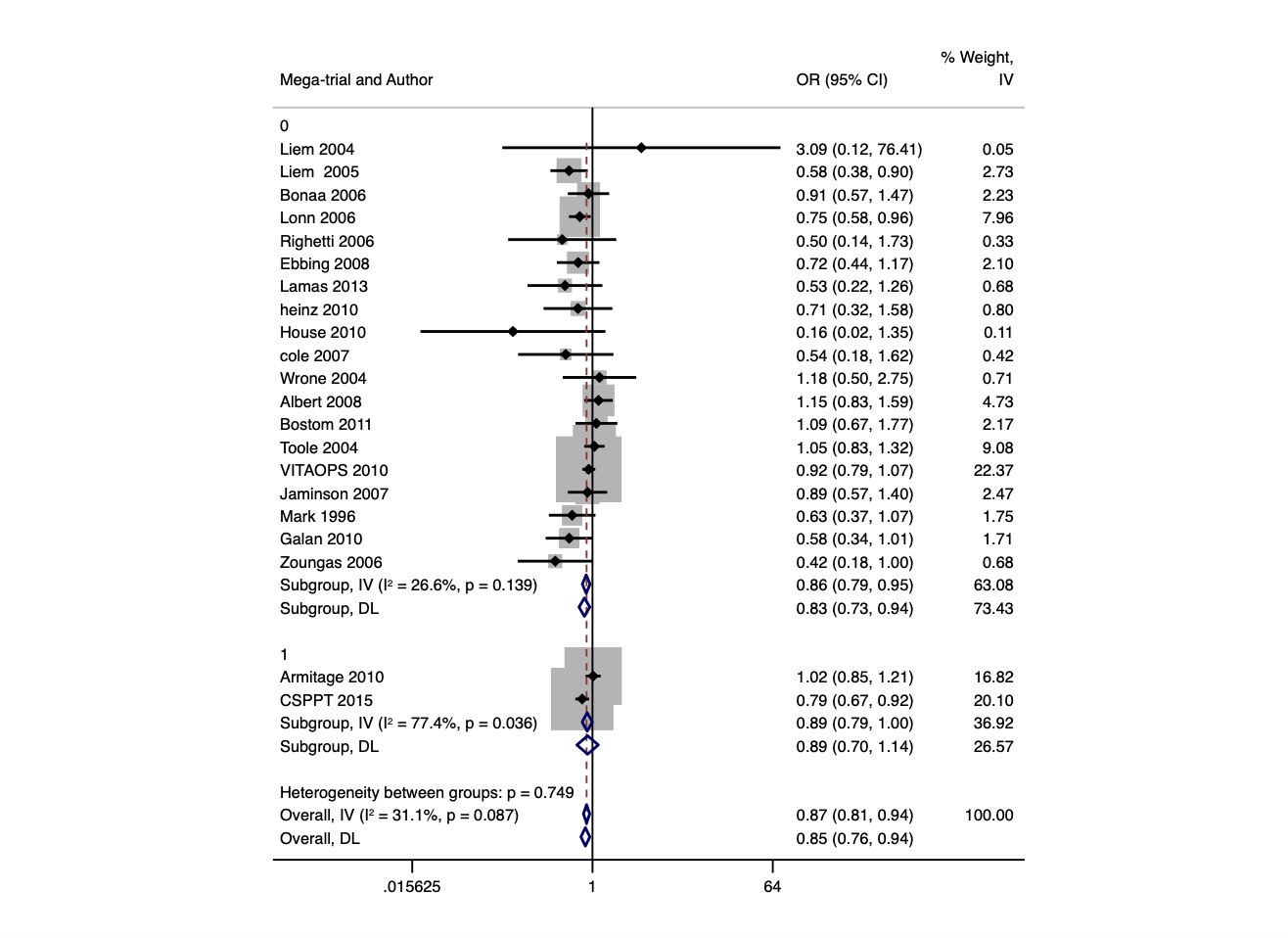

4. **ORIGIN 2012**

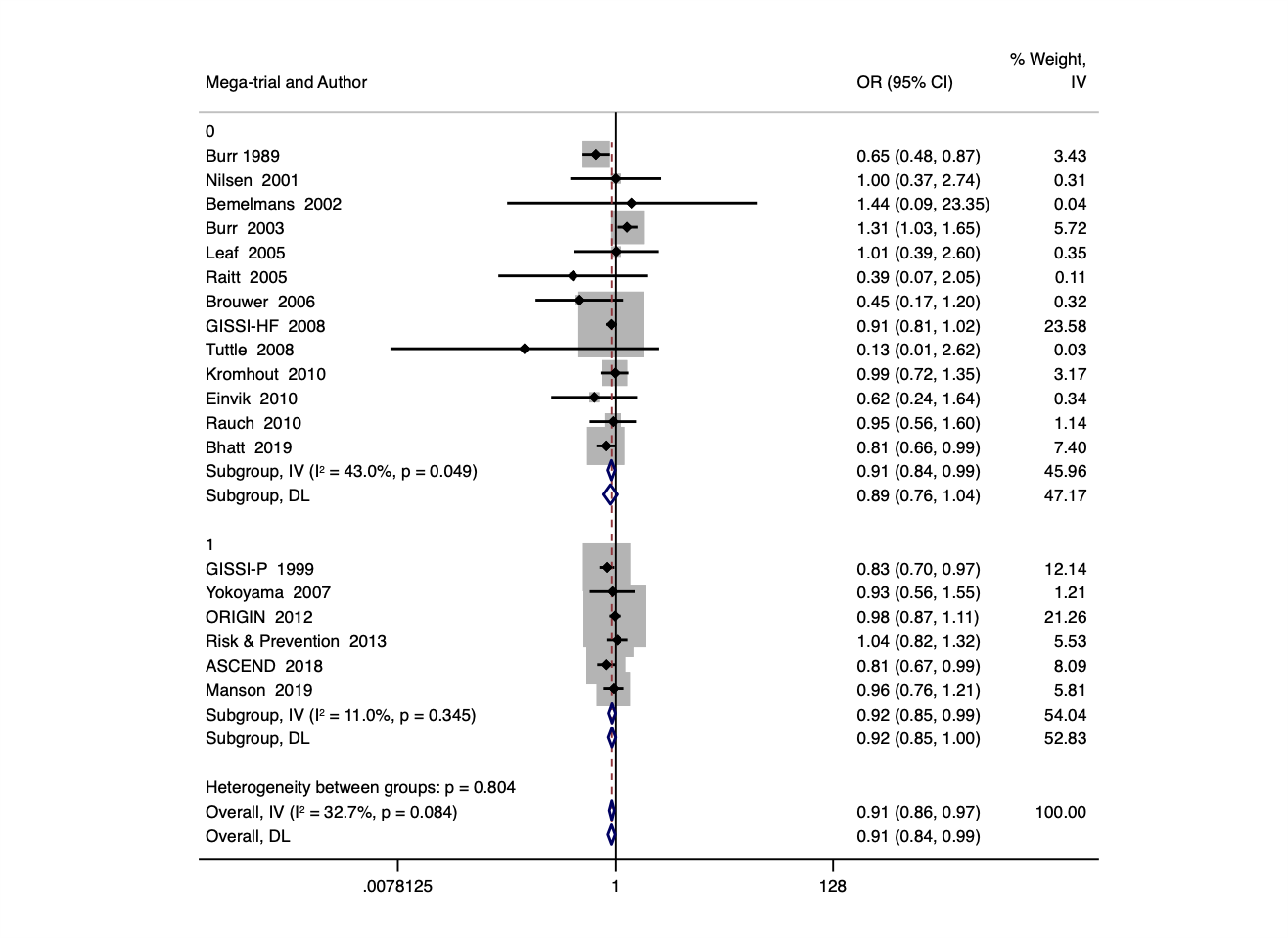

5. **GLOBAL LEADERS 2018**

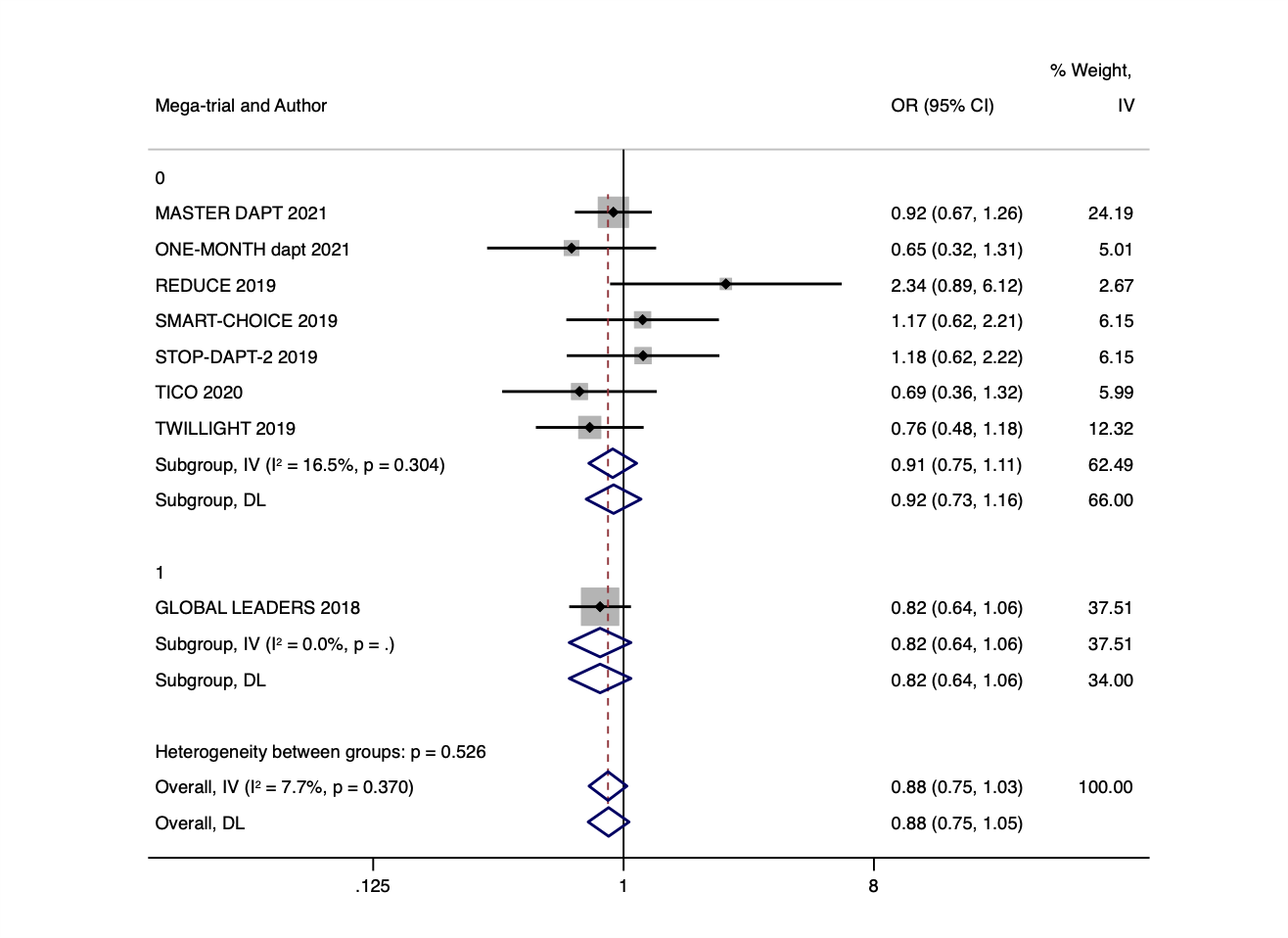

6. **ACCORD 2008**

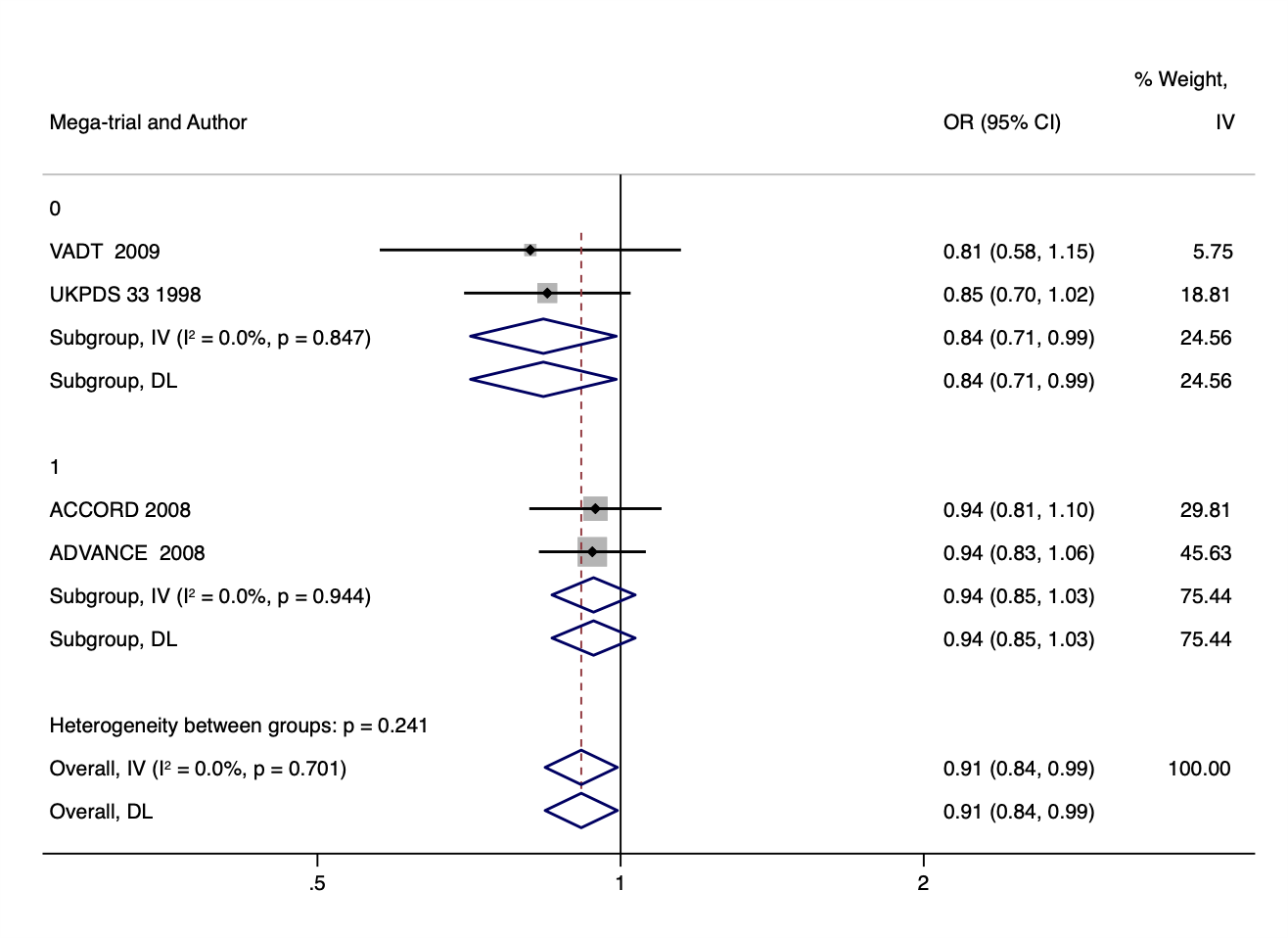

7. **INVEST 2003**

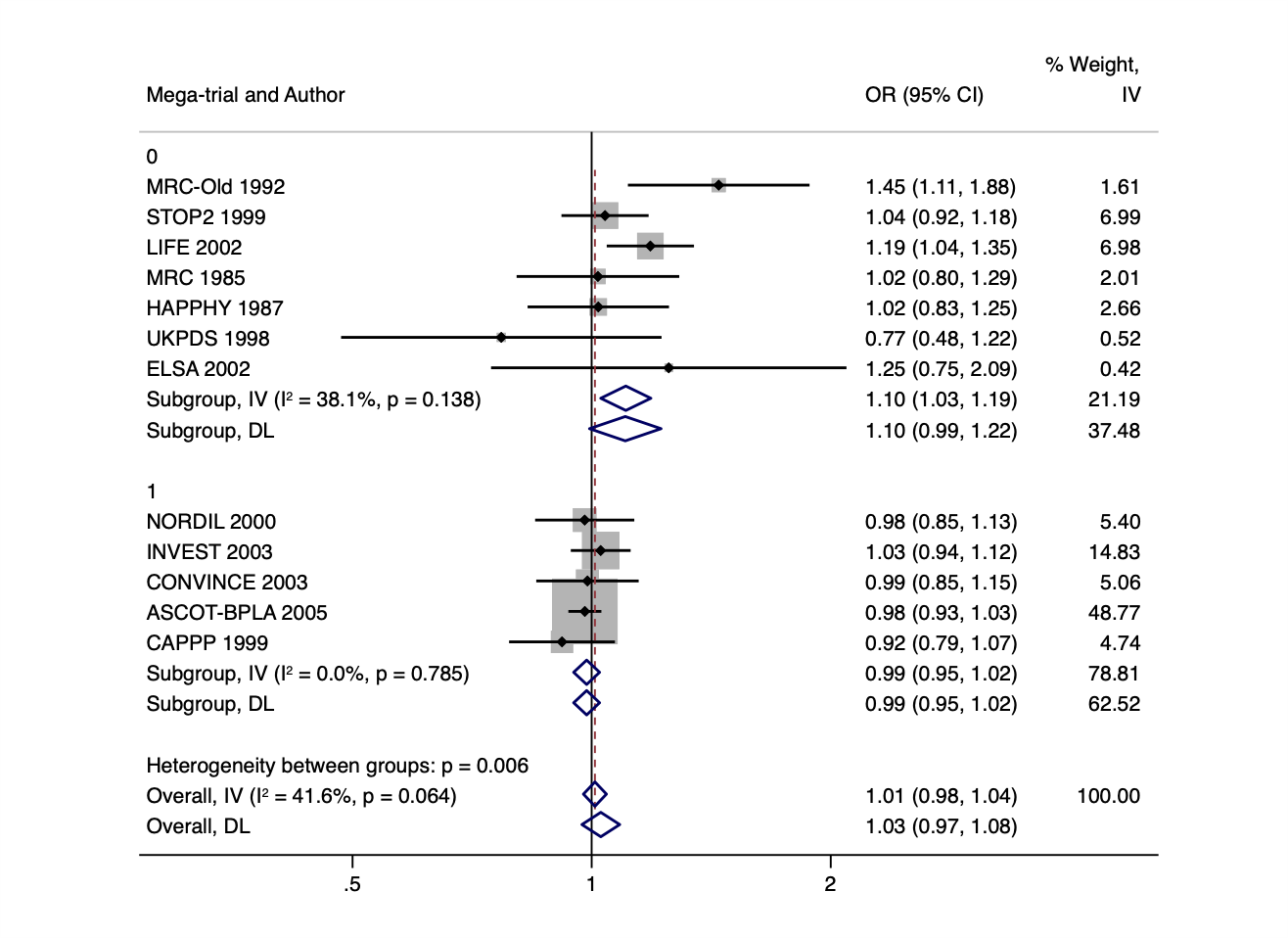

8. **JELIS 2007**

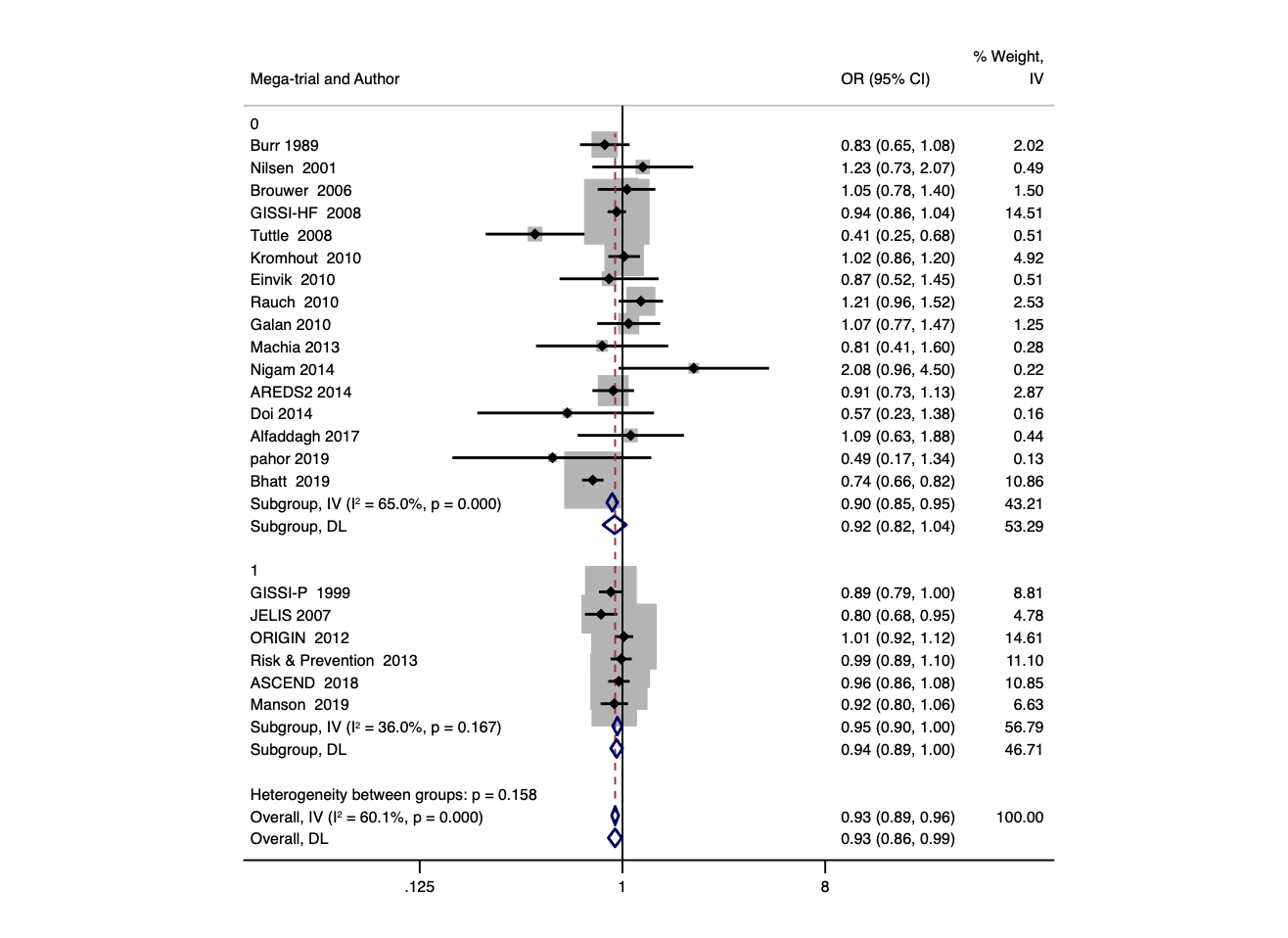

9. **JPPP 2014**

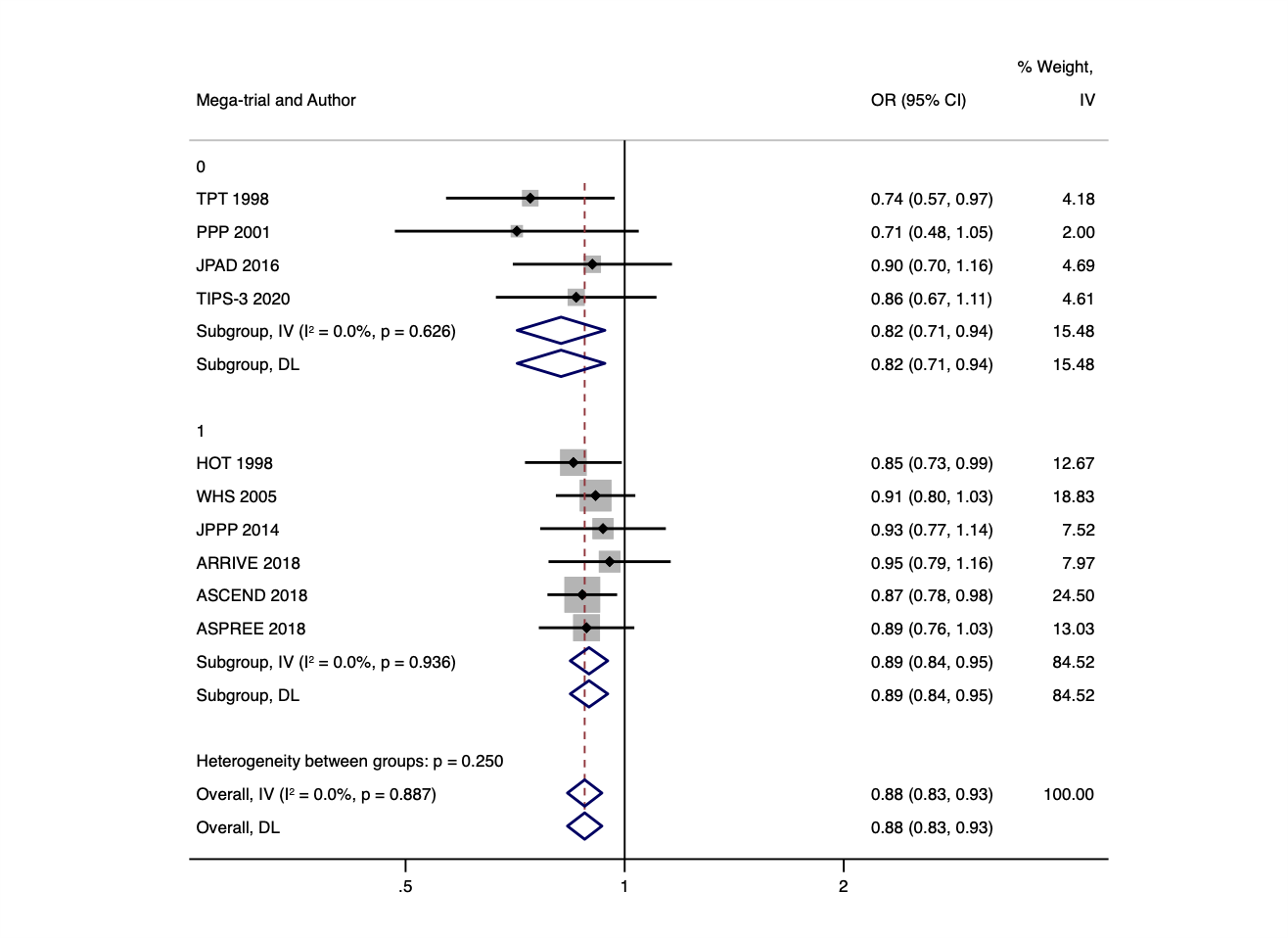

10. **VITAL 2019**

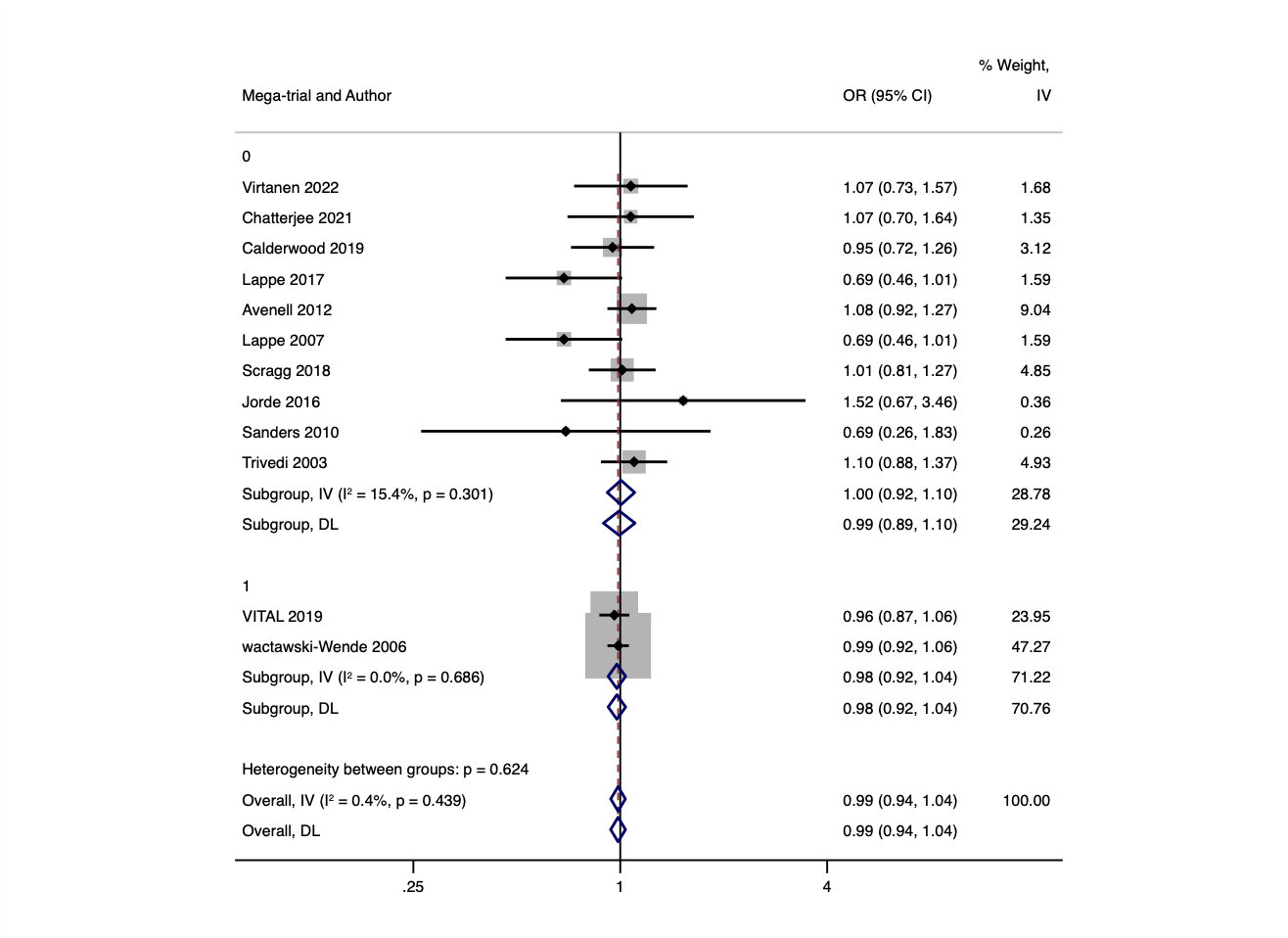

11. **Albert 2021**

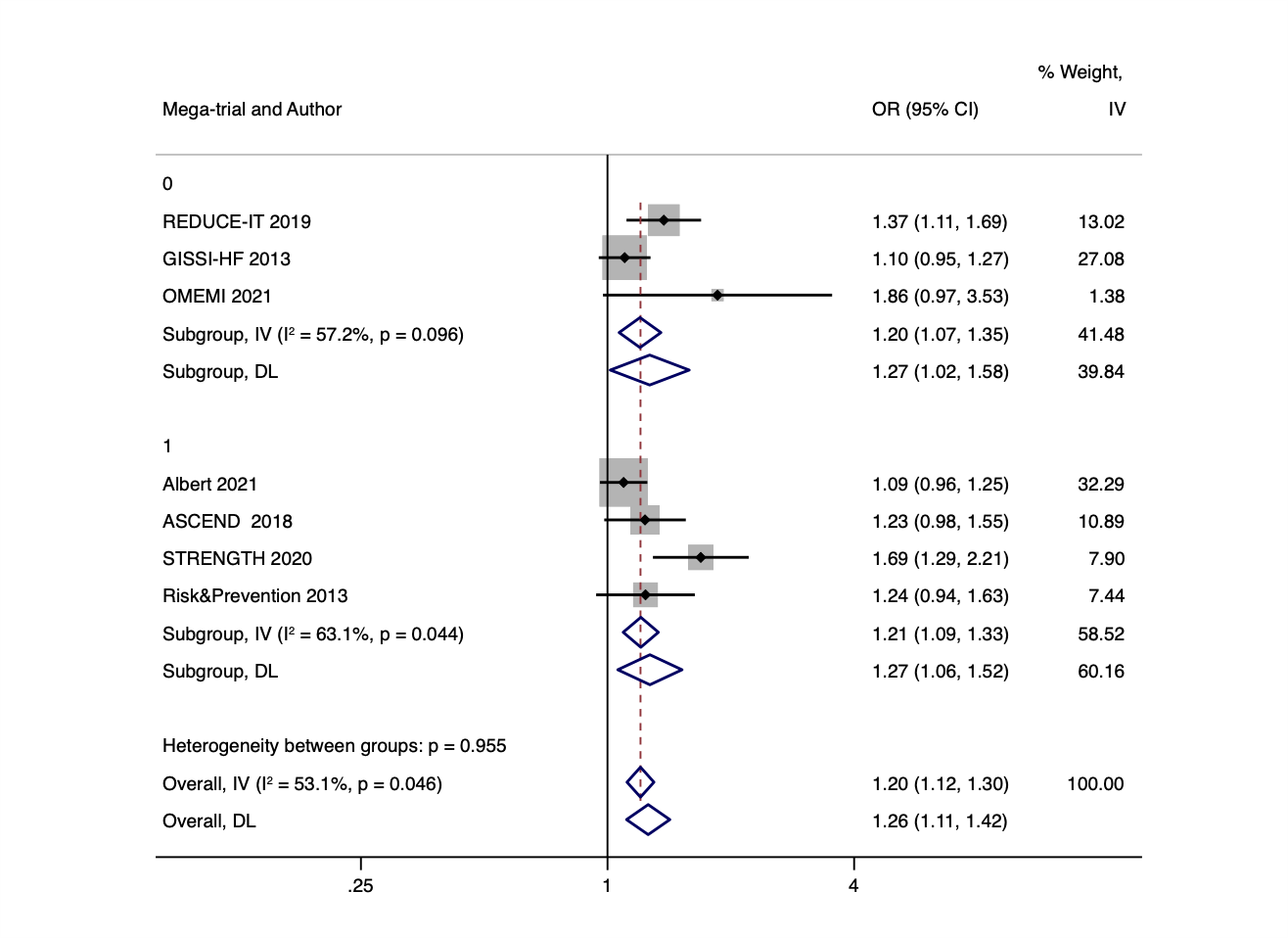

12. **NISSEN 2016**

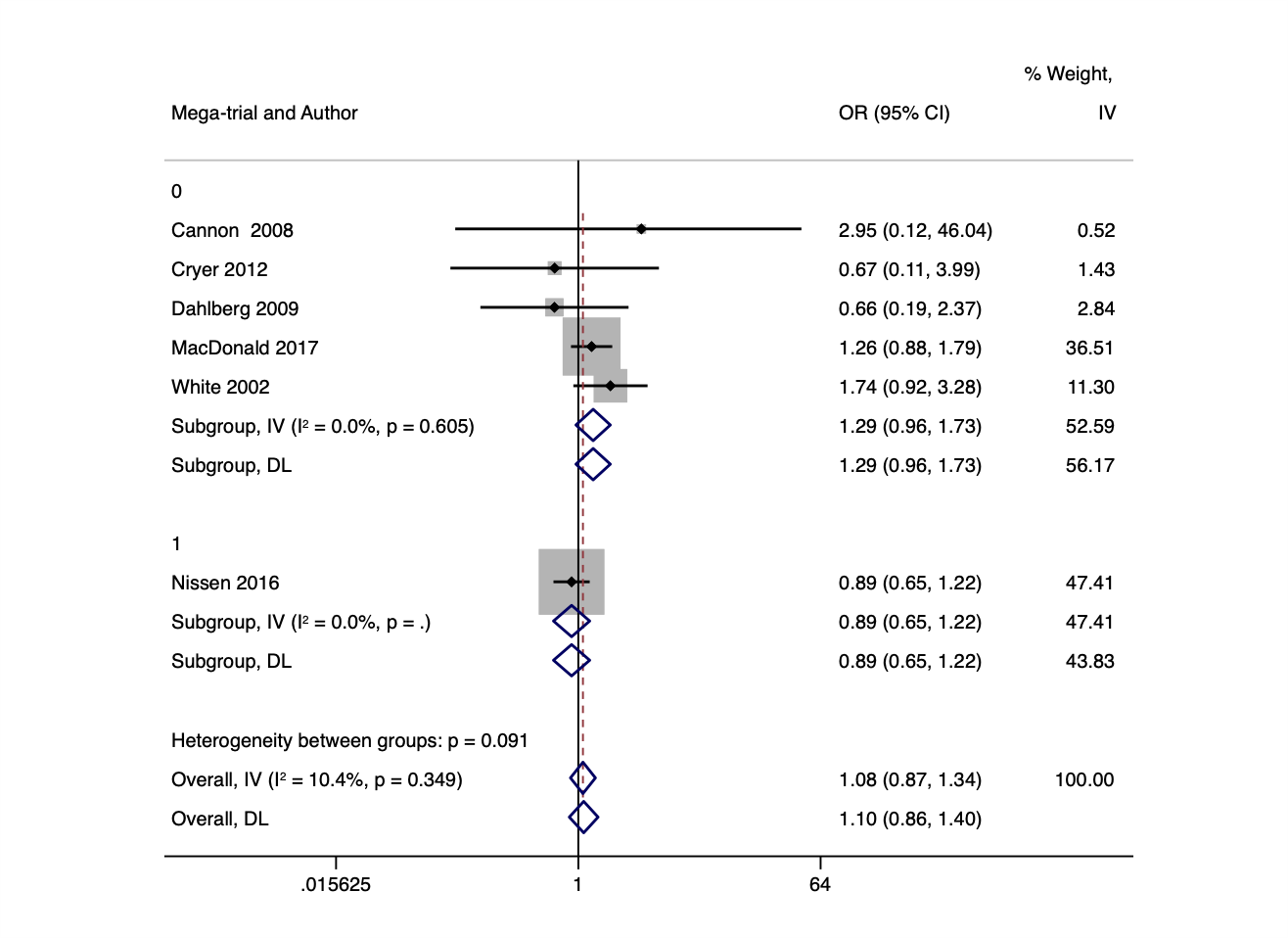

13. **PROFESS 2008**

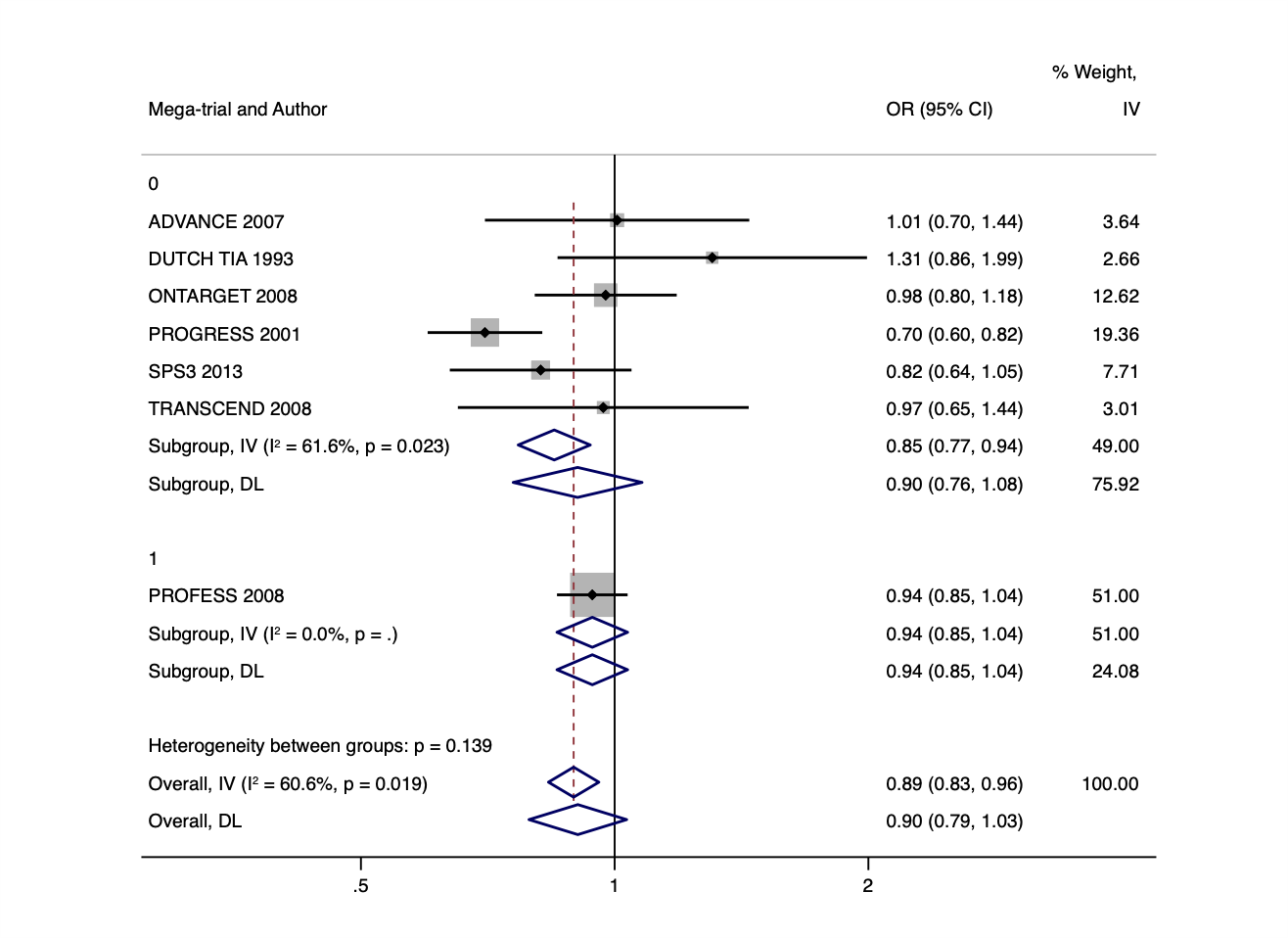

14. **EXTRACT TIMI 25 2006**

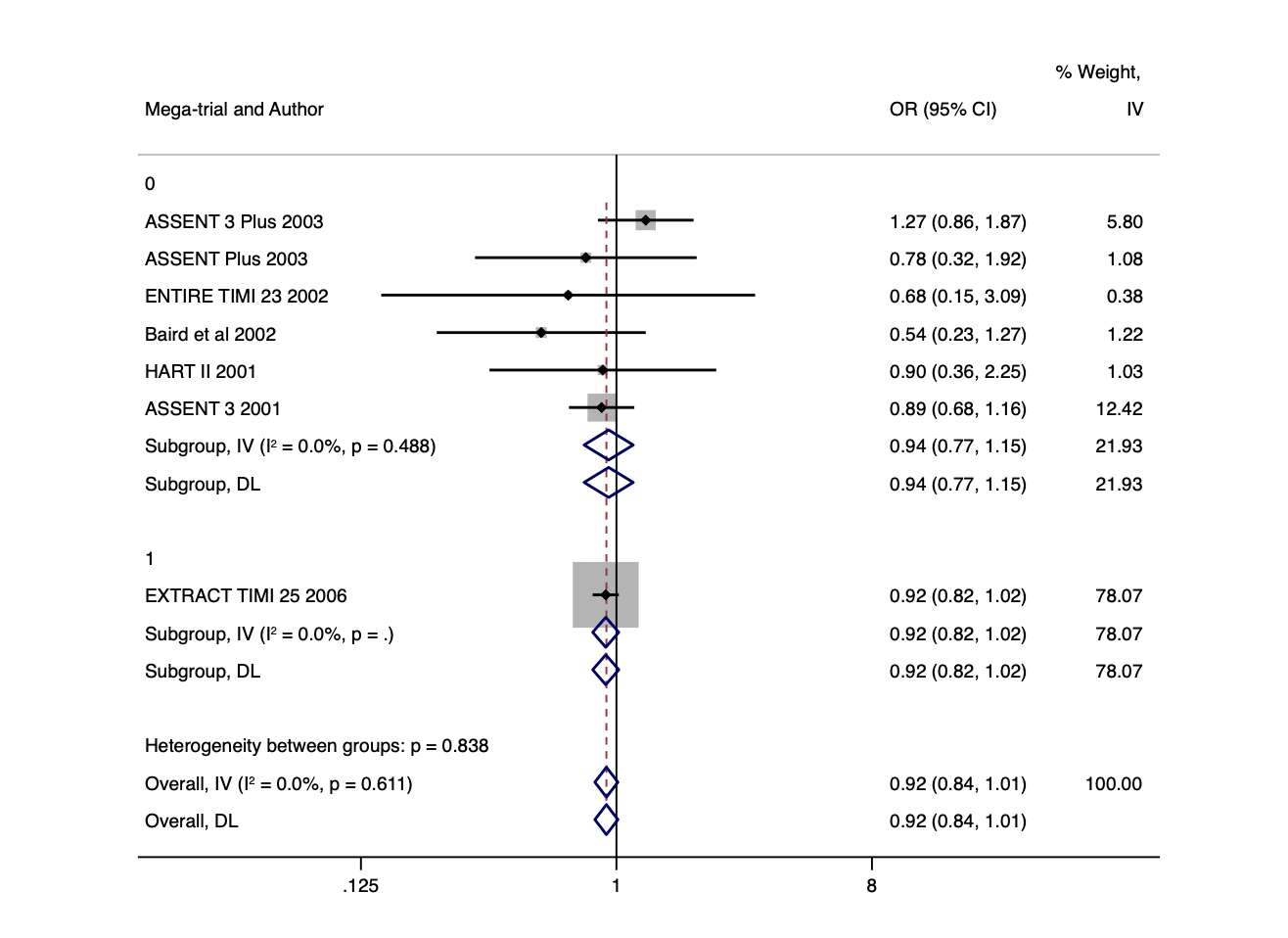

15. **ODYSSEY OUTCOMES 2019**

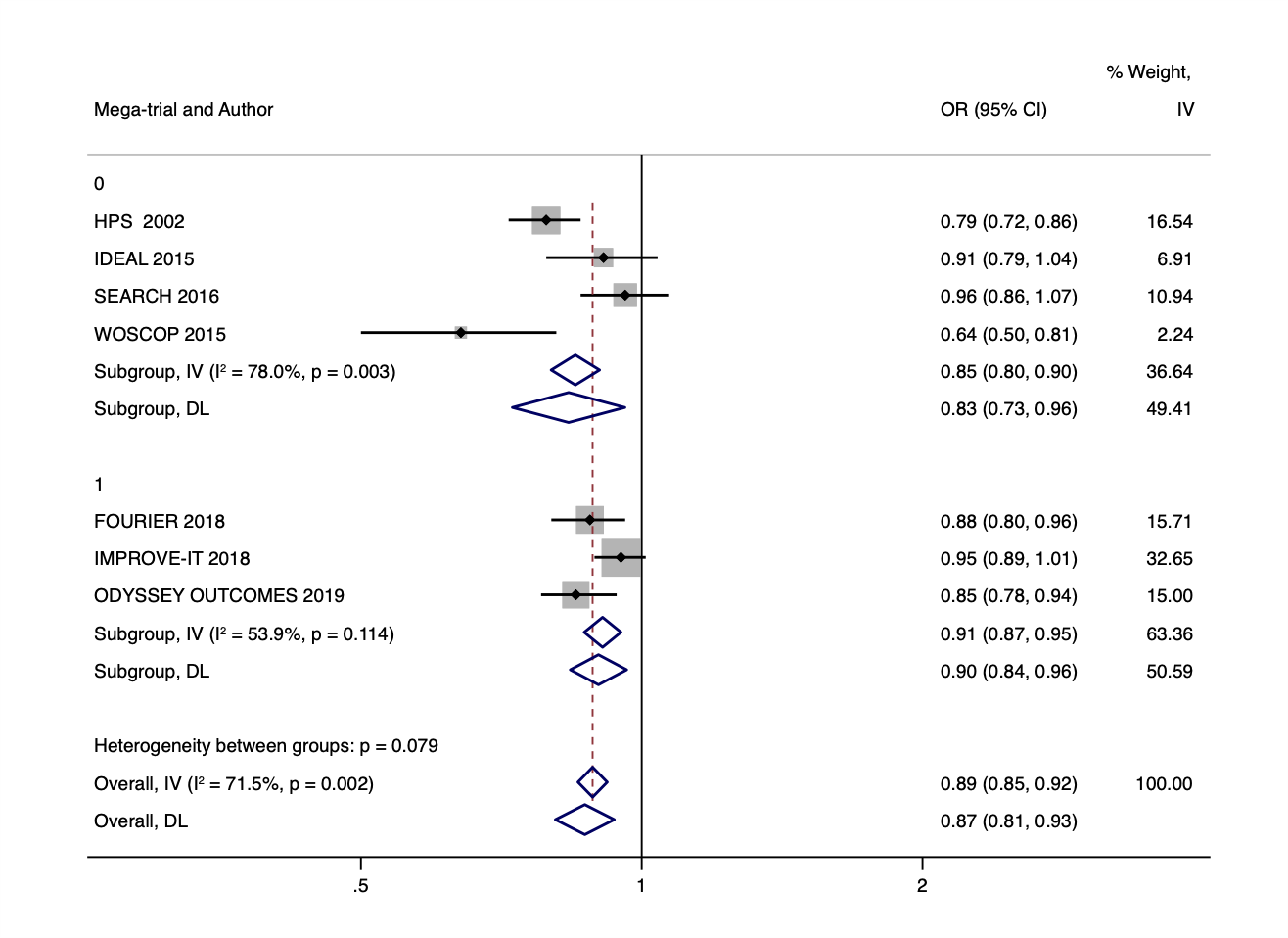

16. **BaSICS 2021**

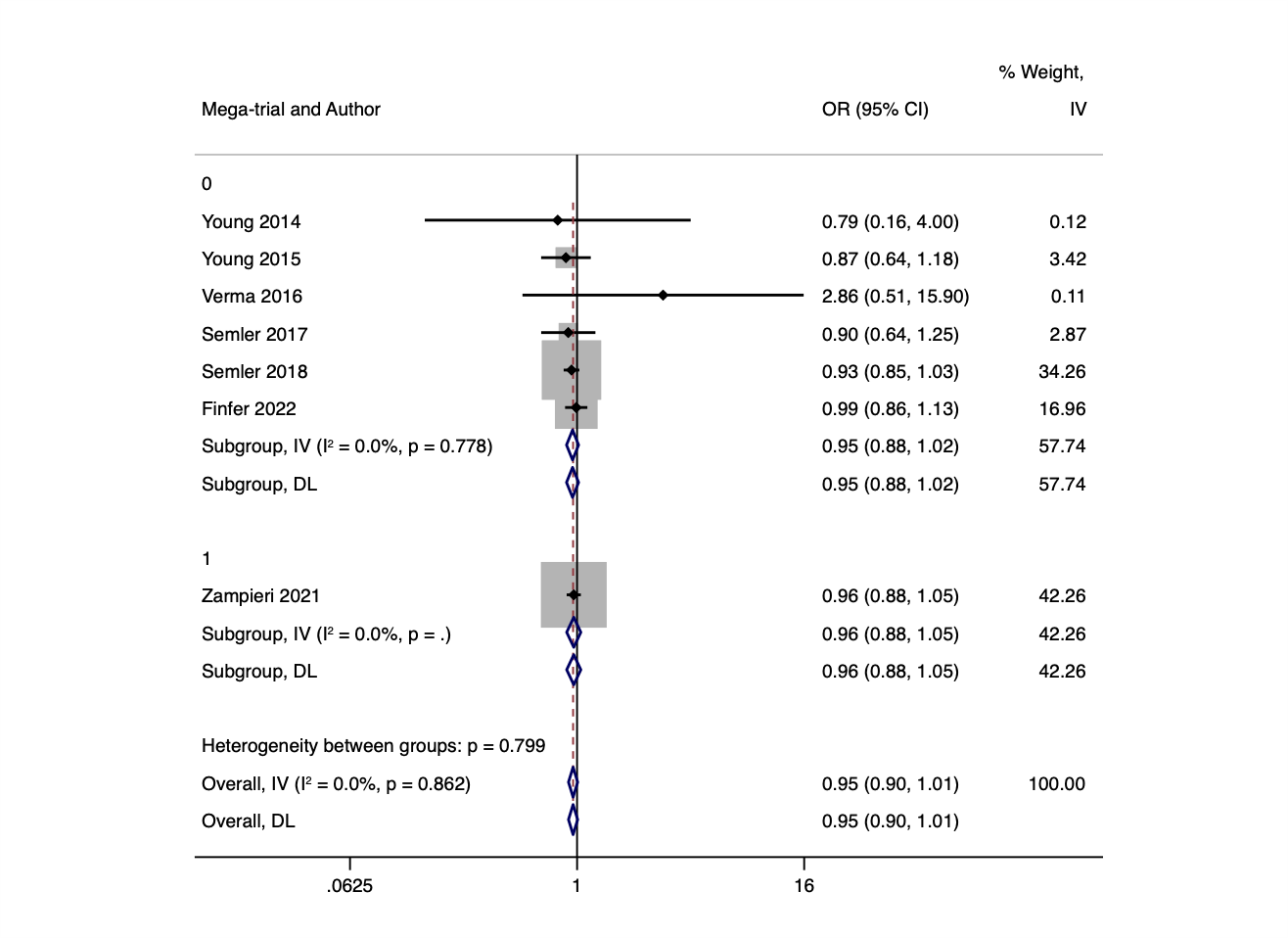

17. **BEAUTIFUL trial 2008**

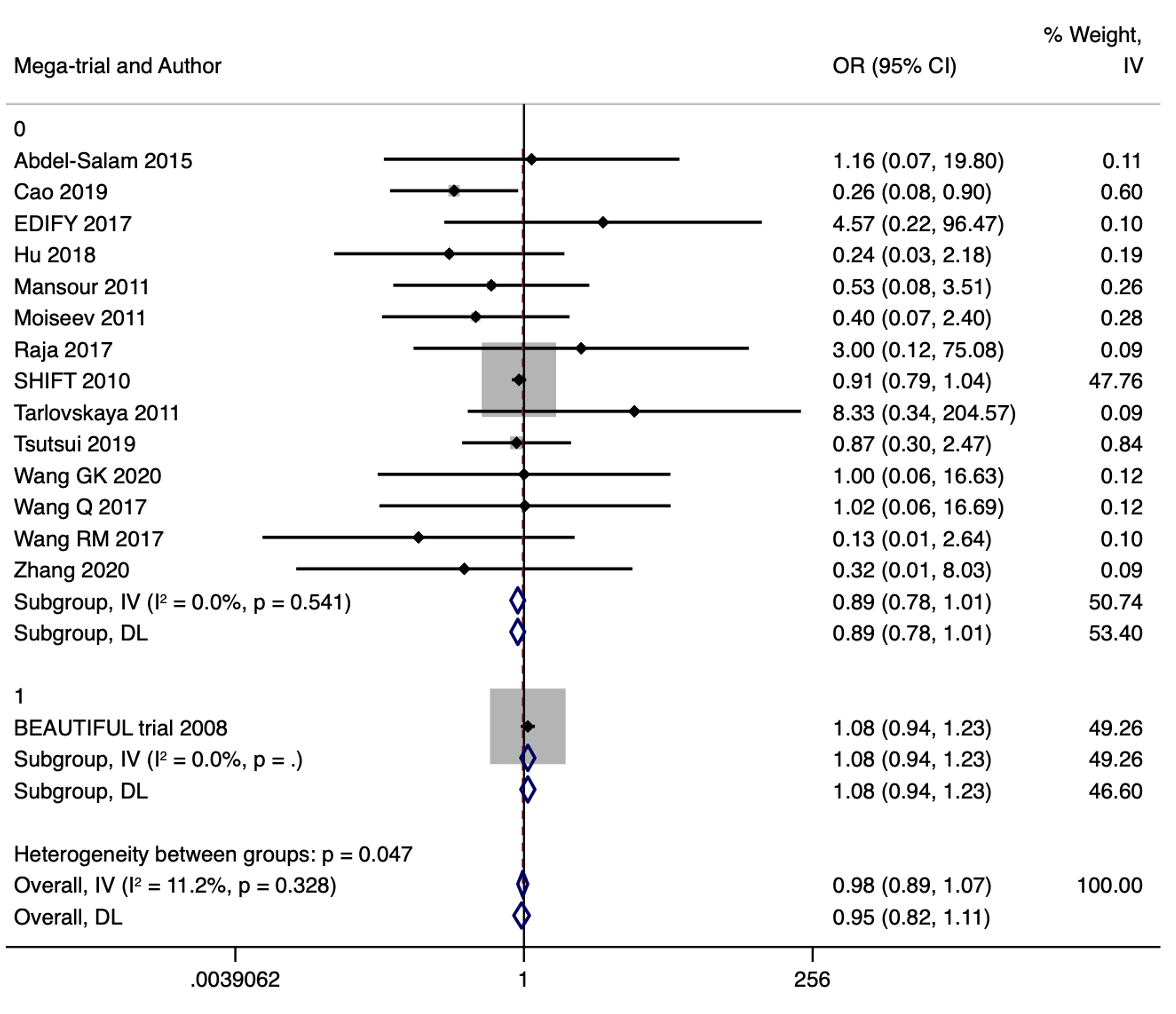

18. **SU.VI.MAX 2004**

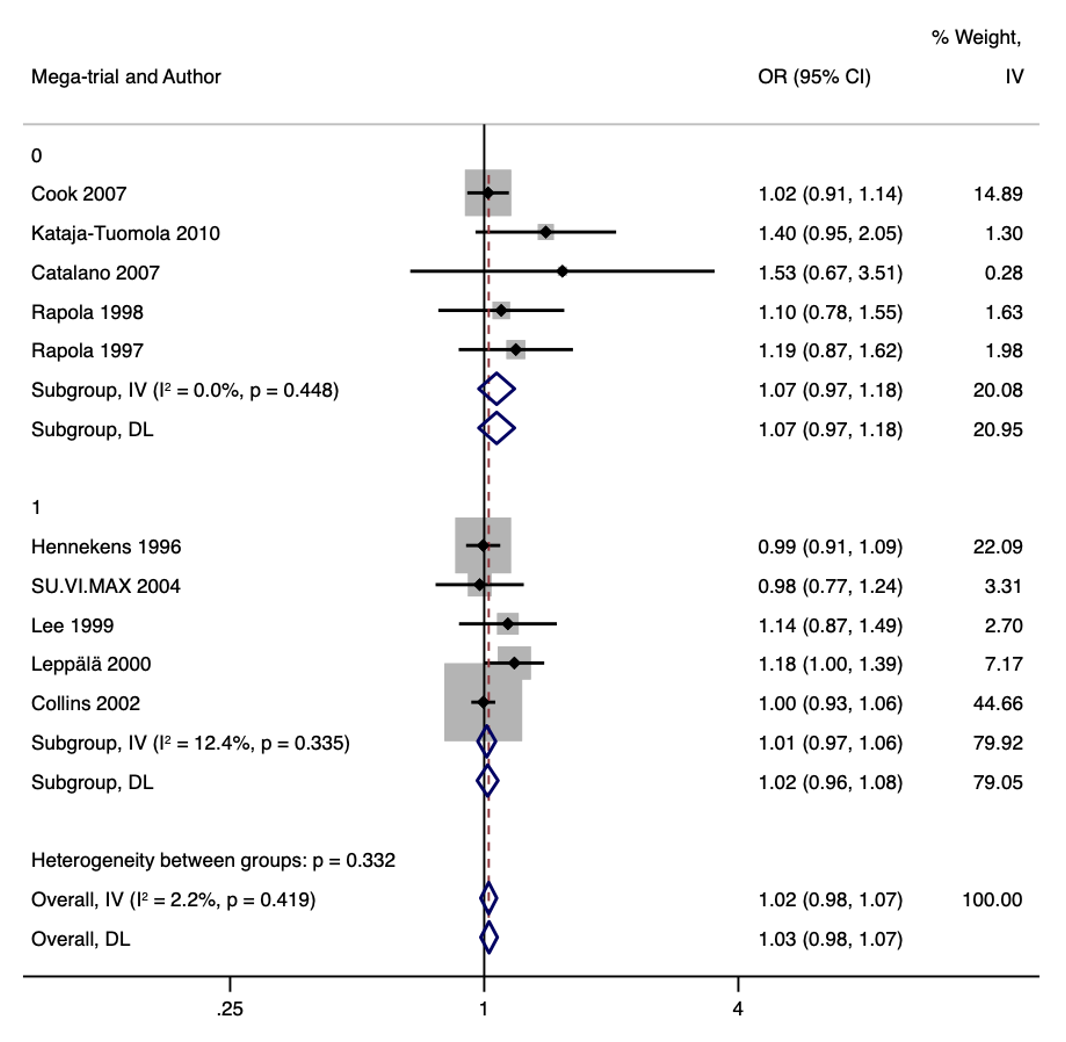

19. **HPS2-THRIVE 2014**

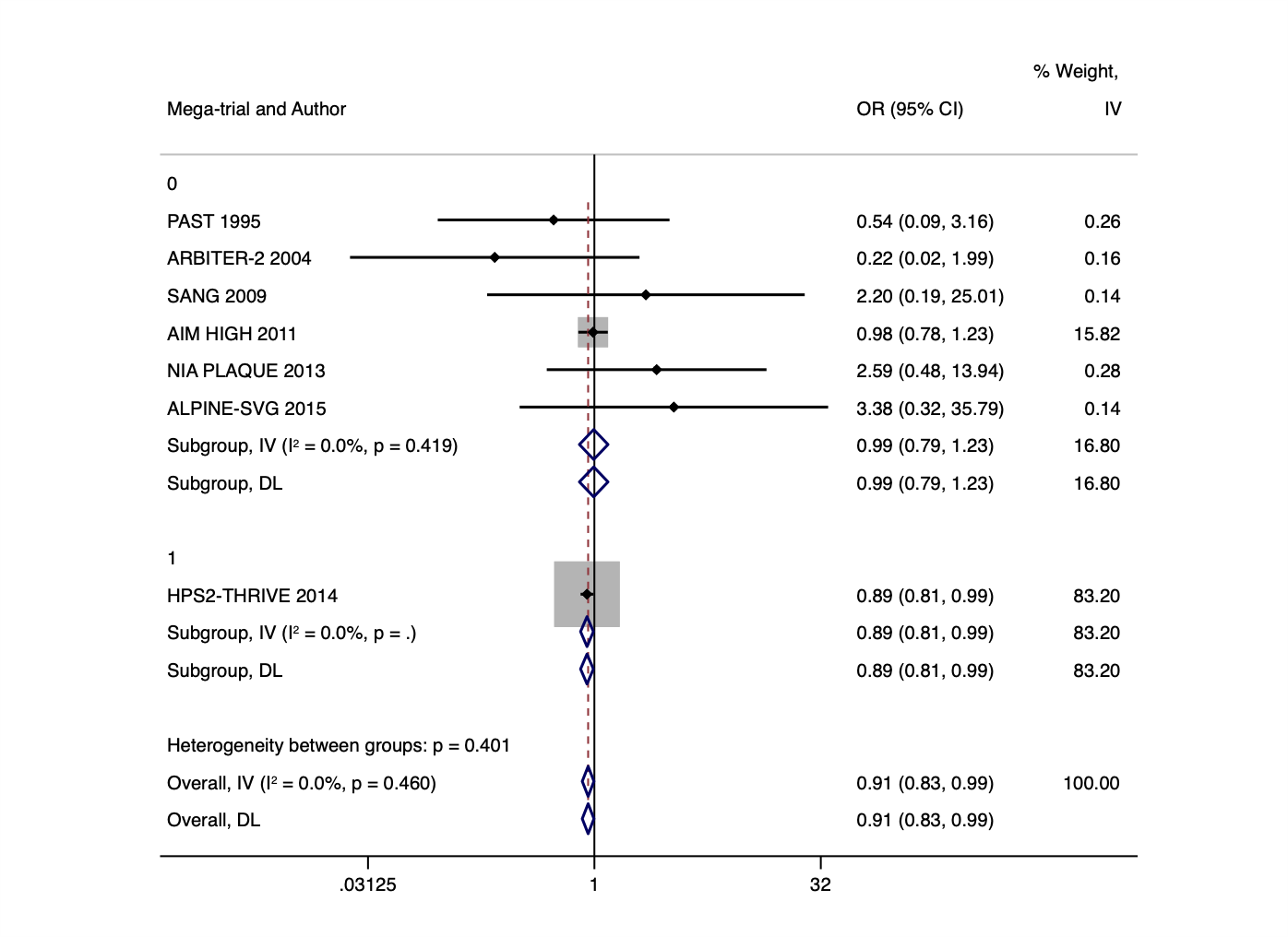

20. **ILLUMINATE 2007**
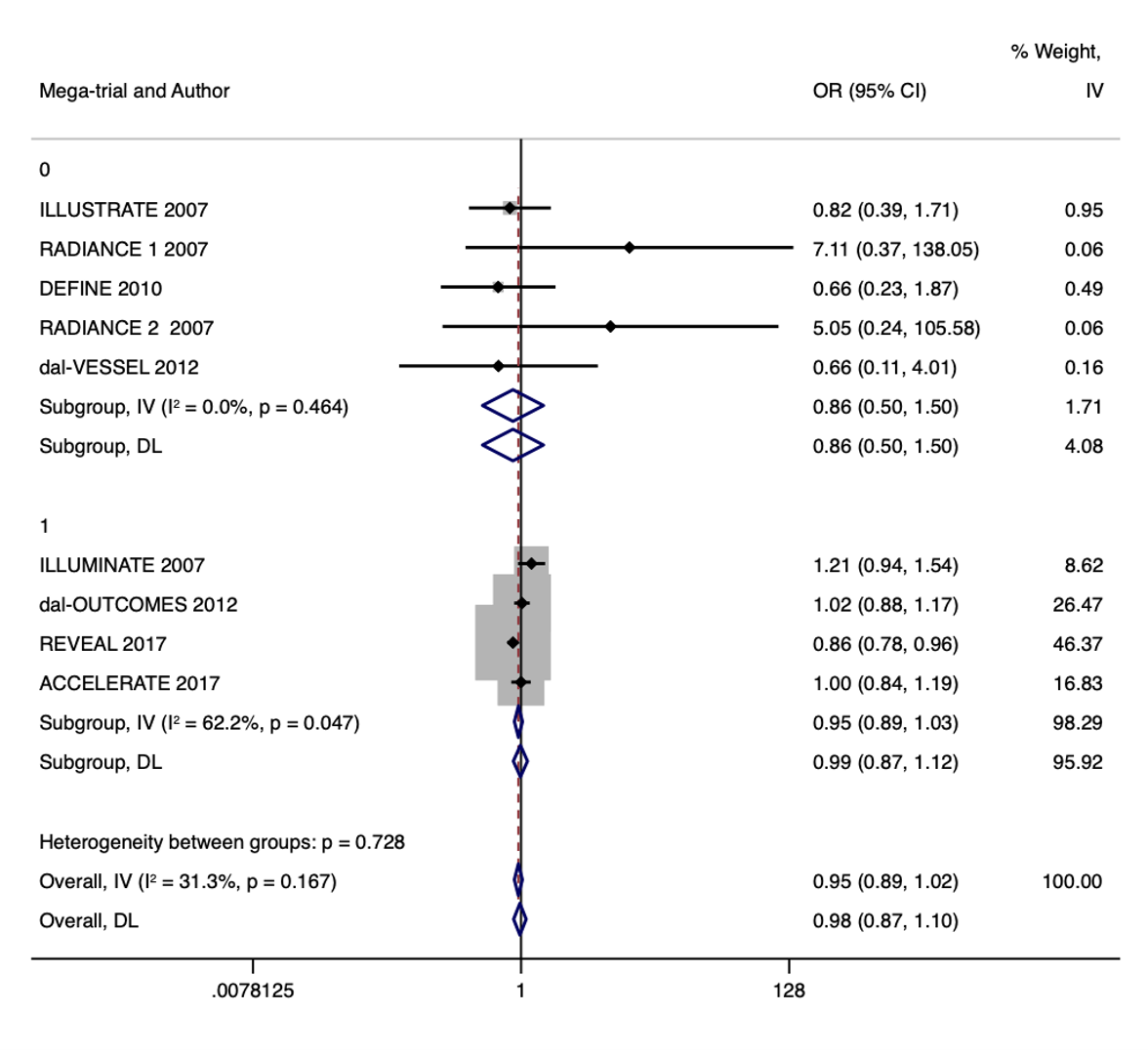

21. **SAVOR-TIMI 53 2013**

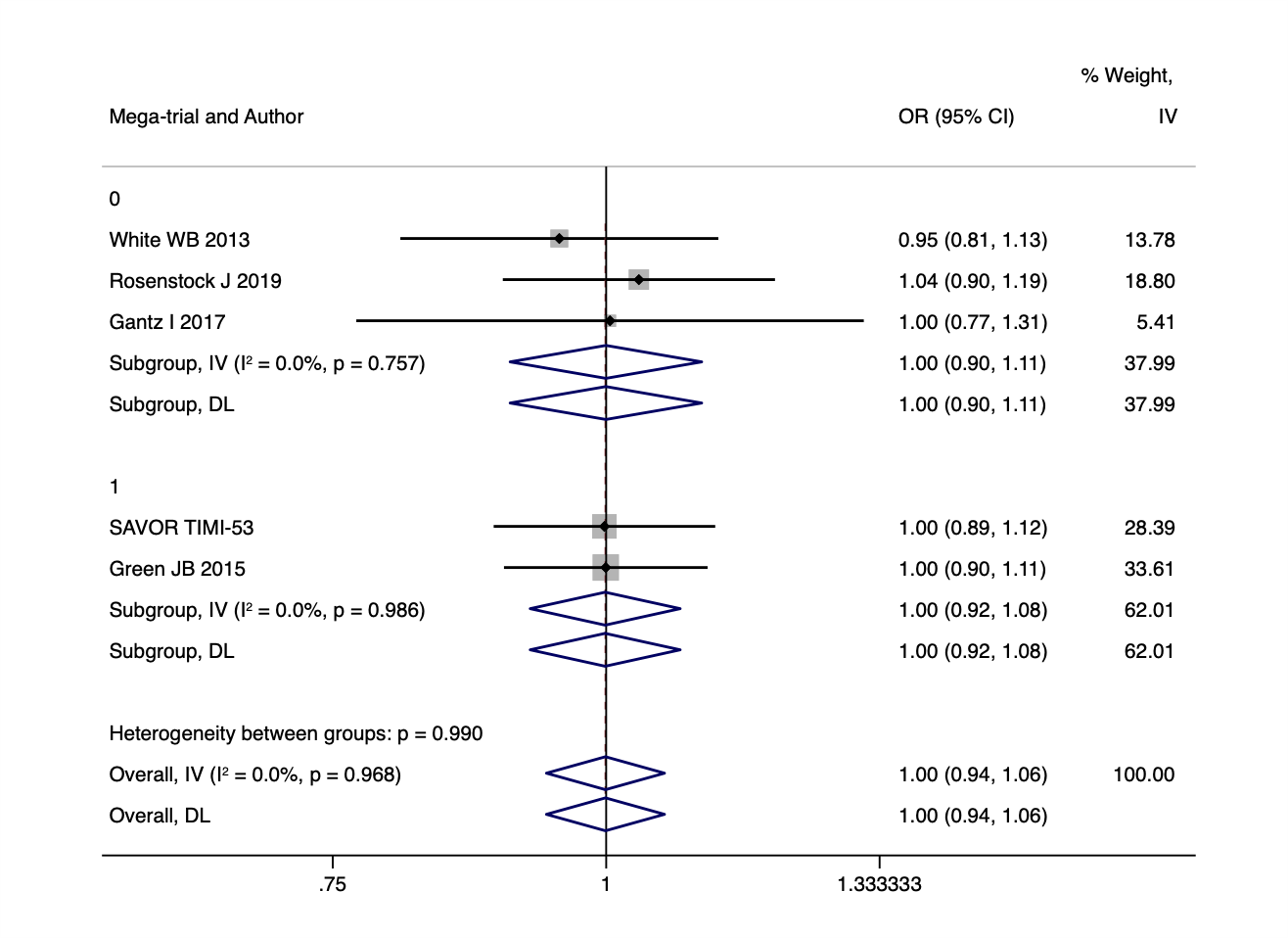

22. **SCORED 2021**

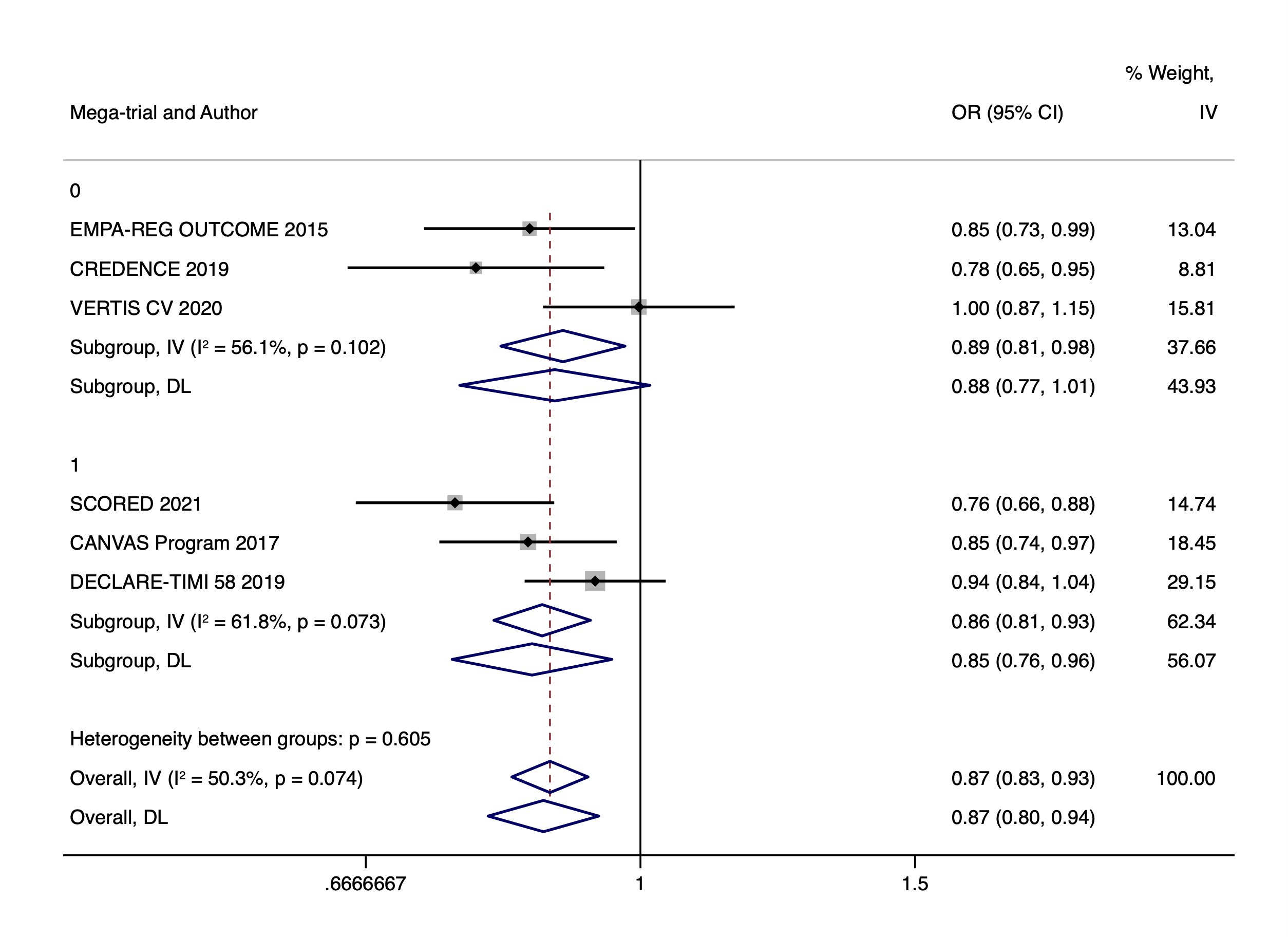

23. **STABILITY 2017**

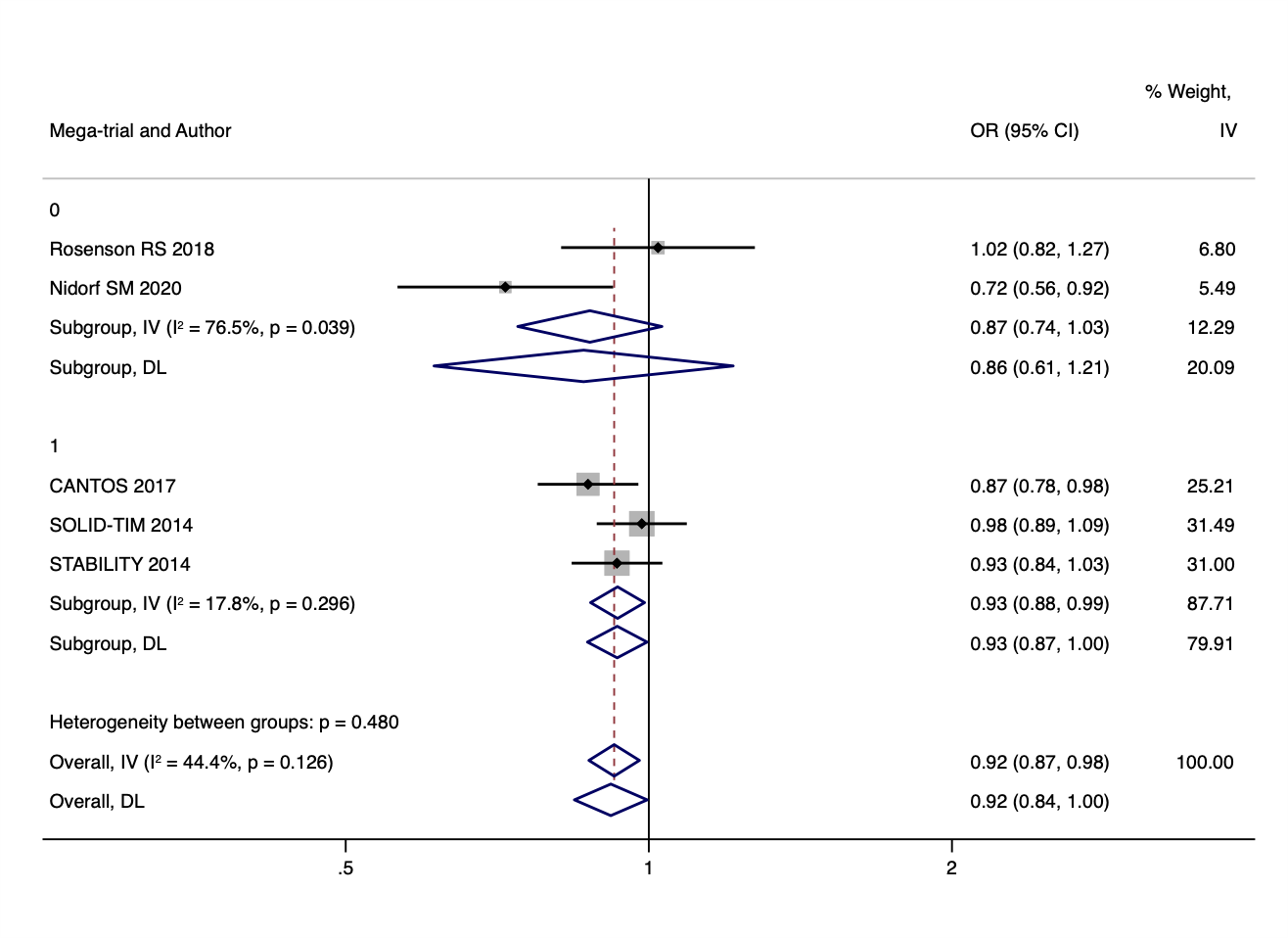

24. **EXCSCEL 2017**

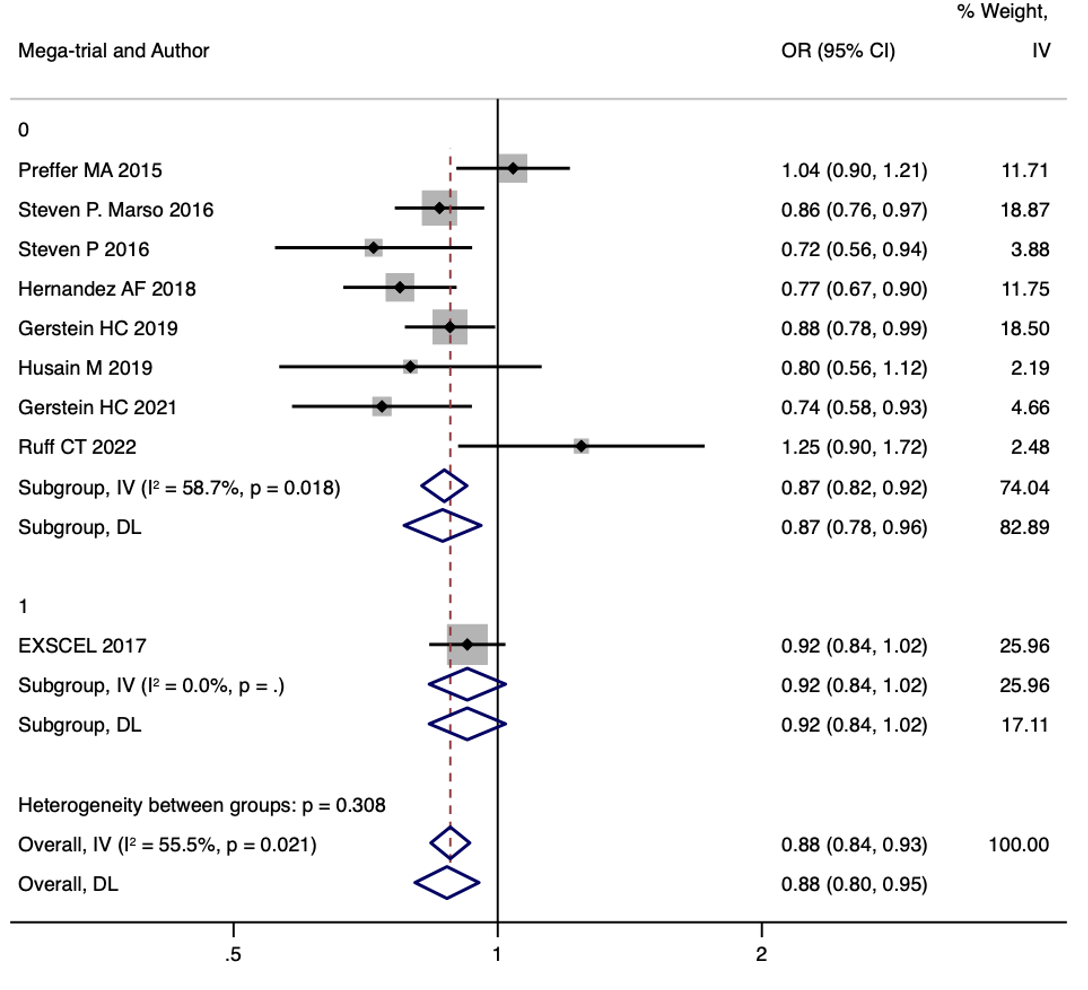

25. **CURRENT OASIS 7 2010**

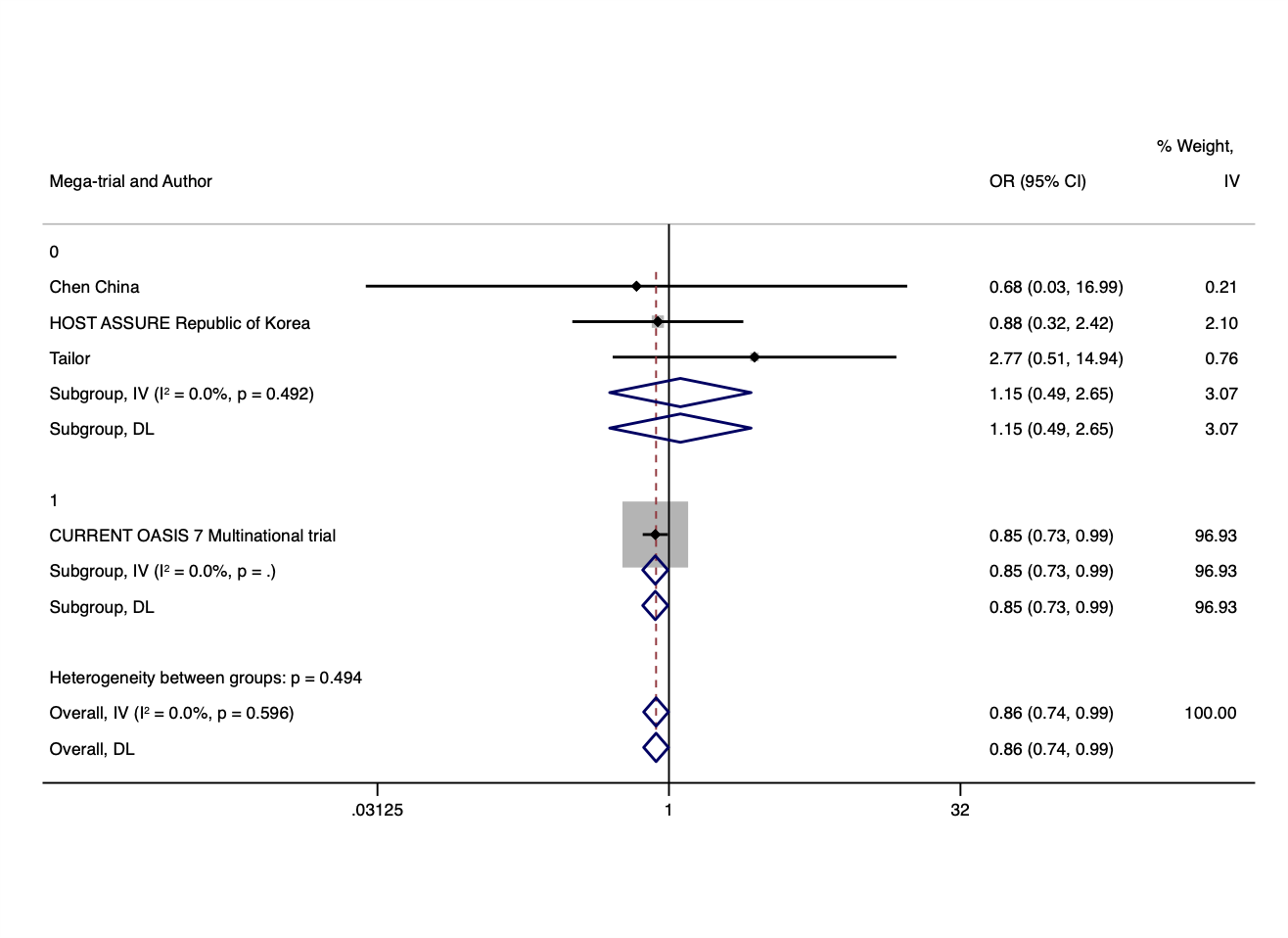

26. **POISE-2 2014**

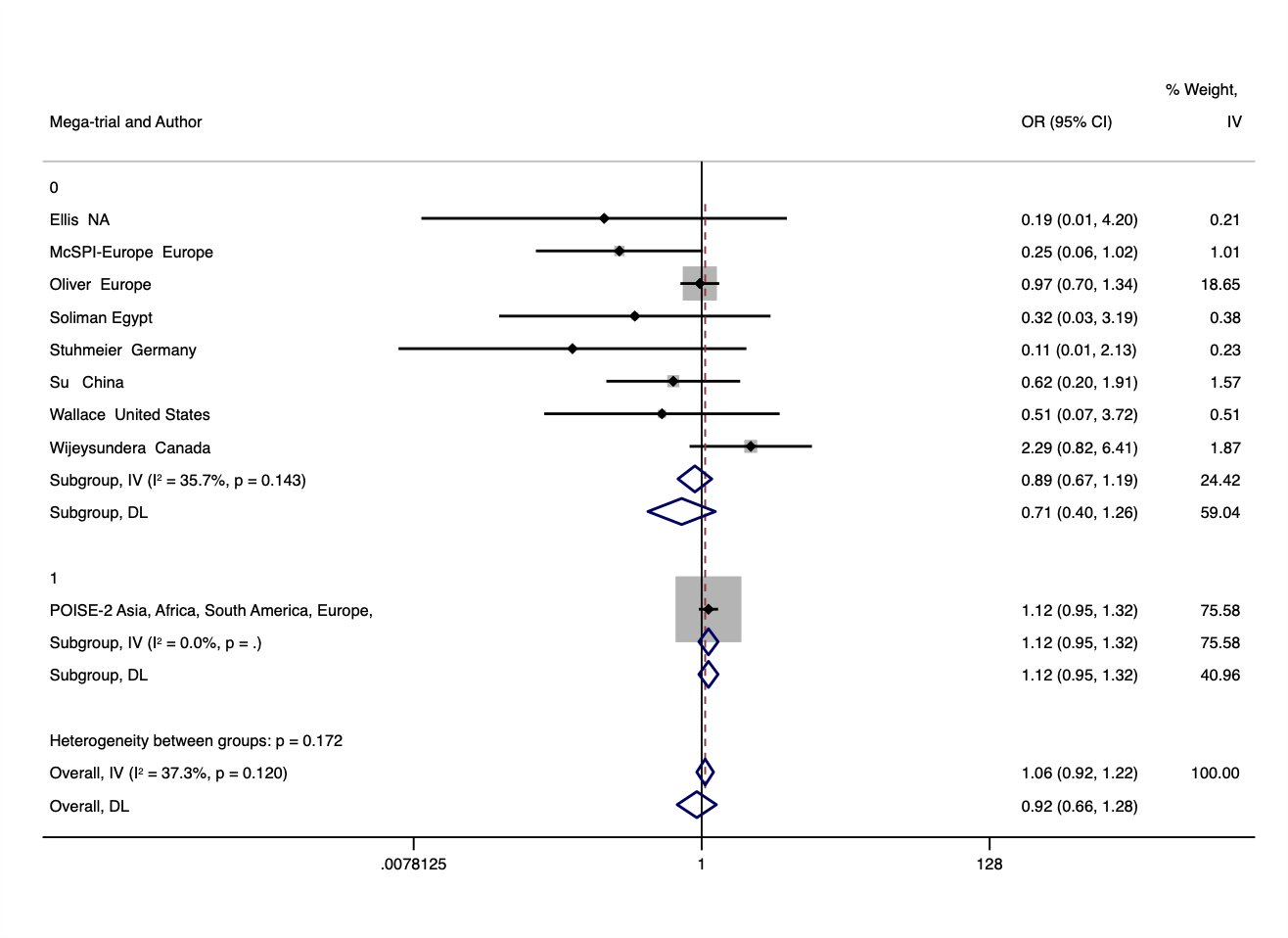

27. **CHARISMA 2006**

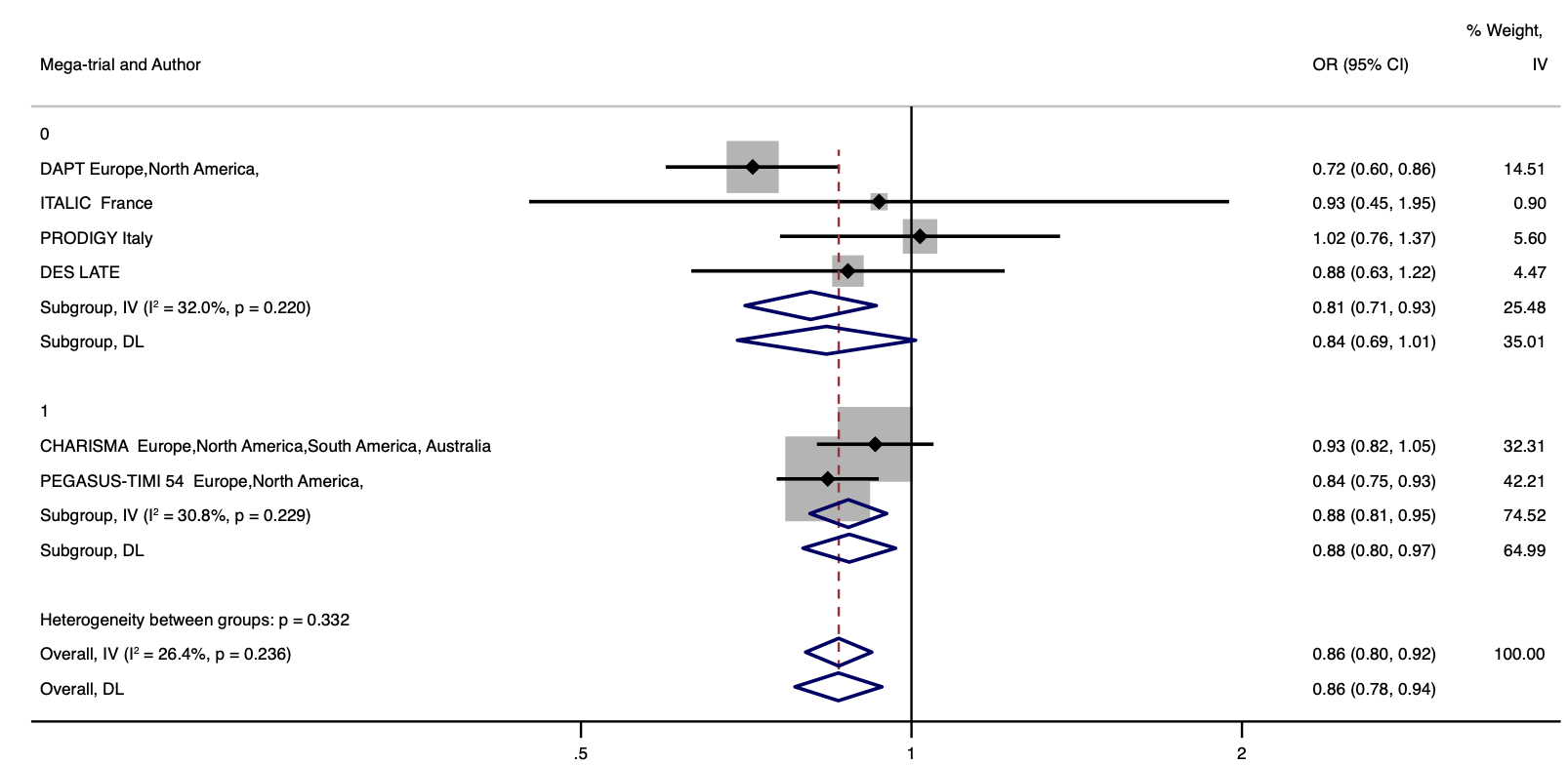

28. **ENGAGE TIMI 2013**

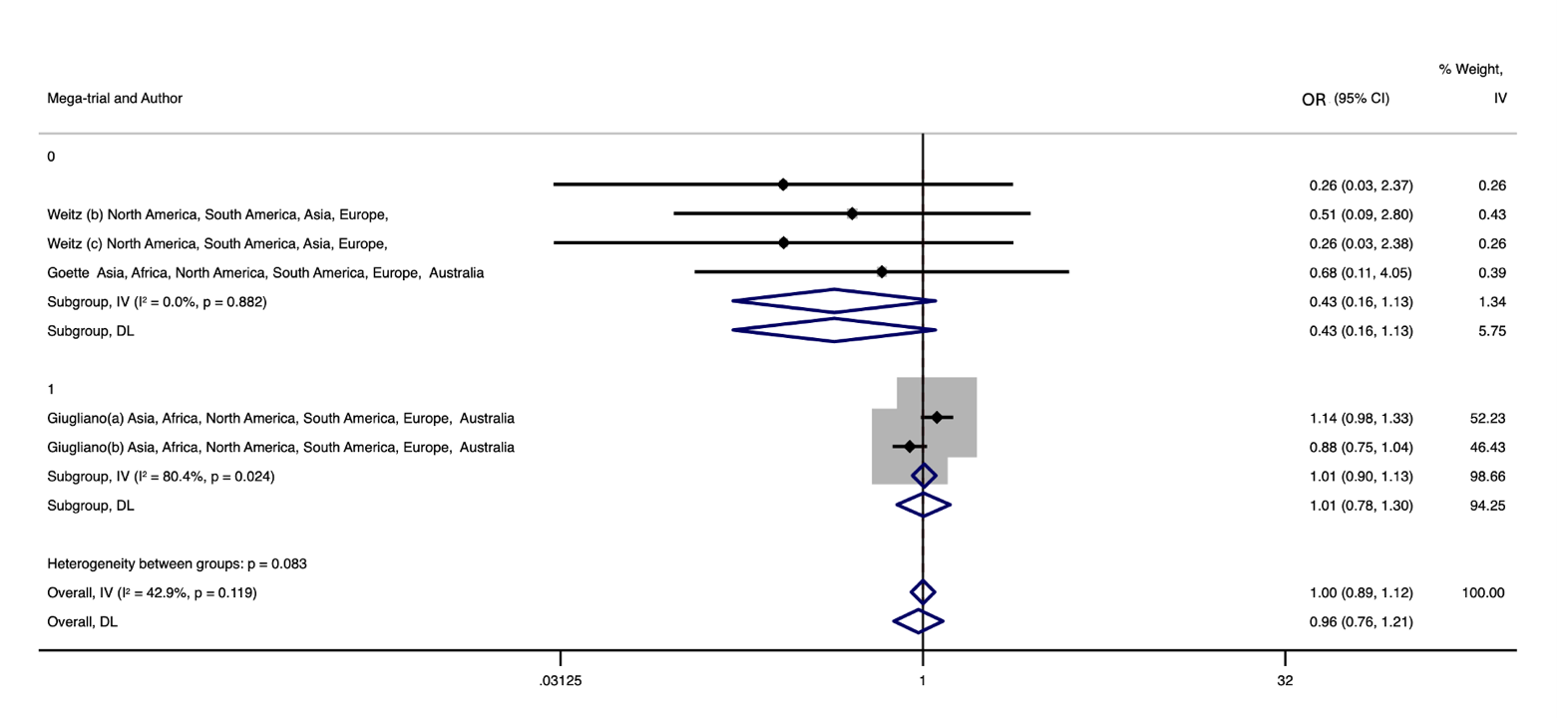

29. **ATLAS 2012**

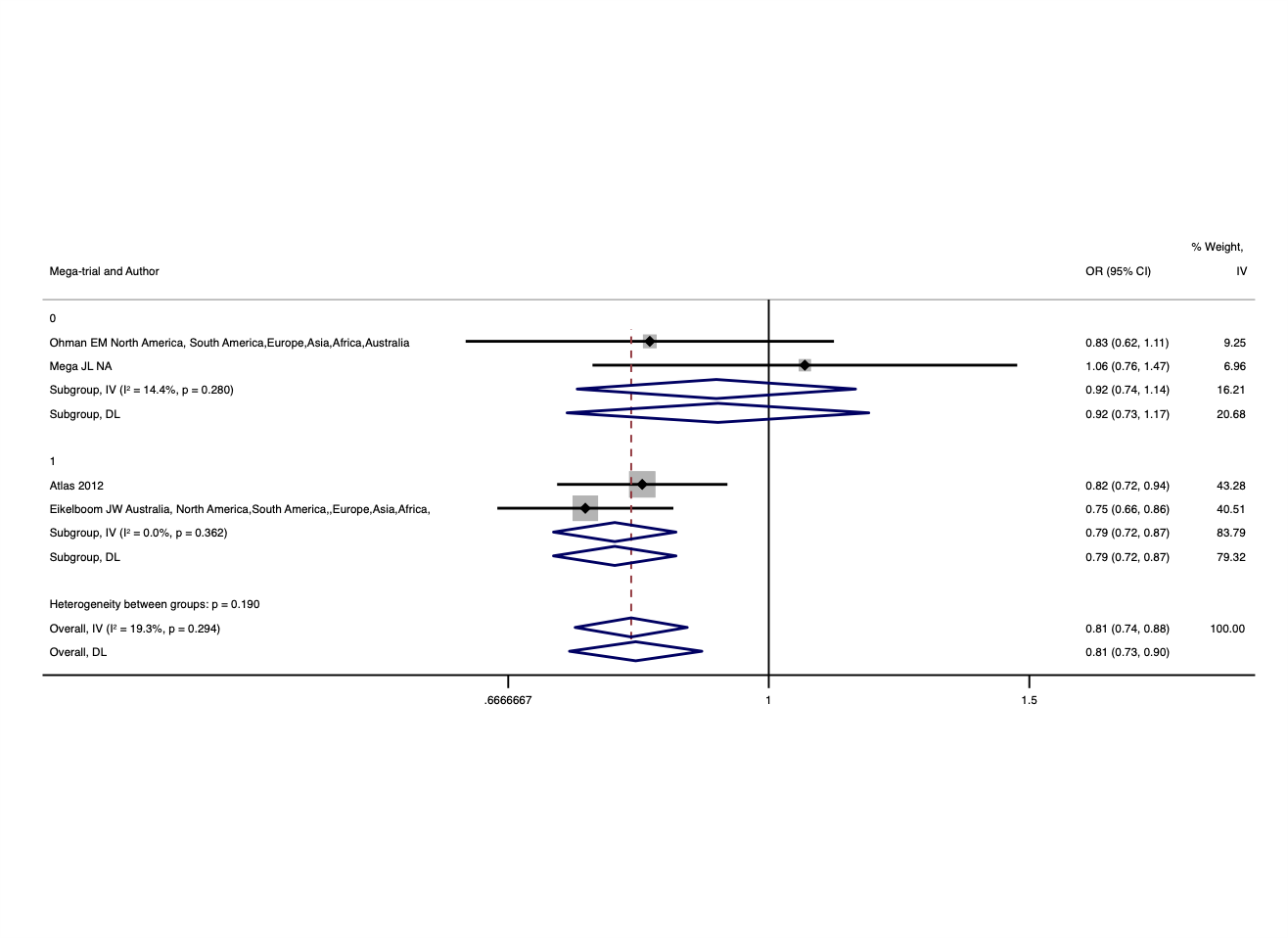

30. **CAMELLIA-TIMI 2018**

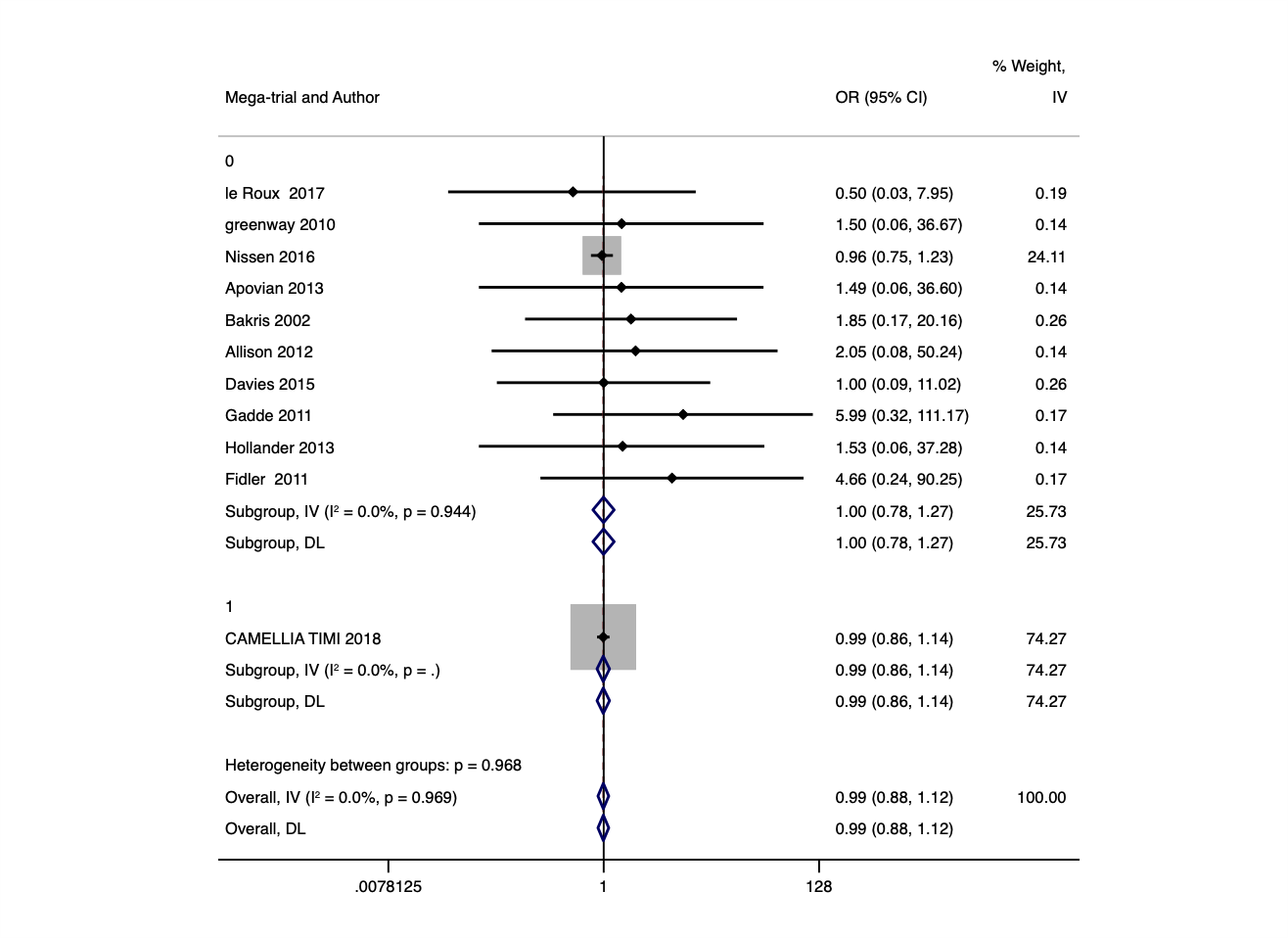

31. **SOLID-TIMI 52 2014**

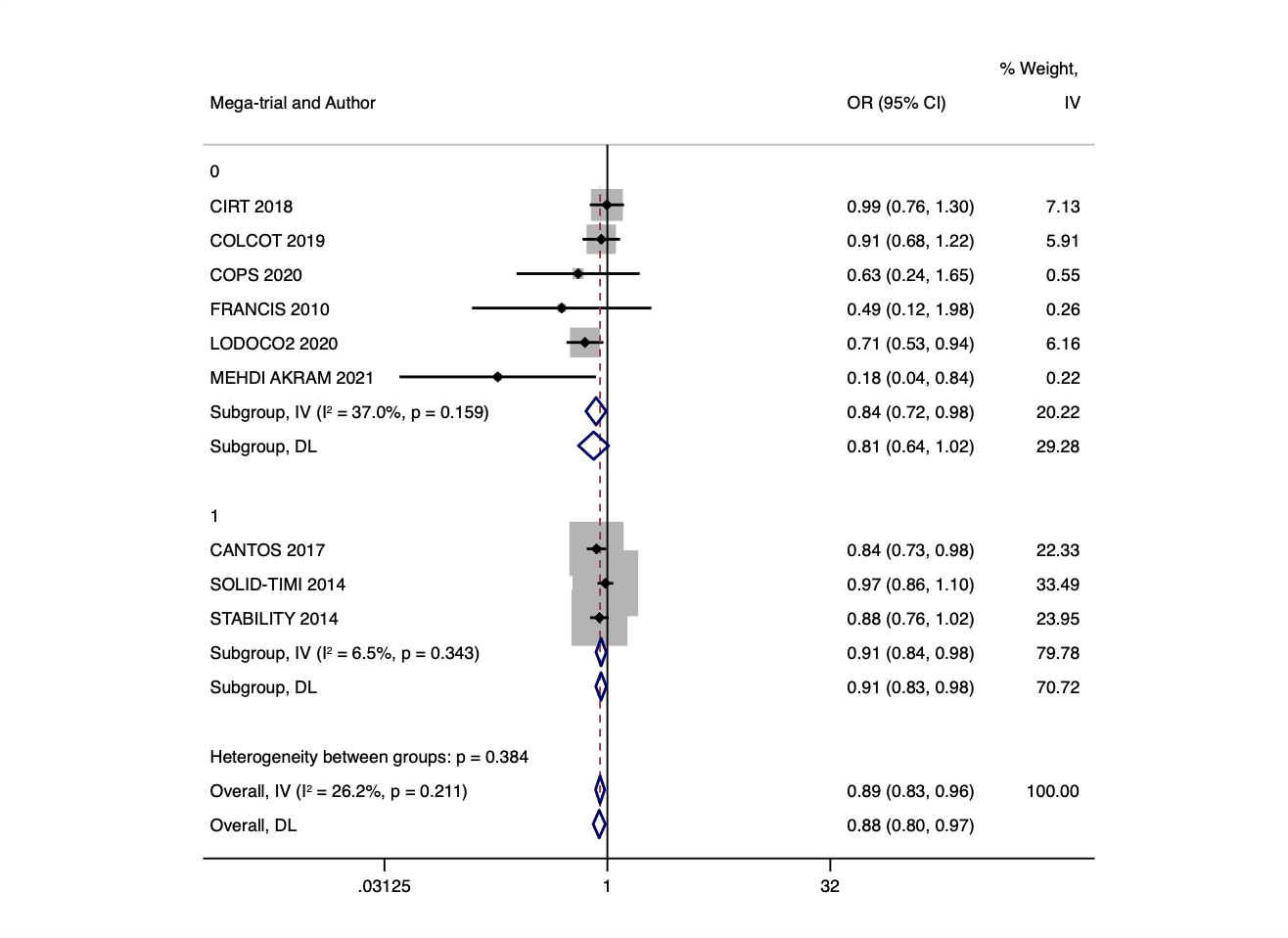

32. **ONTARGET 2008**

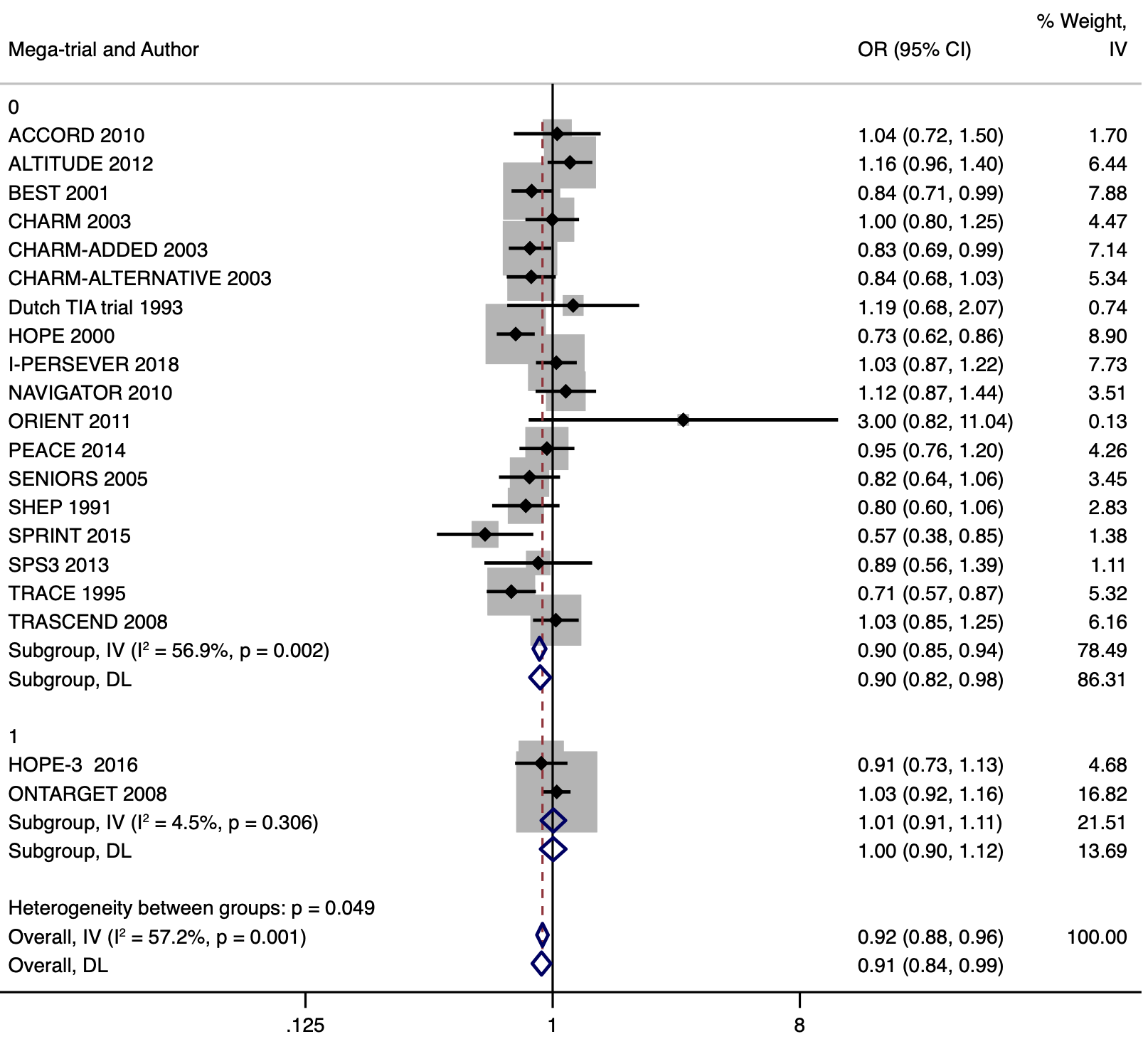

33. **Wallentin 2019**

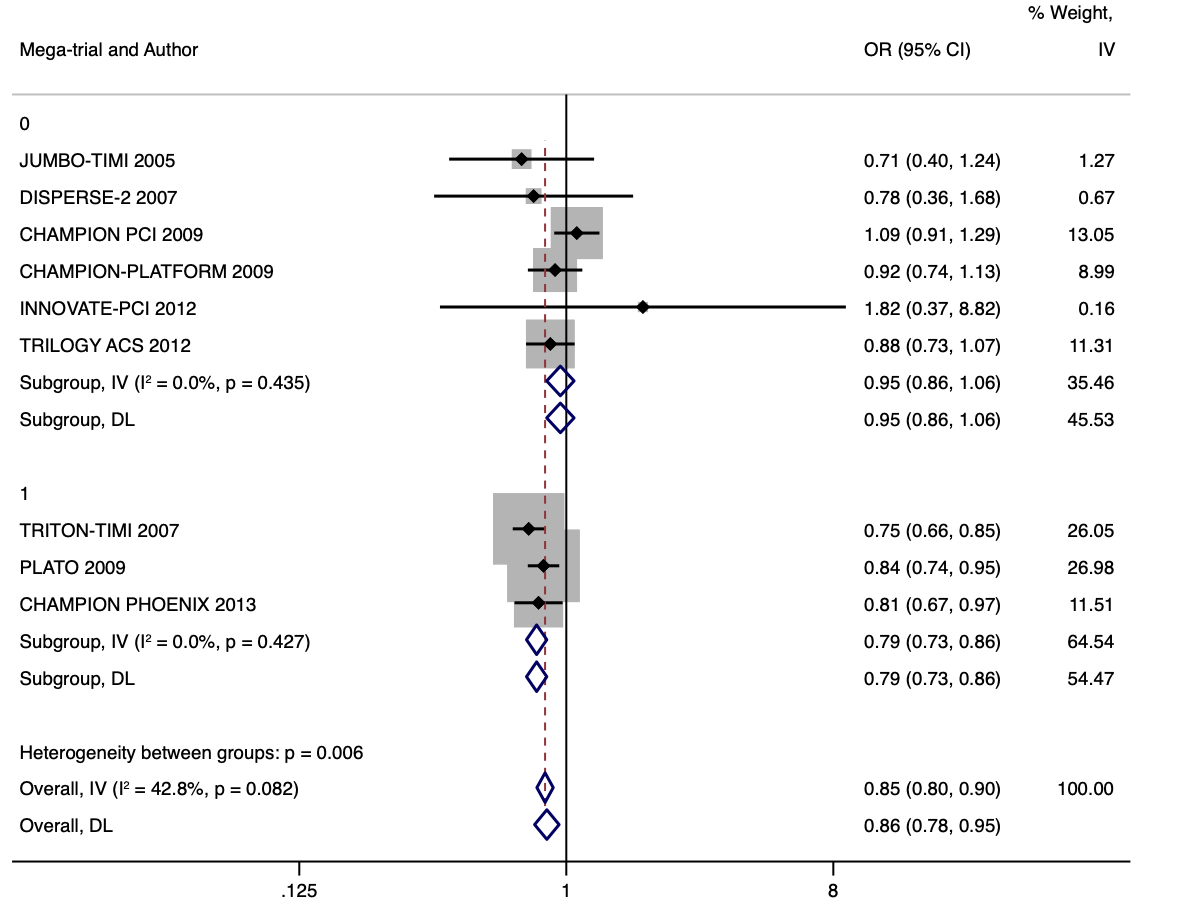

34. **COMMIT 2005**

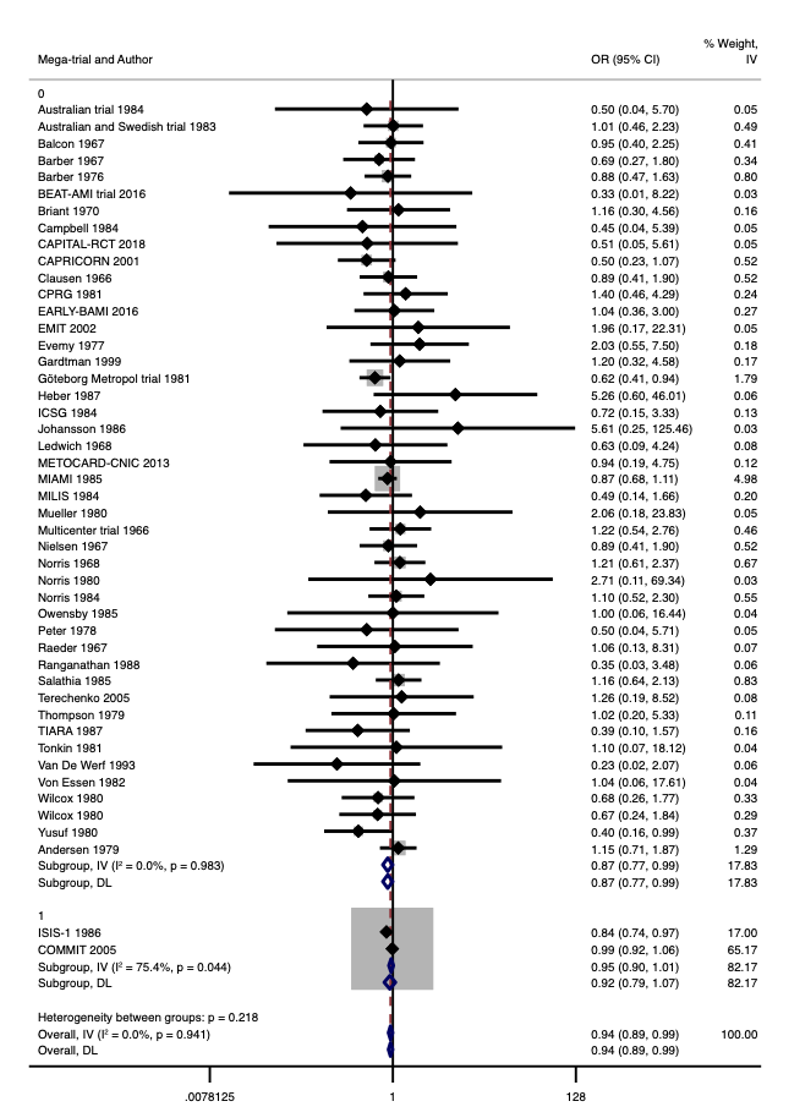

35. **SIGNIFY 2014**

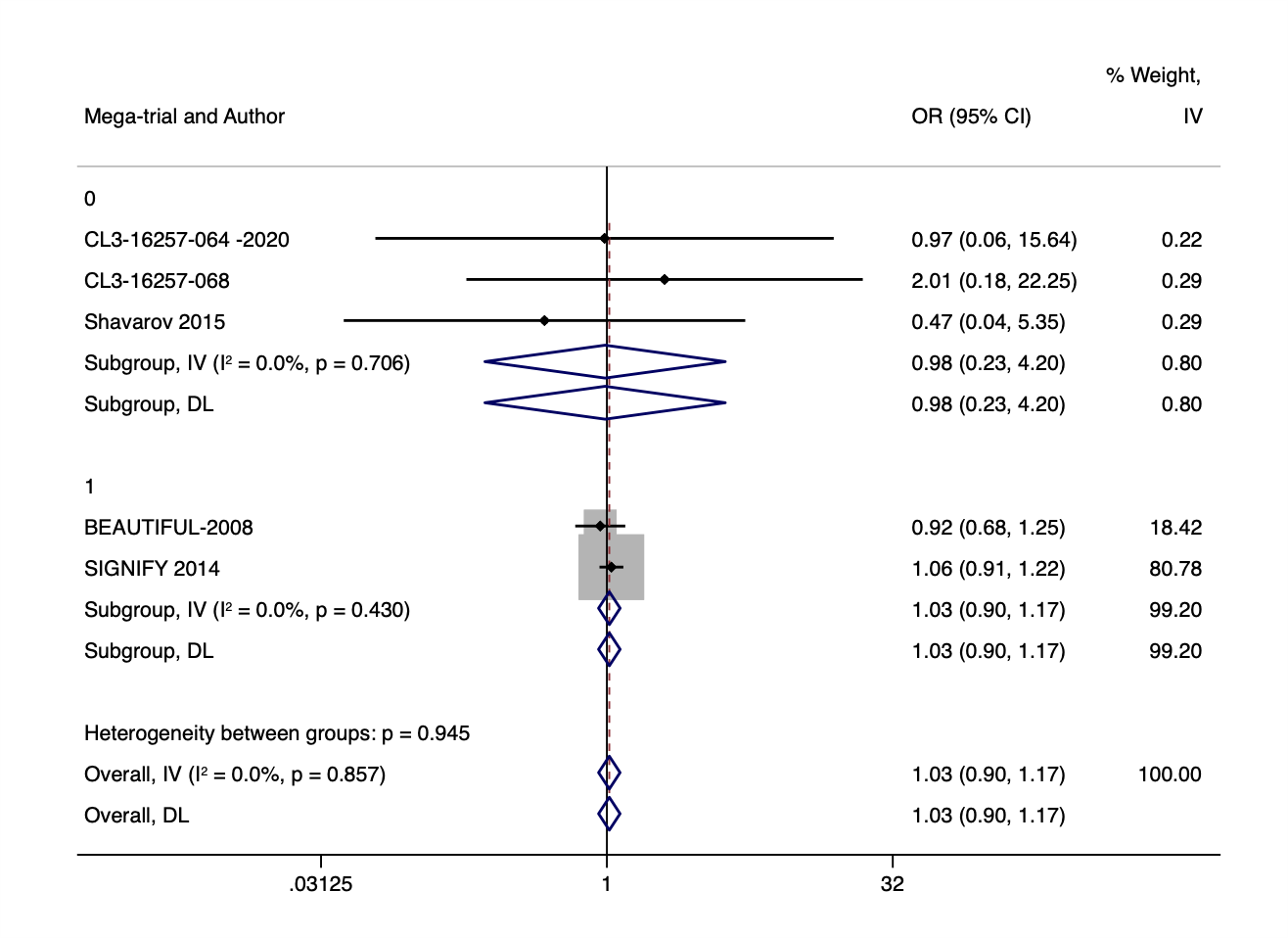
