## Supplementary Material 2 for "Agreement between mega-trials and smaller trials: a meta-research study"

1. **JUPITER 2008**

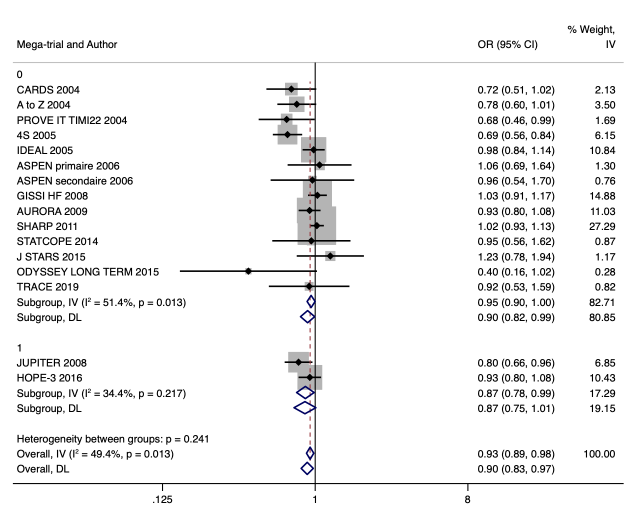

2. **ACCOMPLISH 2008**

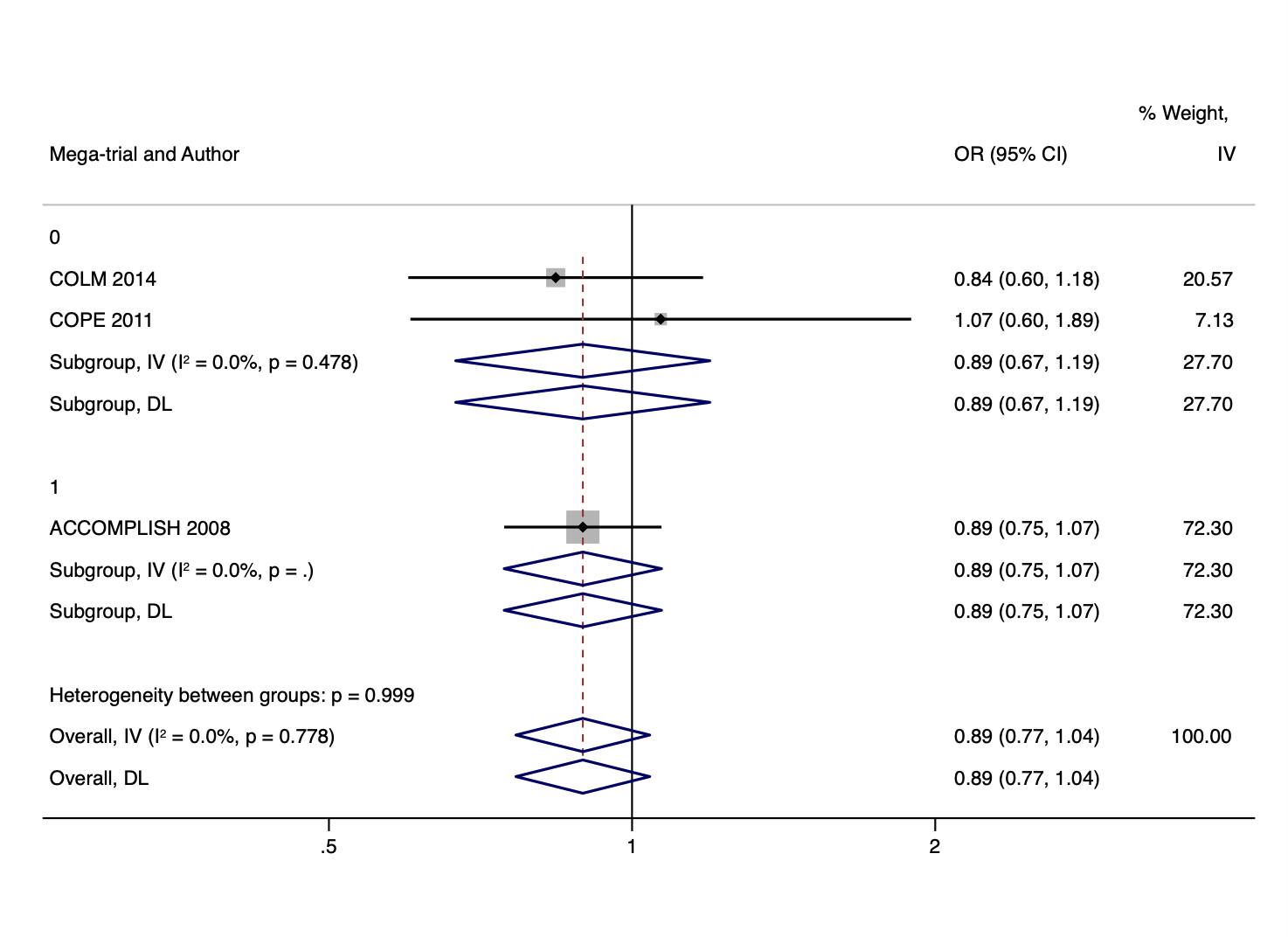

3. **Huo 2015**

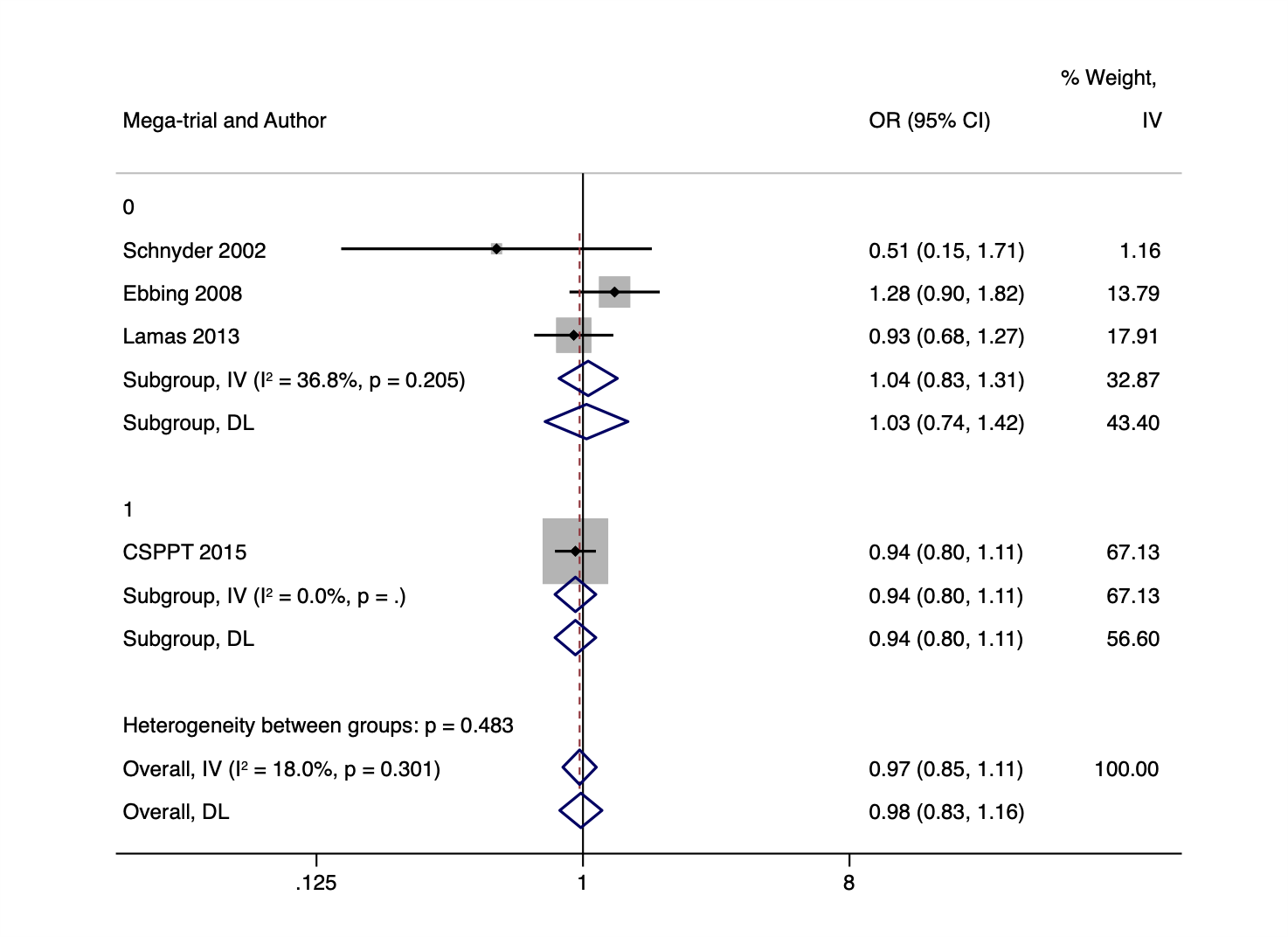

4. **Origin 2012**

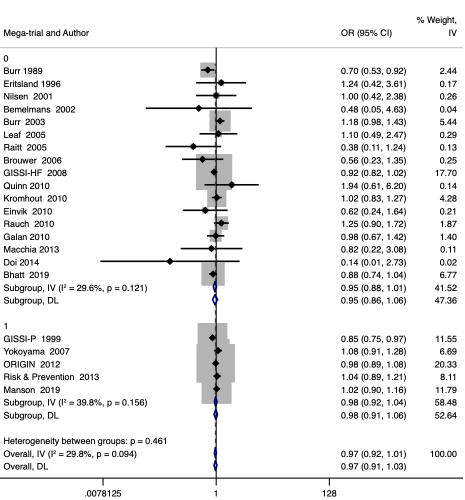

5. **Aberle 2011**

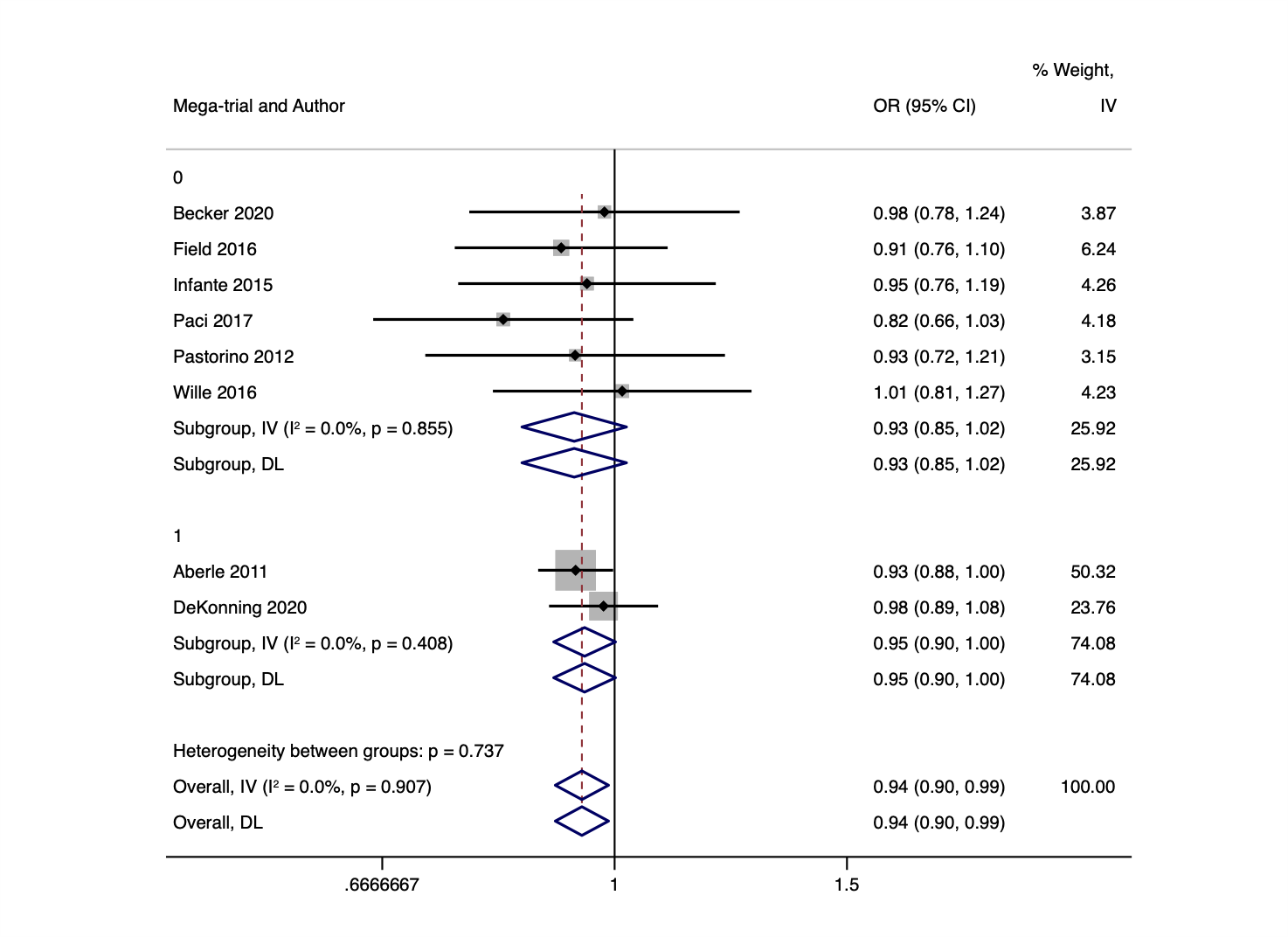

6. **JPP 2014**

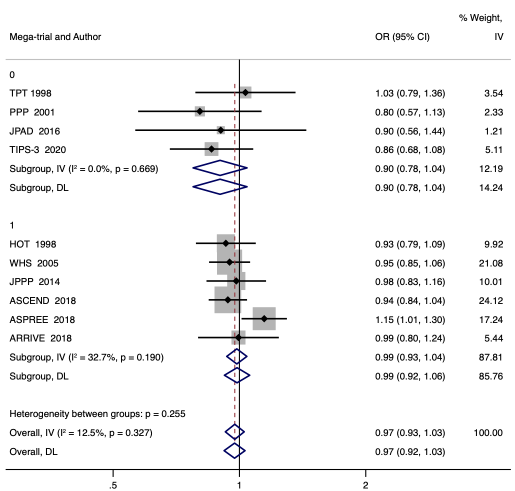

7. **ACCORD 2008**

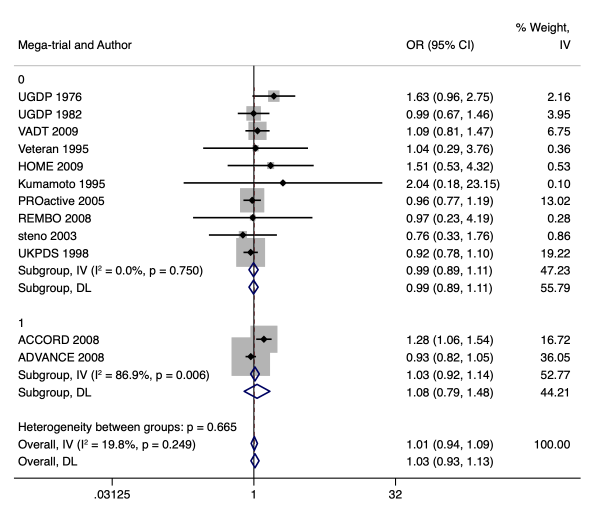

8. **VITAL 2019**

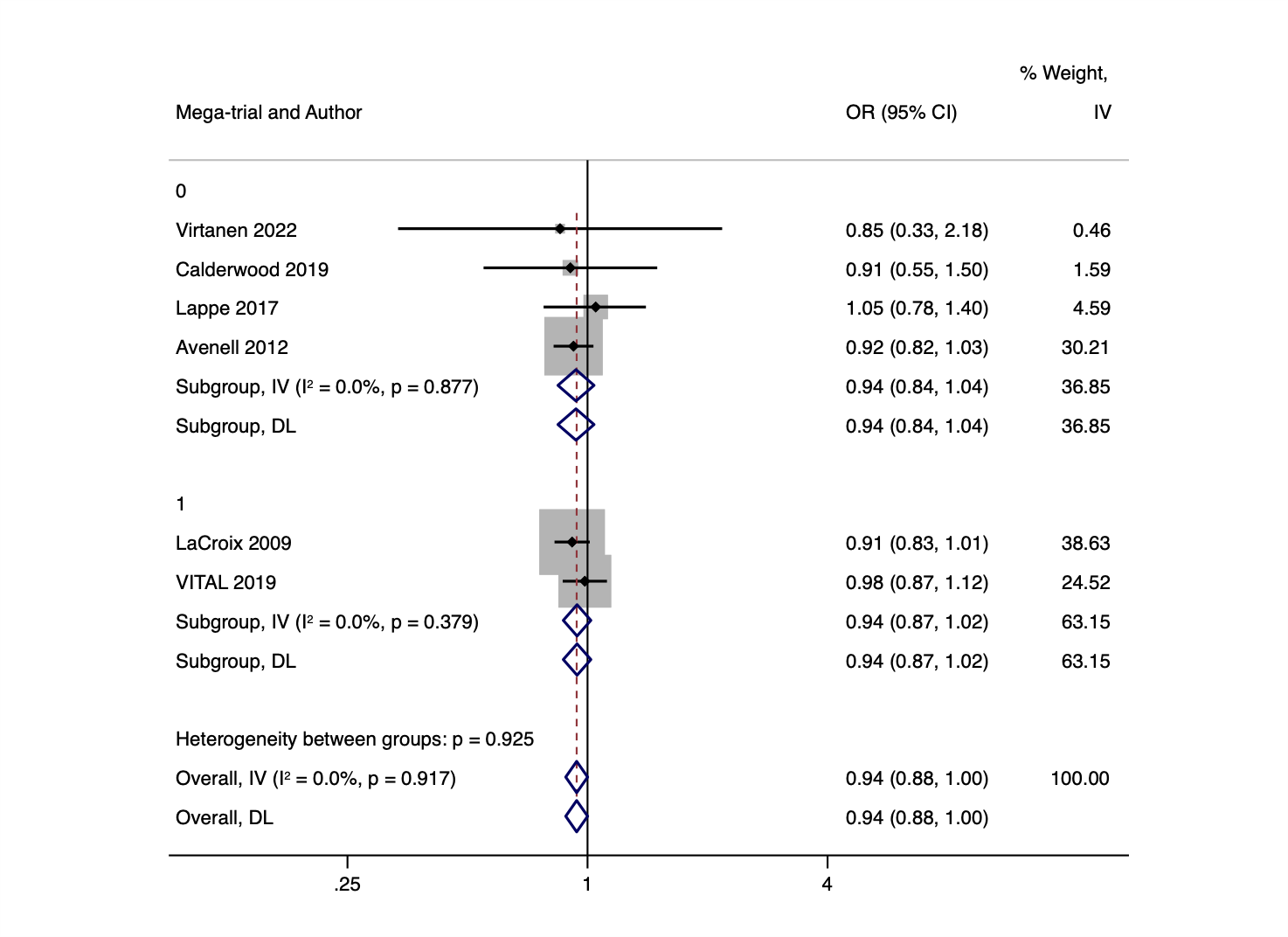

9. **REVEAL 2017**

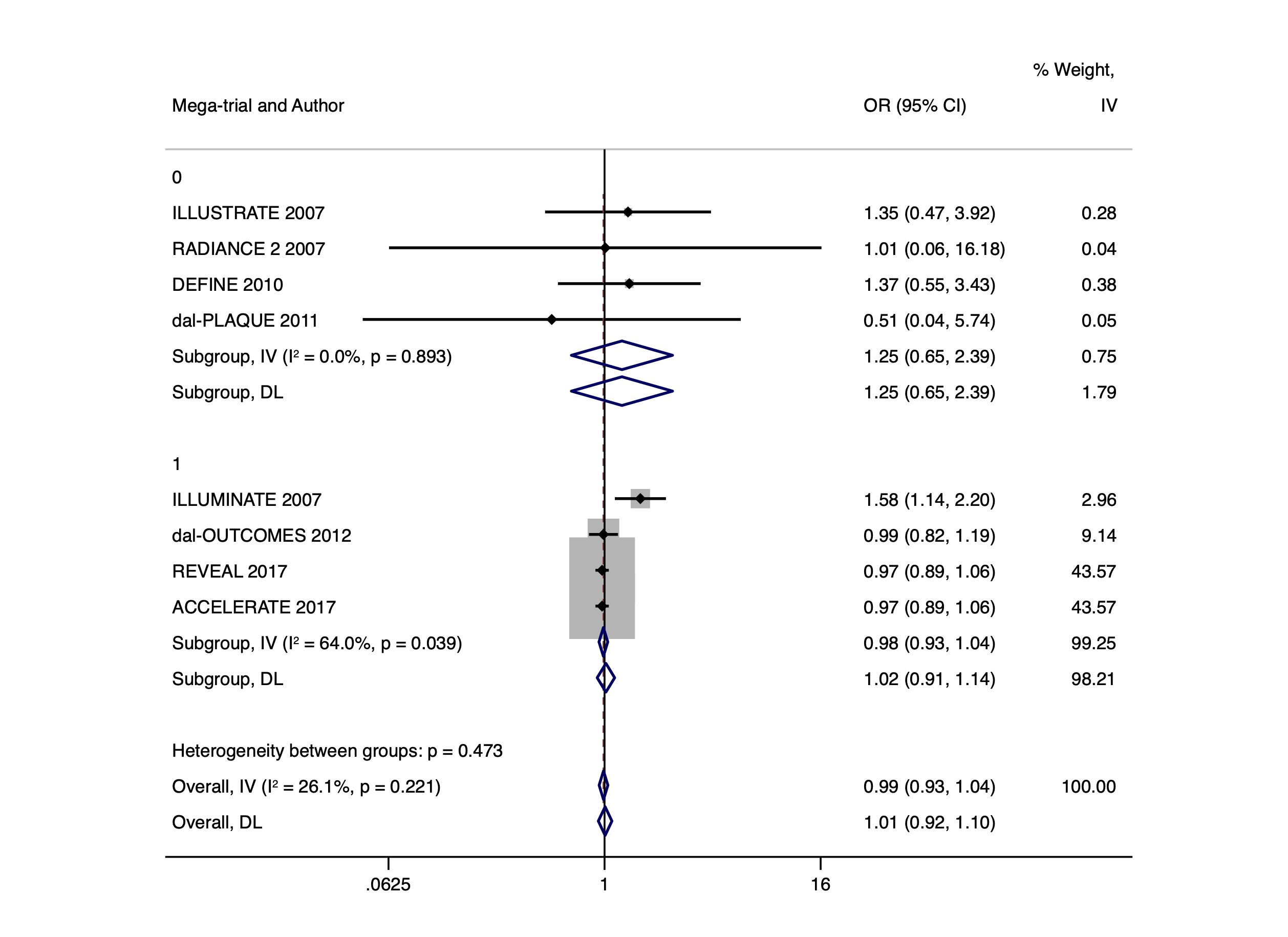

10. **NISSEN 2016**

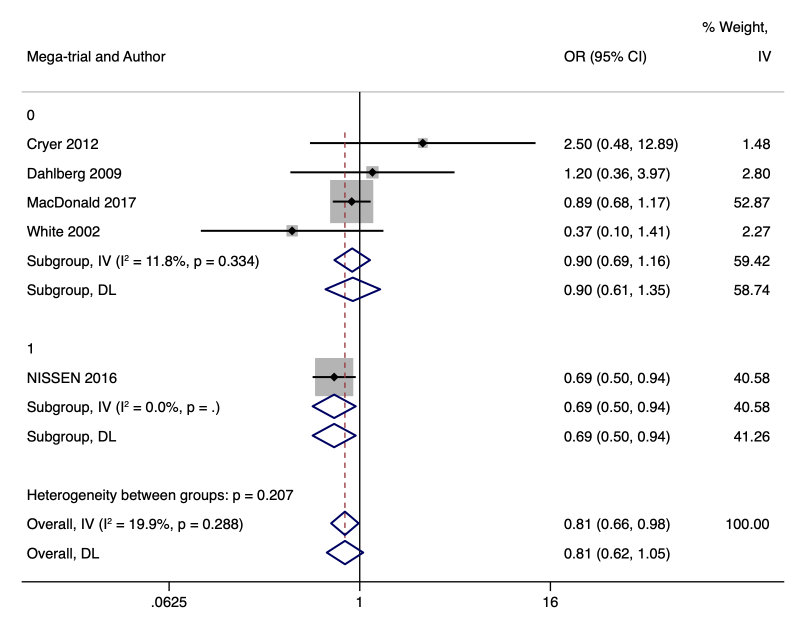

11. **EXTRACT TIMI 25 2006**

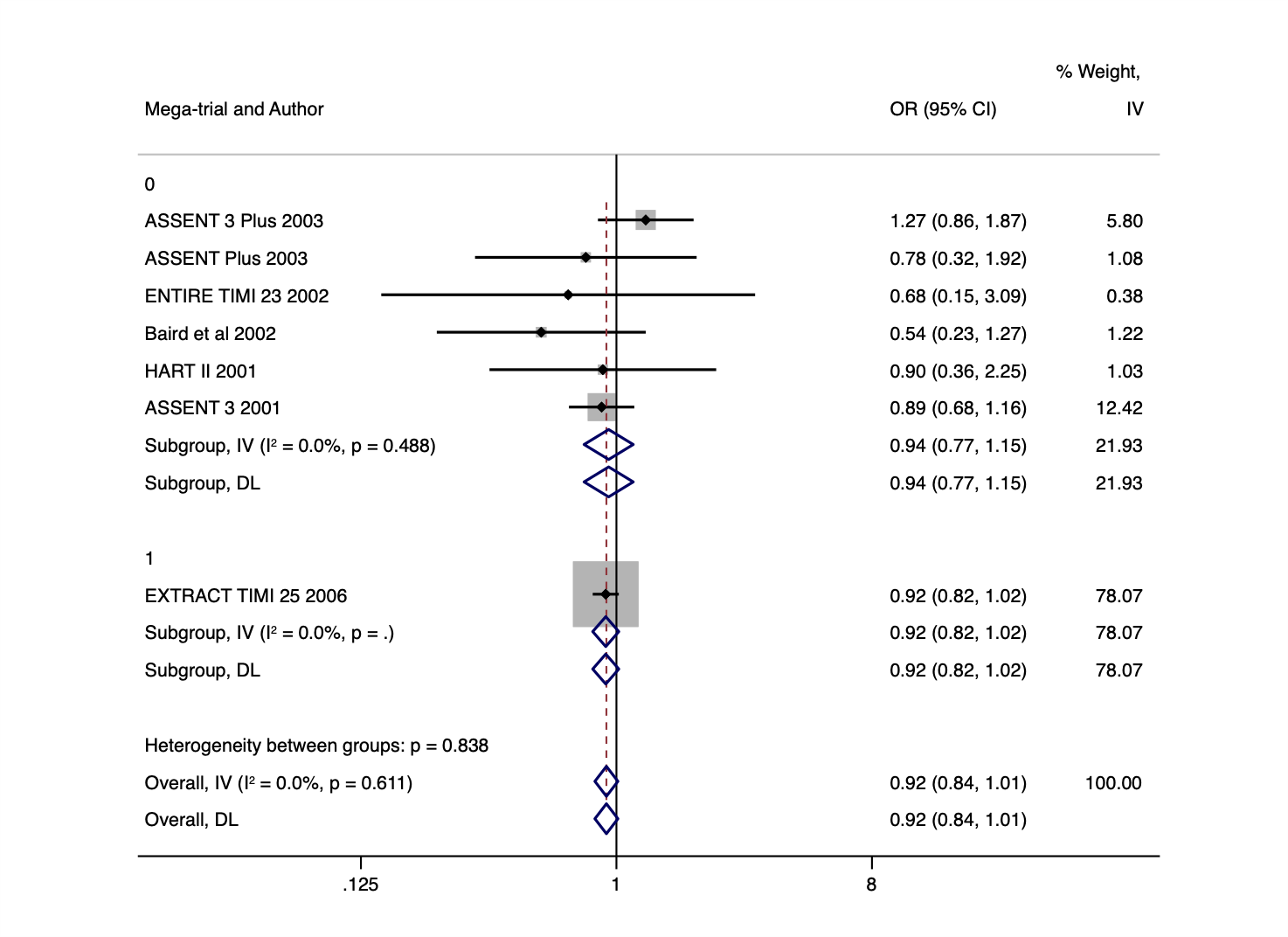

12. **ODYSSEY OUTCOMES**

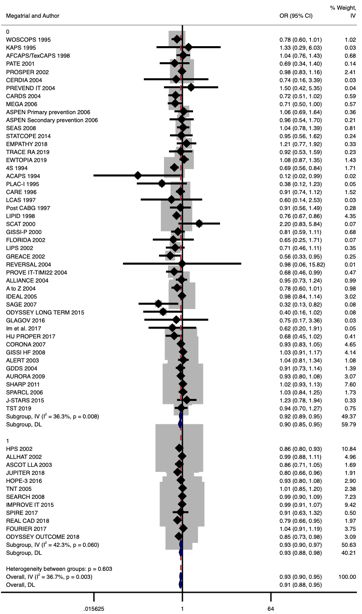

13. **COMMIT 2005**-

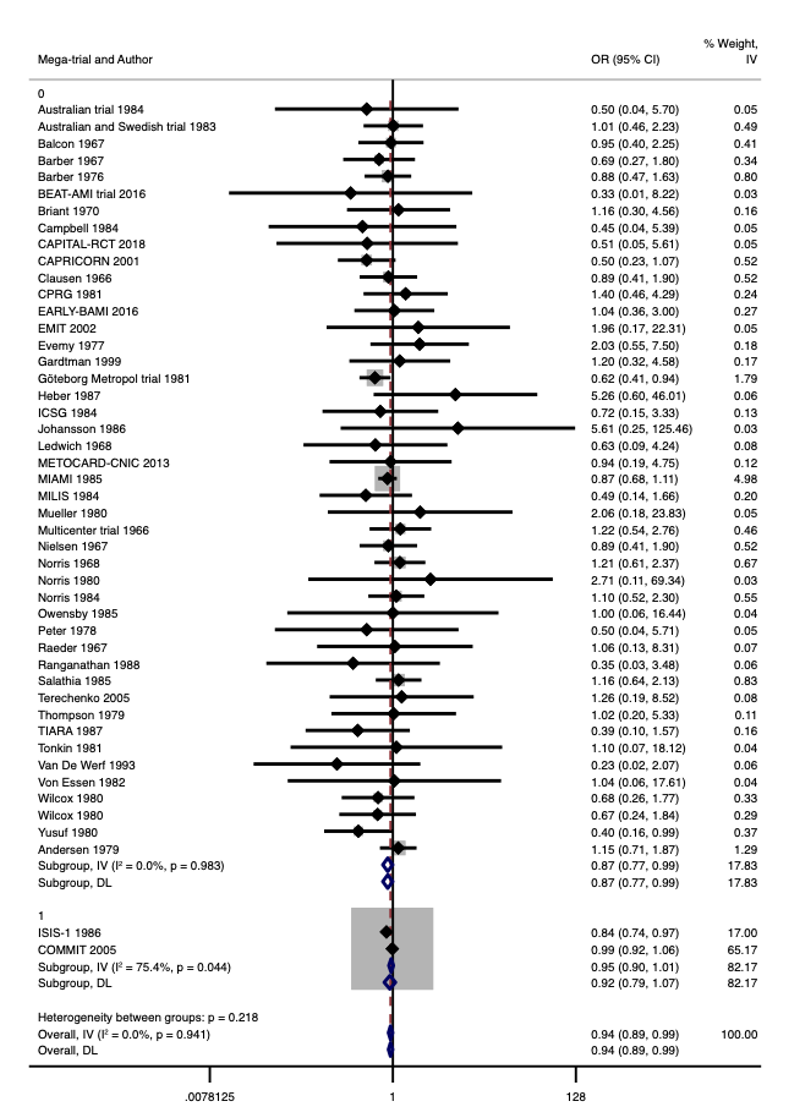

14. **SIGNIFY 2014**

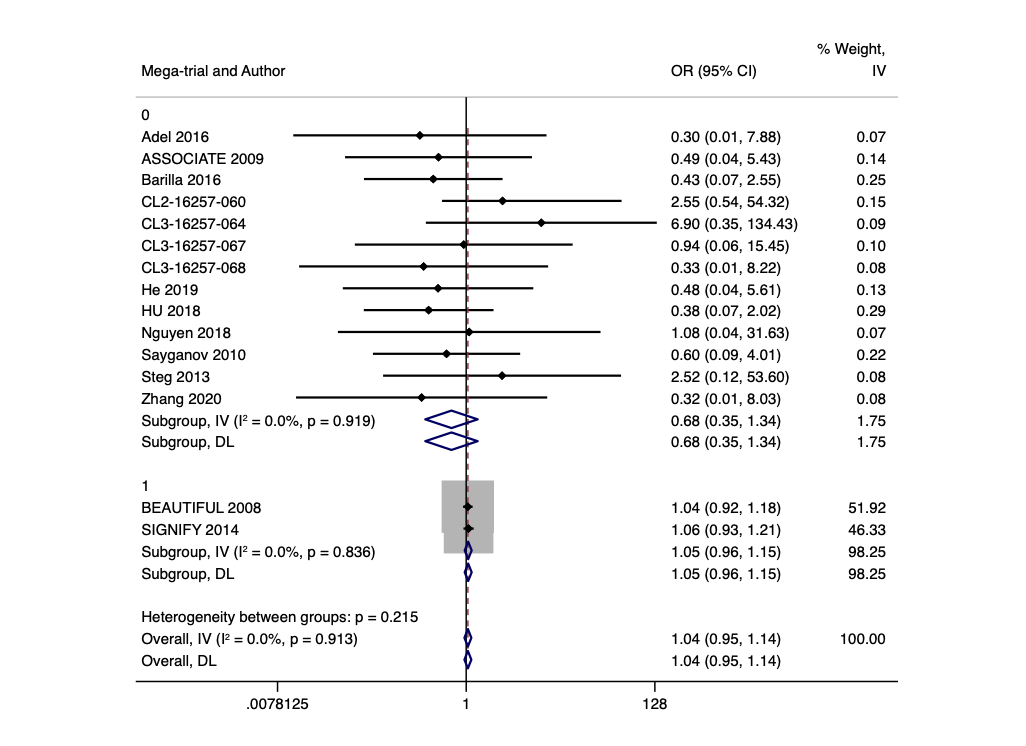

15. **CHARISMA 2006**

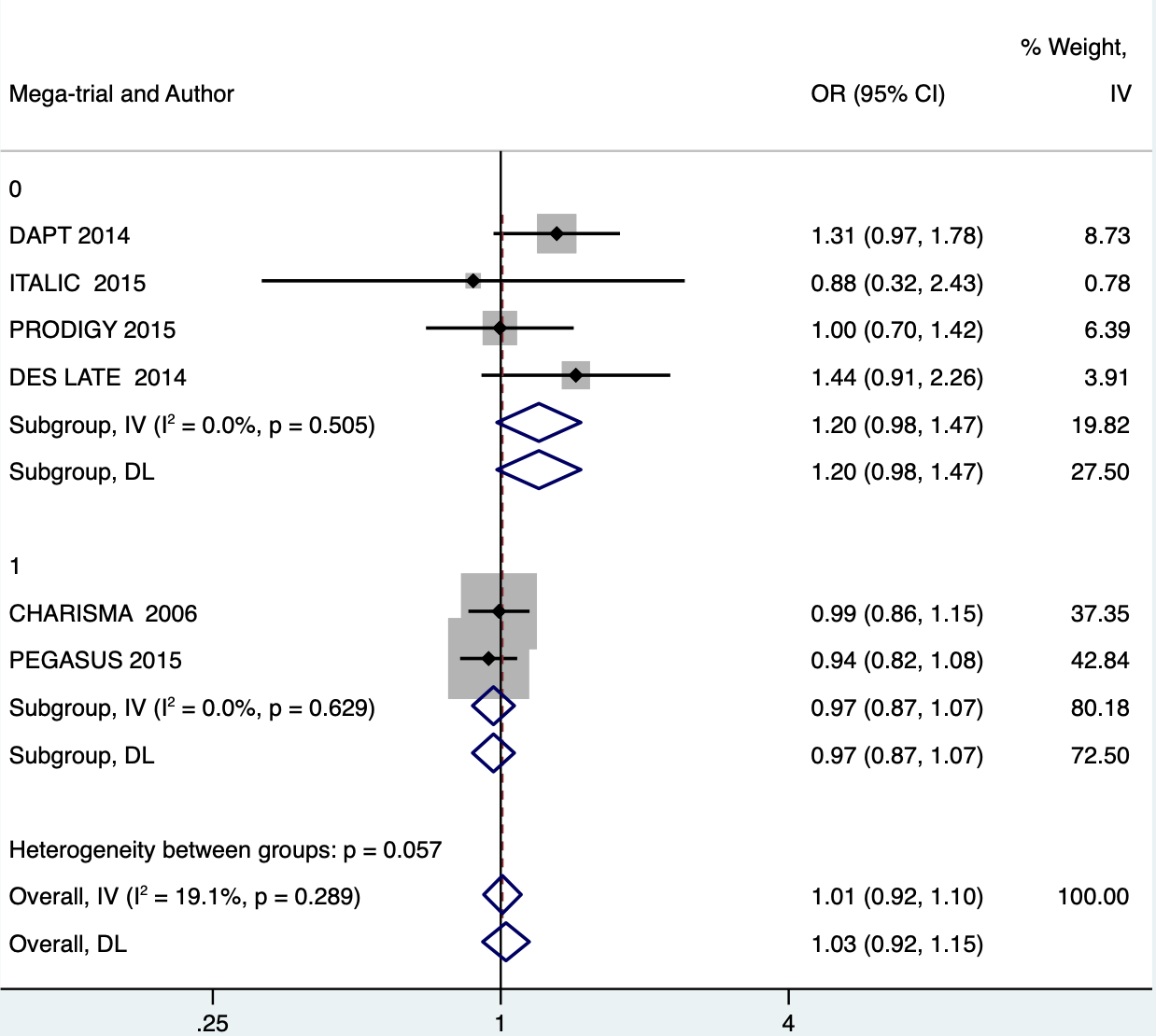

16. **STABILITY**

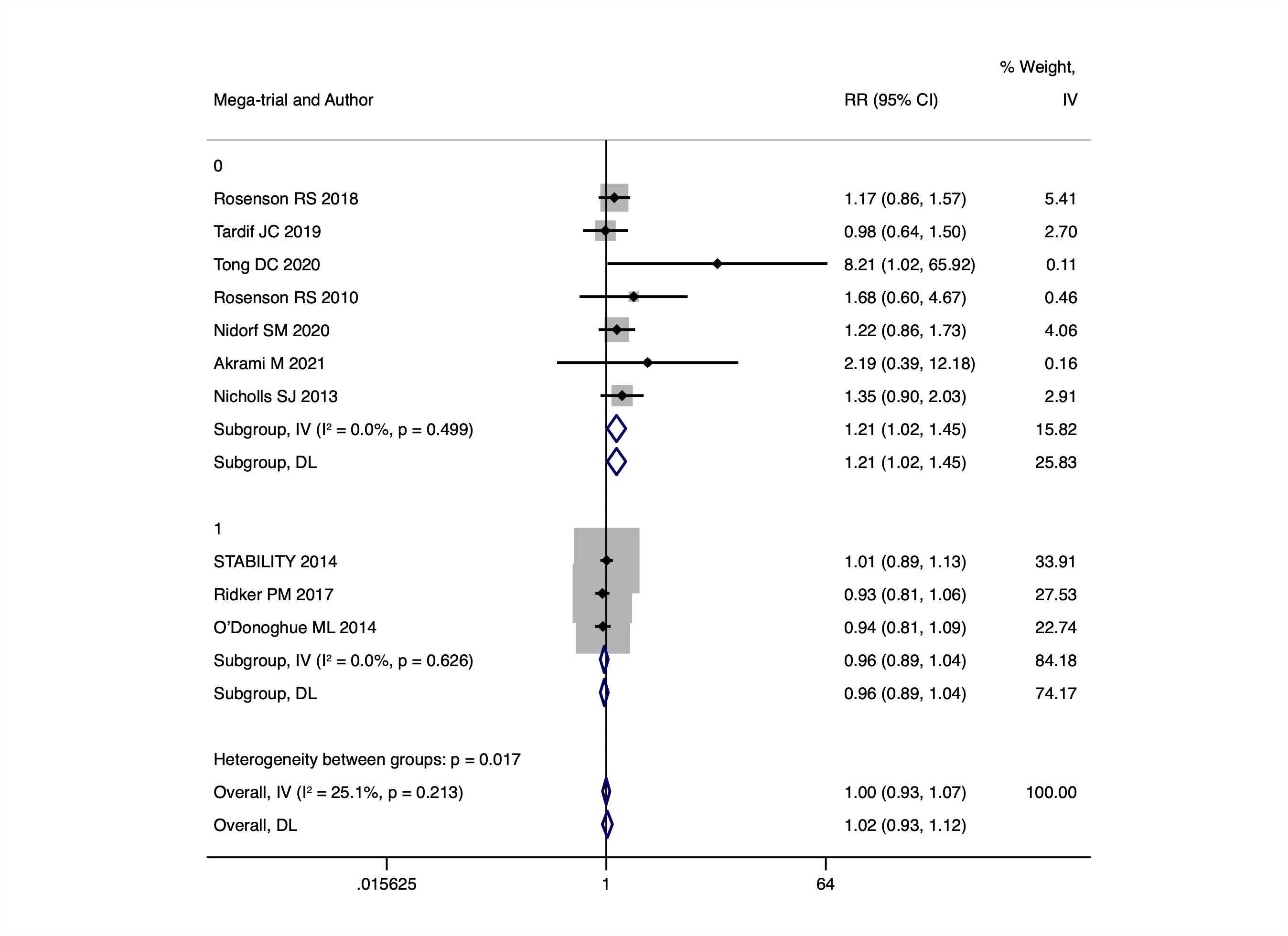

OR

\

17. **POISE-2 2014**
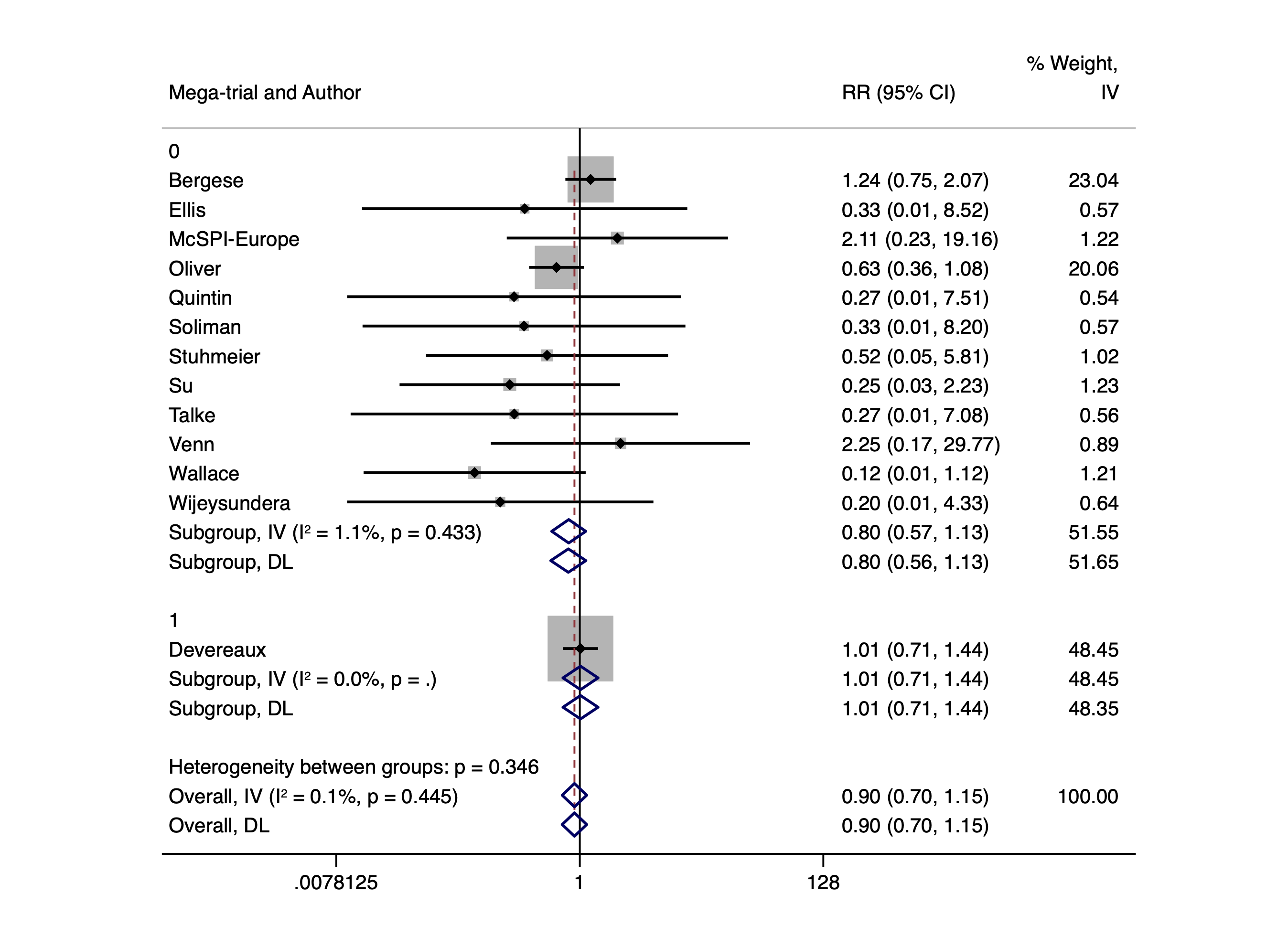

OR

18. **CURRENT OASIS 7- 2010**

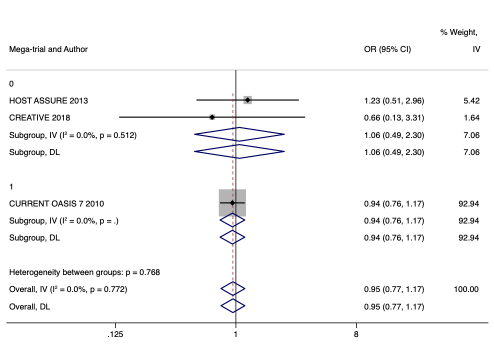

19. **ONTARGET 2008**

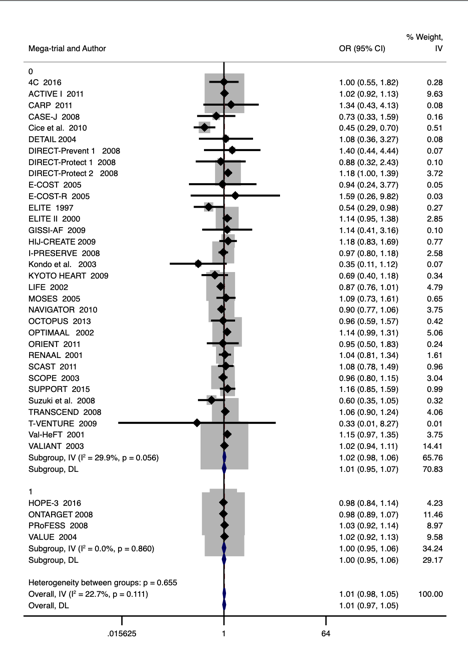

20. **SU.VI.MAX 2004**

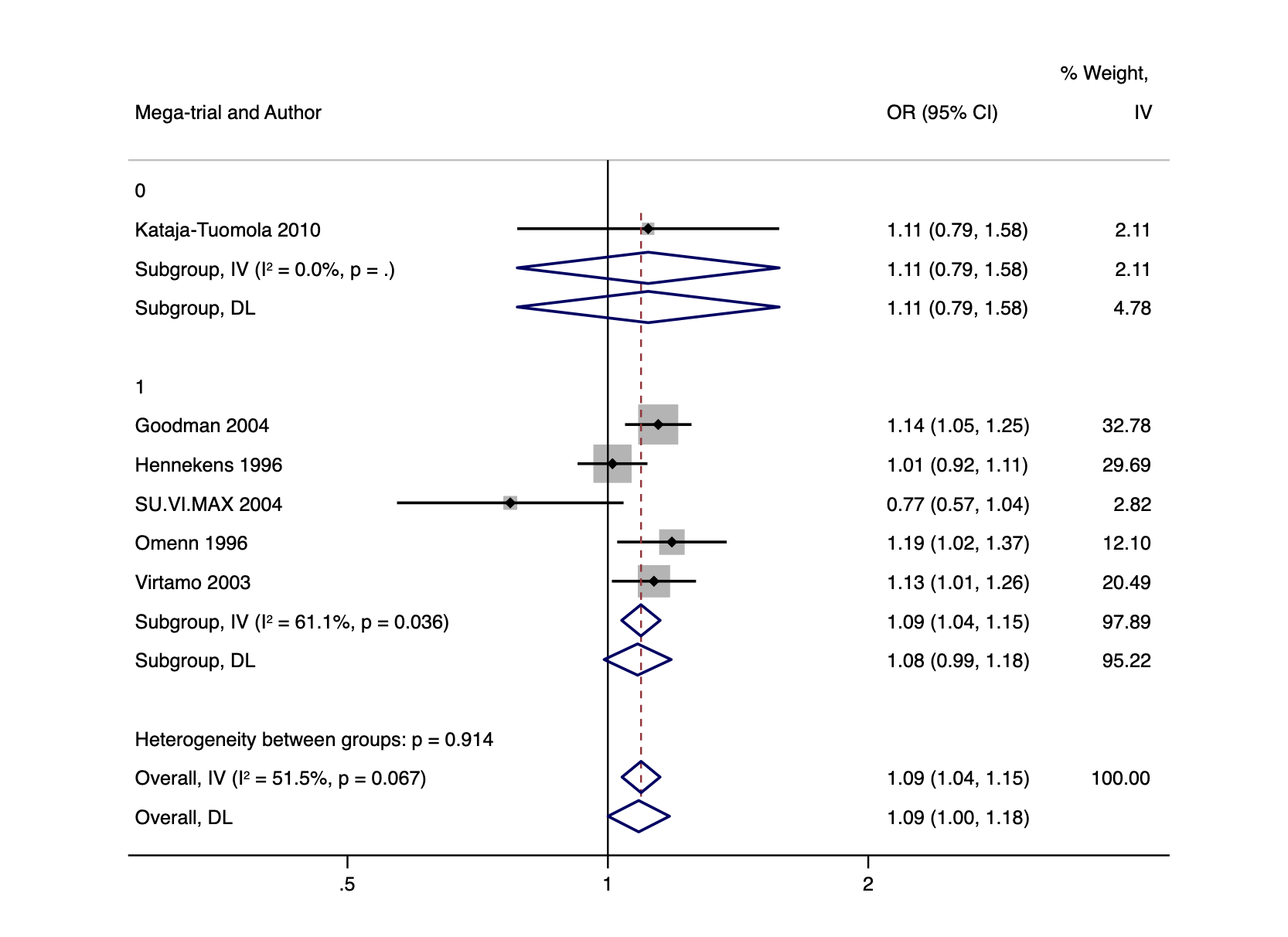

21. **ATLAS 2012**

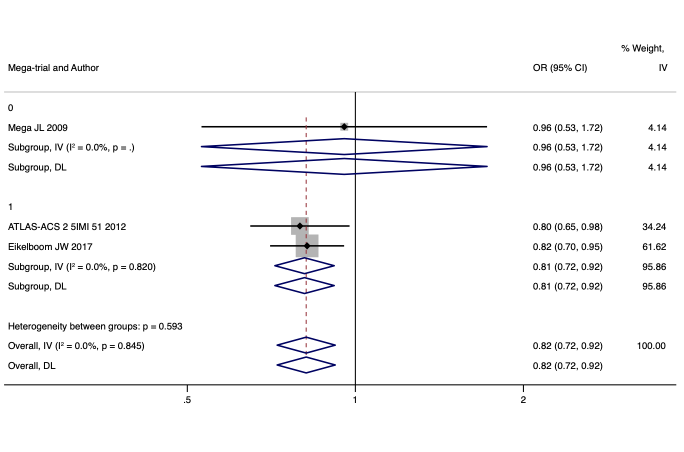

22. **GLOBAL LEADERS 2018**

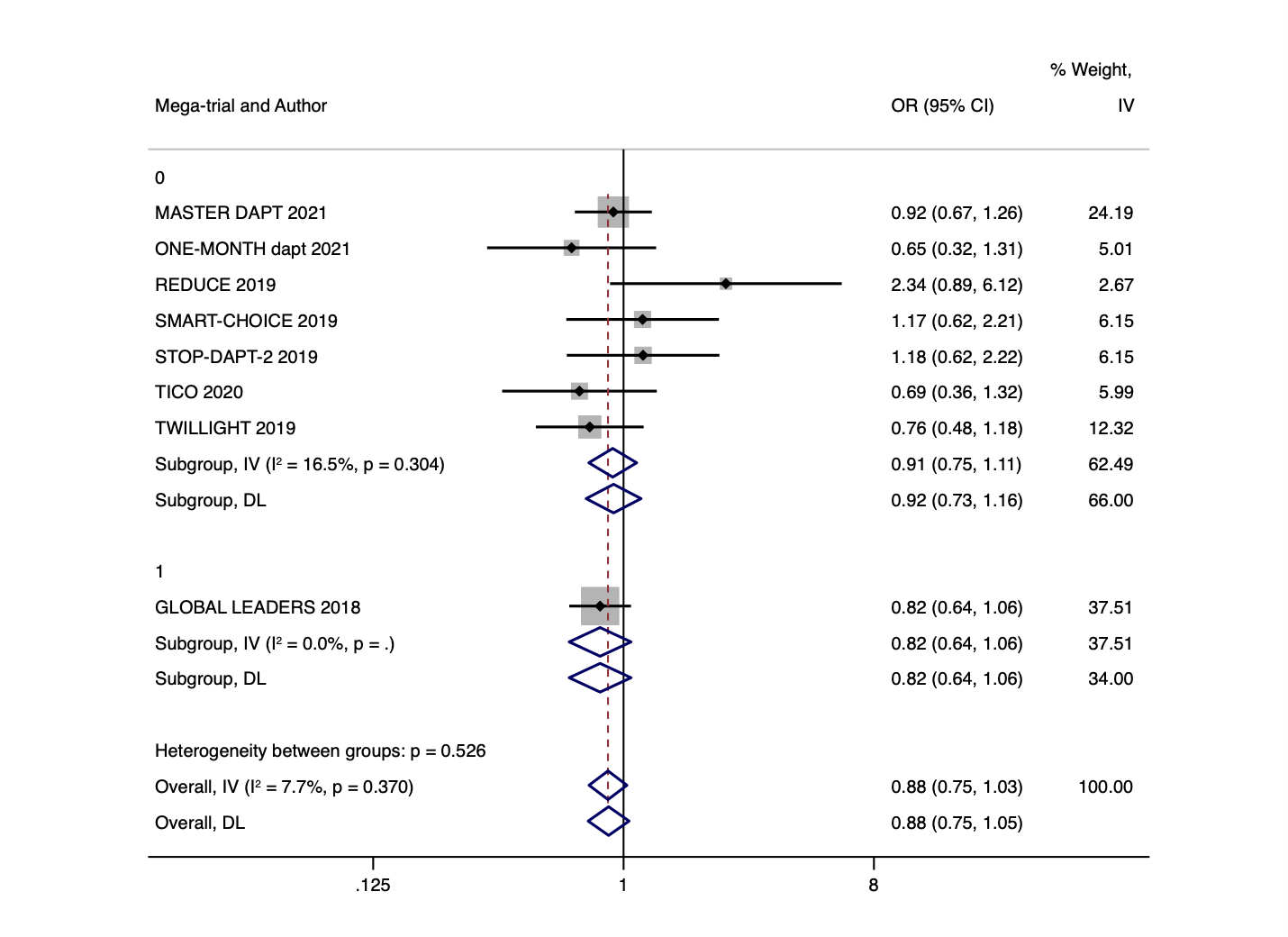

**23 WALLENTIN** – There is no Forest plot for this study as there was only one megatrial and one smaller trial.

24 **SAVOR TIMI-53 2013**

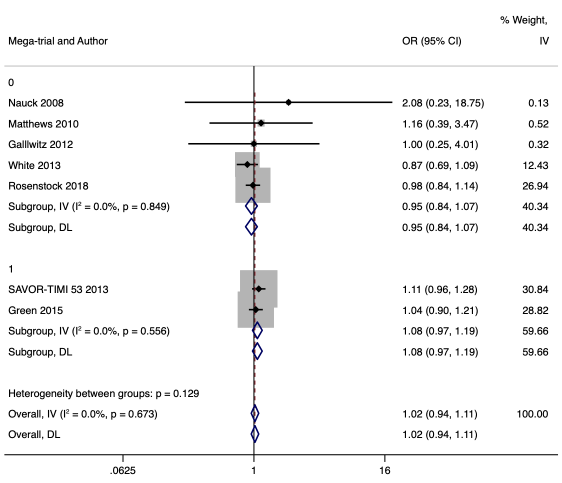

25 **SCORED 2021**

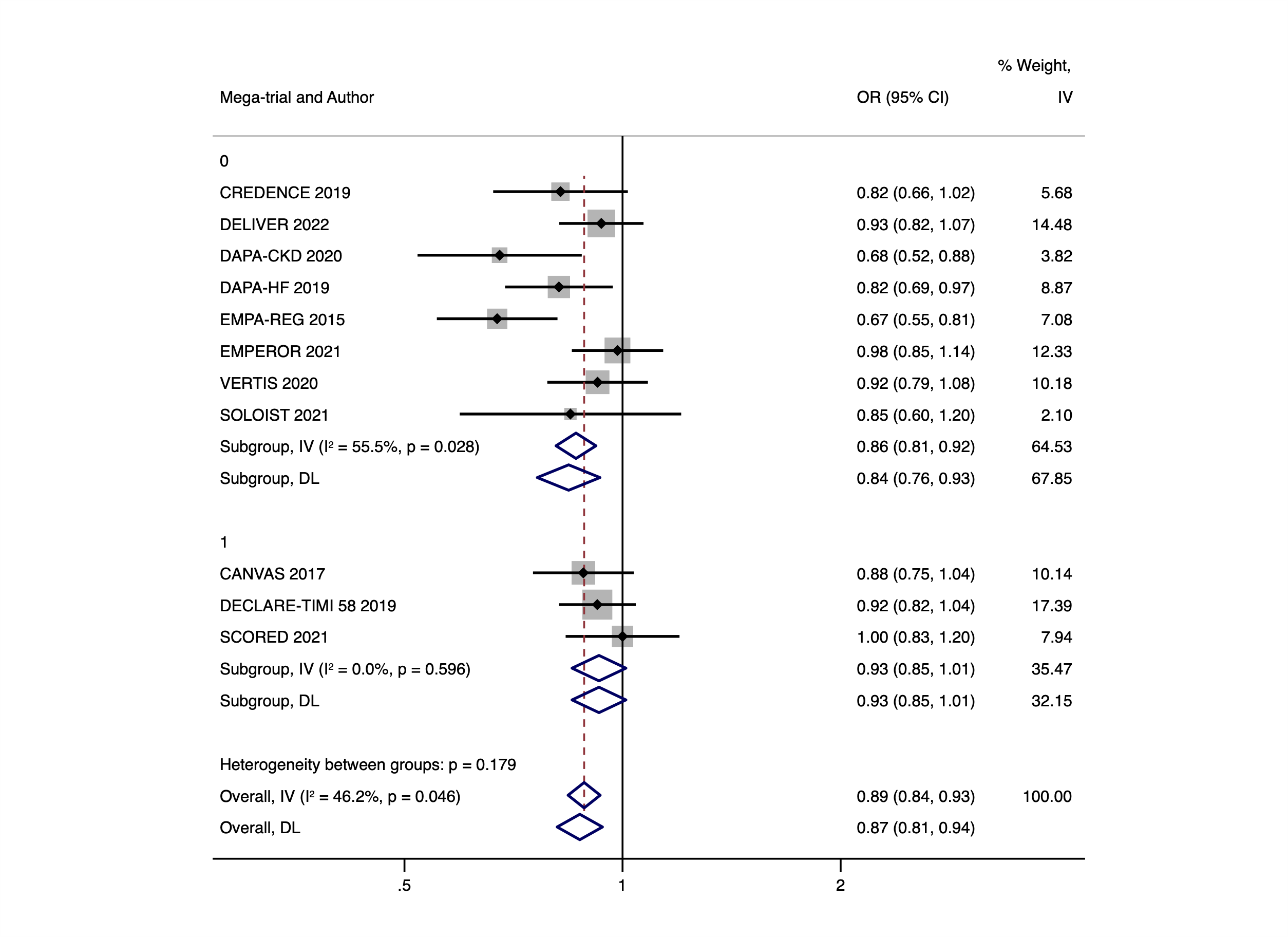

**26. EXSCEL 2017**

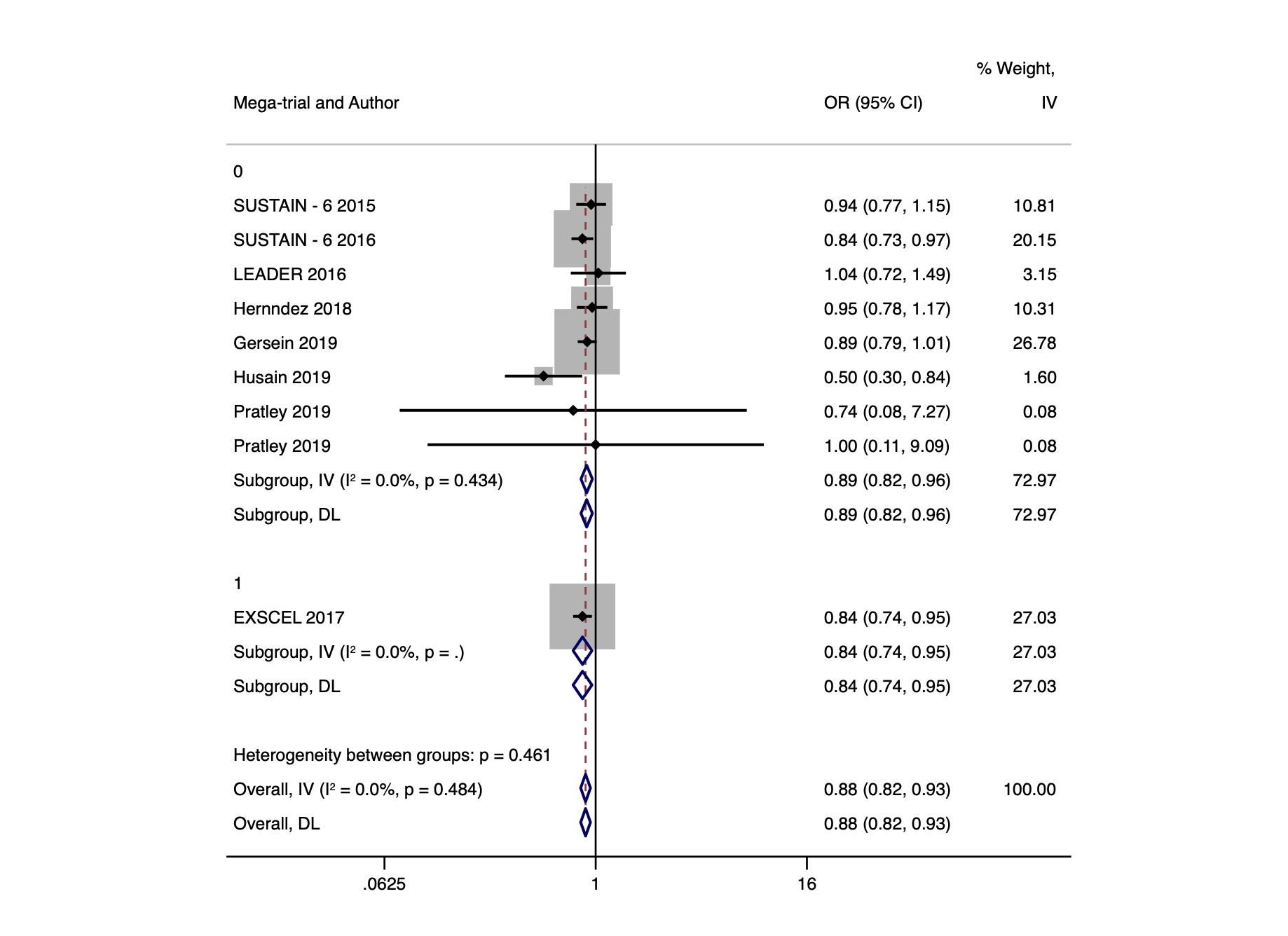
